# Genome-wide association studies to identify shared and distinct mechanisms of fibrosis across 12 organ-systems

**DOI:** 10.64898/2026.02.18.26346458

**Authors:** Ebrima Joof, Tamara Hernandez-Beeftink, Gina Parcesepe, Georgie M Massen, Ritah Nabunje, Hayley J Power, Ruby Woodward, Feras Altunusi, Olivia C Leavy, the DEMISTIFI Consortium, Hilary J Longhurst, R Gisli Jenkins, Jennifer K Quint, Louise V Wain, Richard J Allen

## Abstract

**Introduction:** Fibrosis can affect organs throughout the body and is present in a wide range of diseases. Recent research has suggested that there could be shared biological mechanisms that lead to fibrosis in different organs.

**Methods:** We performed genome-wide association studies using UK Biobank for fibrosis in 12 different organ-systems and meta-analysed results with previously published studies of fibrotic diseases. We considered genetic associations that colocalised across ≥3 organs as those likely to be involved in general fibrotic mechanisms and also identified novel genetic variants not previously reported as associated with fibrosis. Genetic correlation of fibrosis between organs was calculated using linkage disequilibrium score regression (LDSC). Discovery analyses were performed using European ancestry individuals and results were tested further in African, South Asian and East Asian ancestry groups.

**Results:** We identified eight genetic loci that colocalised across three or more organs. One of these signals, located near the *SH2B3* and *ATXN2* genes, showed evidence of a shared causal variant for fibrosis across five organs. We also identified two novel fibrotic associations, one implicating alternative splicing of *TFCP2L1* for urinary fibrosis and another implicating a missense variant in *FAM180A* for intestinal-pancreatic fibrosis. We observed significant genetic correlations for all organs, particularly for liver and skeletal fibrosis.

**Conclusion:** We found evidence of shared genetic associations for fibrosis across organs, both at individual genetic loci and genome-wide. This highlights specific genes that may contribute to fibrosis across organs and diseases, which may facilitate the development of new therapies.

## Introduction

Fibrosis, the scarring or thickening of tissue, can occur in most organs^1^. Fibrotic-related diseases are estimated to account for 18-35% of deaths globally^2, 3^. There has been accumulating evidence that fibrosis mechanisms share similarities across organs and diseases^4^. Understanding the shared and distinct drivers of fibrosis across organs and diseases could lead to large therapeutic impacts, given its high overall burden.

Although, the triggers of fibrosis vary between organs, severe or repetitive tissue injury leading to excessive accumulation of persistent fibroblast populations, integrin-mediated activation of TGF-β signalling, cytokine-mediated recruitment and activation of immune cells, and metabolic reprogramming represent core pathogenic mechanisms shared across multiple organs^5^. Examples of shared risk factors include external factors that cause inflammation, body mass index (BMI) and telomere length, which are reported as associated with fibrosis in the lung, liver, heart, kidney, joints and biliary systems^6-12^.

One method for identifying biological mechanisms important for disease is through genetic studies. Genome-wide association studies (GWAS) involve testing genetic variants individually from across the genome for their association with disease. There have been previous GWAS that investigated individual fibrotic diseases^13-22^. A GWAS of T1 mapping (a measure of interstitial fibrosis derived from Magnetic Resonance Imaging (MRI) scans) in the liver, pancreas, kidney and heart found modest genetic correlation between the T1 GWAS across organs and several overlapping genetic signals associated with fibrosis in multiple organs^23^. We hypothesised that by identifying genetic determinants that are associated with development of any fibrotic disease within an organ, rather than investigating individual fibrotic diseases, we could identify shared and distinct biological mechanisms that drive fibrosis across organs.

## Methods

### Overview

We performed GWAS of fibrosis separately in each organ in a large population biobank (UK Biobank), meta-analysed results with previous GWAS of individual fibrotic diseases and then performed functional follow-up analyses to characterise shared fibrotic mechanisms (**Figure 1**).

**Figure 1:**
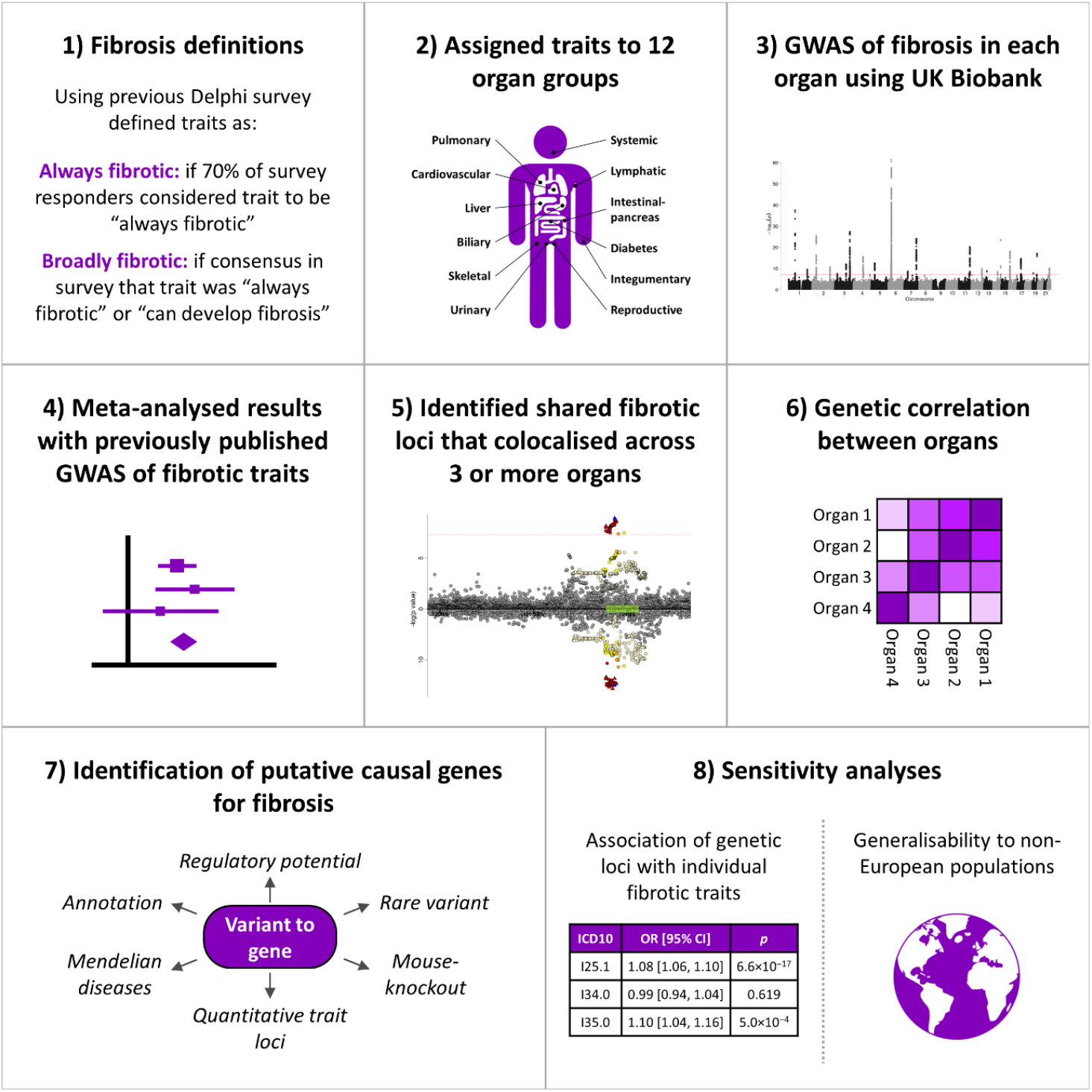
Study overview.

### Defining fibrosis

We used two definitions of fibrosis. We defined diseases as ‘broadly fibrotic’ if there was clinical consensus the disease was either “always fibrotic” or “can develop fibrosis” from a previous Delphi survey^24^. We defined diseases as ‘always fibrotic’ if ≥70% of respondents of the Delphi survey considered the disease “always fibrotic”. These diseases were then assigned to 12 organ-systems (biliary, cardiovascular, diabetes, intestinal-pancreas, liver, lymphatic, pulmonary, reproductive, skeletal, skin, systemic, urinary) and mapped to Systematised Nomenclature of Medicine Clinical Terms (SNOMED-CT) and International Classification of Disease 10th Revision (ICD-10) codes (**Supplementary Table 1**). ‘Always fibrotic’ diseases were only found in the biliary, liver, pulmonary and skin organ-systems.

### UK Biobank GWAS

We performed GWAS for the ‘broadly fibrotic’ and ‘always fibrotic’ case definitions in each organ separately using UK Biobank (a cohort study of 500,000 participants)^25^. For each organ group, cases were defined as individuals with at least one fibrotic disease recorded (as ICD-10 codes) in their hospital episode statistics (HES) or mortality records. We used only HES and mortality records to ensure consistency of case definition across phenotypes and minimise self-reporting bias. The HES data included records from both admission and non-admission encounters. We selected 10 controls per case matched on age and sex and excluded individuals that withdrew consent, failed genotyping quality control, had a genotyping call rate<95%, high heterozygosity or were sex mismatches. For related individuals (2^nd^ degree or closer) the individual with the lower genotype call rate was excluded. Individuals were assigned to ancestry groups as has been described previously^26^. Briefly, k-means clustering was performed on the first two genetic principal components and four clusters were chosen to infer ancestry grouping (European, African, East Asian and South Asian). Analyses were performed in each ancestry group separately and we used the European individuals for our primary analysis due to limited sample sizes for other populations.

Logistic regression, assuming an additive genetic effect and conditioning on the first 10 genetic principal components, was performed using PLINK^27^. We included biallelic autosomal variants with minor allele frequency≥1%, in Hardy-Weinberg Equilibrium (*p*>10^−50^), missingness<5% and imputation quality>0.8. Linkage disequilibrium score regression (LDSC)^28^ intercept was used to assess genomic inflation, and genomic control was applied when the intercept≥1.05. GWAS were only performed where there were≥50 cases.

### Meta-analysis with publicly available GWAS

We meta-analysed our UK Biobank GWAS with previously published GWAS of fibrotic diseases available in the GWAS Catalog^29^ (accessed August 2023). We used search terms ‘fibrosis’, ‘fibrotic’ and the exact names of diseases in our consensus list (**Supplementary Table 1**). Where genome-wide summary statistics were not publicly available, we contacted authors. Where two studies had included the same cohorts, we excluded the smaller study to minimise sample overlap. Ancestry assignment was based on what was reported in the study. A list of studies identified and how they were selected for inclusion or exclusion can be found in **Supplementary Methods, Supplementary Figure 1 and Supplementary Table 2**.

Meta-analyses were performed using a fixed-effect, inverse variance-weighted model in METAL^30^. LDSC intercept was calculated to assess genomic inflation. Variants reaching genome-wide significance (*p*<5×10^−8^) in the European meta-analysis were deemed to be significant. Conditional analyses were performed using GCTA-COJO (Genome-wide Complex Trait Analysis-Conditional and Joint analyses)^31^ to identify independent variants.

### Variants associated with fibrosis across multiple organs

To assess shared genetic loci for fibrosis across different organs, we investigated whether any of the genome-wide significant variants also showed a suggestive association (after Bonferroni correction) with fibrosis in a second organ. For variants showing a suggestive association in multiple organs, colocalisation analyses were performed using coloc^32^ across all 12 organs to identify genetic loci with shared causal variants for fibrosis. We used a threshold of ≥80% probability of shared causal genetic variant (H_4_) to determine organs that colocalise (**Supplementary Methods**).

For association signals that colocalised across ≥3 organs, we tested the association of the sentinel variant with individual fibrotic diseases in UK Biobank defining cases and controls using electronic health records.

### Novel associations and replication analyses

We defined novel associations as variants not previously reported as associated (*p*<5×10^−8^) with any fibrotic disease in GWAS Catalog. To ensure associations were robust, we performed replication analyses for novel variants using the All of Us study^33^ (which included 164,396 European ancestry individuals with genetic data, **Supplementary Methods**). Novel variants were deemed to replicate if significant after Bonferroni correction.

### Functional follow-up

We determined putative causal genes (**Supplementary Methods**) for i) genetic loci that colocalised in ≥3 organs and ii) novel variants that replicated in All of Us. In summary, fine-mapping was conducted following an approximate Bayes factor approach to generate 95% credible sets for each locus, i.e., a set of variants with high probability of containing the causal variant. Variants within credible sets were annotated using VEP (Variant Effect Predictor v115)^34^. Functional effects of variants were predicted using SIFT (Sorting Intolerant From Tolerant)^35^ and CADD (Combined Annotation Dependent Depletion)^36^. Regulatory potential was evaluated using RegulomeDB^37^.

Genetic loci were tested for colocalisation with gene expression (eQTL) and alternative splicing (sQTL) across 54 tissues using GTEx (Genotype-Tissue Expression v8)^38^ and with protein levels (pQTL) in whole-blood using UK Biobank. Nearby genes were tested for association with fibrotic diseases through using available data from Mendelian disease studies (Orphanet^39^), rare variant analyses (AstraZeneca PheWAS Portal^40^) and mouse knockout studies (International Mouse Knockout Consortium^41^).

### Genetic correlation

Genome-wide genetic correlations across organs were calculated using LDSC^28^ (**Supplementary Methods**). To determine whether genetic correlation between organs was due to individuals included as cases in multiple organs, we compared genetic and phenotypic correlations using only UK Biobank. To assess whether genetic correlations are explained by shared heritable risk factors, partial genetic correlations were calculated adjusting for genetic determinants of C-reactive protein (CRP) levels (a measure of inflammatory response), BMI and telomere length (**Supplementary Methods**).

### Multi-ancestry analyses

Variants that colocalised in ≥3 organs were tested for their association in the African, East Asian and South Asian meta-analyses (**Supplementary Table 3**). Variants were deemed associated if significant after Bonferroni correction.

### Comparison of ‘always fibrotic’ and ‘broadly fibrotic’ definitions

For the four organs that had both ‘always fibrotic’ and ‘broadly fibrotic’ definitions, we compared the heritability and calculated the genetic correlation between each definition using LDSC.

## Results

### European GWAS meta-analysis

GWAS were performed separately in 12 organ-systems in UK Biobank and meta-analysed with 15 previous studies (**Supplementary Figures 2 and 3**). Between 3,674 and 546,819 individuals were included in each GWAS (**Table 1**).

**Table 1:** Sources of data and sample sizes for genome-wide meta-analyses of European individuals. Supplementary Table 1 shows the names/codes of the diseases defined in UK Biobank.

|  | Broadly fibrotic |  |  |  |  |  | Always fibrotic |  |  |  |  |  |
| --- | --- | --- | --- | --- | --- | --- | --- | --- | --- | --- | --- | --- |
|  | Individual studies |  |  | Sample size of meta-analysis |  |  | Individual studies |  |  | Sample size of meta-analysis |  |  |
|  | Source: disease | Cases | Controls | Cases | Controls | Total | Source: disease | Cases | Controls | Cases | Controls | Total |
| **Biliary** | UK Biobank: Biliary broadly fibrotic | 4,916 | 49,160 | **12,937** | **65,649** | **78,586** | UK Biobank: Biliary always fibrotic | 883 | 8,830 | **883** | **8,830** | **9,713** |
|  | Cordell (2021): Primary biliary cholangitis | 8,021 | 16,489 |  |  |  |  |  |  |  |  |  |
| **Cardiovascular** | UK Biobank: Cardiovascular broadly fibrotic | 33,671 | 336,710 | **33,671** | **336,710** | **370,381** | - | - | - | - | - | - |
| **Diabetes** | UK Biobank: Diabetes broadly fibrotic | 24,907 | 249,070 | **47,324** | **309,010** | **356,334** |  |  |  | - | - | - |
|  | Mansour Aly (2021): Type 2 diabetes | 9,486 | 2,744 |  |  |  | - | - | - |  |  |  |
|  | Bonàs-Guarch (2018): Type 2 diabetes | 12,931 | 57,196 |  |  |  |  |  |  |  |  |  |
| **Intestinal-pancreatic** | UK Biobank: Intestinal-pancreatic broadly fibrotic | 49,710 | 368,292 | **49,994** | **369,227** | **419,221** | - | - | - | - | - | - |
|  | Garcia-Etxebarria (2022): Crohn's disease | 284 | 935 |  |  |  |  |  |  |  |  |  |
| **Liver** | UK Biobank: Liver broadly fibrotic | 4,437 | 44,370 | **7,738** | **72,148** | **79,886** | UK Biobank: Liver always fibrotic | 1,414 | 14,140 | **2,126** | **15,566** | **17,692** |
|  | Buch (2015): Alcohol-related liver cirrhosis | 712 | 1,426 |  |  |  |  |  |  |  |  |  |
|  | Namjou (2019): Non-alcoholic fatty liver disease | 1,106 | 8,571 |  |  |  | Buch (2015): Alcohol-related liver cirrhosis | 712 | 1,426 |  |  |  |
|  | Anstee (2020): Non-alcoholic fatty liver disease | 1,483 | 17,781 |  |  |  |  |  |  |  |  |  |
| **Lymphatic** | UK Biobank: Lymphatic broadly fibrotic | 334 | 3,340 | **334** | **3,340** | **3,674** | - | - | - | - | - | - |
| **Pulmonary** | UK Biobank: Pulmonary broadly fibrotic | 2,598 | 25,980 | **6,723** | **46,444** | **53,167** | UK Biobank: Pulmonary always fibrotic | 1,764 | 17,640 | **5,889** | **38,104** | **43,993** |
|  | Allen (2022): Idiopathic pulmonary fibrosis | 4,125 | 20,464 |  |  |  | Allen (2022): Idiopathic pulmonary fibrosis | 4,125 | 20,464 |  |  |  |
| **Reproductive** | UK Biobank: Reproductive broadly fibrotic | 7,813 | 78,130 | **7,813** | **78,130** | **85,943** | - | - | - | - | - | - |
| **Skeletal** | UK Biobank: Skeletal broadly fibrotic | 6,516 | 65,160 | **28,866** | **139,983** | **168,849** | - | - | - | - | - | - |
|  | Ishigaki (2022): Rheumatoid arthritis | 22,350 | 74,823 |  |  |  |  |  |  |  |  |  |
| **Skin** | UK Biobank: Skin broadly fibrotic | 6,593 | 65,930 | **20,285** | **104,863** | **125,148** | UK Biobank: Skin always fibrotic | 6,126 | 61,260 | **13,710** | **89,603** | **103,313** |
|  | Riesmeijer (2024): Dupuytren's disease | 7,584 | 28,343 |  |  |  |  |  |  |  |  |  |
|  | Julia (2018): Systemic lupus erythematosus | 907 | 1,524 |  |  |  | Riesmeijer (2024): Dupuytren's disease | 7,584 | 28,343 |  |  |  |
|  | Bentham (2015): Systemic lupus erythematosus | 5,201 | 9,066 |  |  |  |  |  |  |  |  |  |
| **Systemic** | UK Biobank: Systemic broadly fibrotic | 1,620 | 16,200 | **11,300** | **35,330** | **46,630** | - | - | - | - | - | - |
|  | López-Isac (2019): Systemic sclerosis | 9,095 | 17,584 |  |  |  |  |  |  |  |  |  |
|  | Taylor (2017): Sjögren's syndrome | 585 | 1,546 |  |  |  |  |  |  |  |  |  |
| **Urinary** | UK Biobank: Urinary broadly fibrotic | 6,011 | 60,110 | **47,406** | **499,413** | **546,819** | - | - | - | - | - | - |
|  | Wuttke (2019): Chronic kidney disease | 41,395 | 439,303 |  |  |  |  |  |  |  |  |  |

### Variants associated with fibrosis across multiple organs

When using the ‘always fibrotic’ case definition, no genetic variant reaching genome-wide significance in one organ was also associated with fibrosis in another organ. When using the ‘broadly fibrotic’ definitions, 42 independent signals reached genome-wide significance in one organ and also showed a suggestive association in at least one further organ. Twenty-nine signals colocalised across ≥2 organs (**Supplementary Table 4**). Eight genetic loci colocalised across ≥3 organs and were investigated further to identify putative causal genes (**Table 2, Supplementary Table 5** and **Supplementary Figure 4**).

**Table 2:** Summary of association and functional analyses for loci that colocalised across three or more organs. Sentinel variants for eight signals that colocalised across three or more organ-systems in the European ‘broadly fibrotic’ GWAS. The effect sizes and *p* values are given for the organs where the signals colocalised. Chr=Chromosome. Position in build 37. Ref allele=reference allele (i.e., the non-effect allele). EAF=Effect allele frequency. QTLs=Quantitative trait loci. The QTL column denotes eQTLs (gene expression QTLs), pQTLs (protein level QTLs) and sQTLs (splicing QTLs) that colocalise (H_4_≥80%) with the fibrosis association, i.e., where there is evidence that there is a shared genetic determinant of fibrosis and gene expression, protein levels or splicing of a particular gene and in which tissue the QTL was observed in (pQTL analyses only performed using whole blood). Full results including fine-mapping and bioinformatic analyses of epigenetic markers, mouse knockout studies, lookup of rare Mendelian diseases and rare variant analyses are in **Supplementary Table 5**. ^a^ Despite this variant not showing an association, other variants in LD were associated and the loci colocalised.

| Chr | Position | rsid | Ref allele | Effect allele | EAF | Organs colocalised | Organ | OR [95% CI] | *p* | Annotation | QTLs |
| --- | --- | --- | --- | --- | --- | --- | --- | --- | --- | --- | --- |
| 1 | 67820194 | rs6679356 | T | C | 17.2% | 3 | Biliary<br>Skeletal<br>Skin | 1.28 [1.23, 1.33]<br>1.07 [1.04, 1.10]<br>1.08 [1.04, 1.12] | $2.71 \times 10^{-38}$<br>$1.46 \times 10^{-6}$<br>$5.27 \times 10^{-6}$ | *IL12RB2*<br>(intron) | eQTL: *IL12RB2* (colon, lung) |
| 1 | 114377568 | rs2476601 | G | A | 10.3% | 4 | Cardiovascular<br>Diabetes<br>Skeletal<br>Skin | 1.07 [1.04, 1.10]<br>1.07 [1.05, 1.10]<br>1.56 [1.51, 1.61]<br>1.10 [1.05, 1.14] | $6.51 \times 10^{-7}$<br>$9.59 \times 10^{-8}$<br>$8.72 \times 10^{-158}$<br>$8.23 \times 10^{-6}$ | *PTPN22*<br>(missense) | sQTL: *TRIM33* (thyroid) |
| 2 | 27742603 | rs780093 | T | C | 61.2% | 3 | Diabetes<br>Liver<br>Urinary | 1.07 [1.05, 1.09]<br>0.91 [0.88, 0.94]<br>1.04 [1.03, 1.06] | $7.15 \times 10^{-16}$<br>$7.18 \times 10^{-8}$<br>$5.21 \times 10^{-7}$ | *GCKR*<br>(intron) | eQTL: *TRIM54* (brain), *GTF3C2* (brain), *PPM1G* (pancreas)<br>sQTL: *SNX17* (adipose), *PPM1G* (adrenal gland), *GTF3C2* (nerve) |
| 7 | 128701331 | rs13246321 | T | C | 10.9% | 4 | Biliary<br>Skeletal<br>Skin<br>Systemic | 1.26 [1.20, 1.30]<br>1.15 [1.11, 1.18]<br>1.17 [1.13, 1.22]<br>1.44 [1.35, 1.54] | $1.36 \times 10^{-22}$<br>$2.74 \times 10^{-16}$<br>$2.54 \times 10^{-15}$<br>$3.83 \times 10^{-30}$ | *TPI1P2/TNPO3*<br>(Downstream) | - |
| 8 | 11343973 | rs2736340 | C | T | 25.7% | 3 | Skeletal<br>Skin<br>Systemic | 1.09 [1.06, 1.11]<br>1.10 [1.07, 1.14]<br>1.21 [1.16, 1.26] | $3.97 \times 10^{-11}$<br>$8.08 \times 10^{-11}$<br>$1.21 \times 10^{-22}$ | *BLK*<br>(Upstream) | eQTL: *BLK* (adipose, breast, lymphocytes, nerve), *FAM167A* (cultured fibroblasts), *RP11-148O21.4* (lymphocytes), *RP11-148O21.2* (lymphocytes) |
| 12 | 111865049 | rs7310615 | G | C | 48.2% | 5 | Cardiovascular<br>Diabetes<br>Pulmonary<br>Skeletal<br>Urinary | 1.07 [1.06, 1.09]<br>1.04 [1.02, 1.05]<br>1.07 [1.03, 1.12]<br>1.08 [1.06, 1.11]<br>1.04 [1.02, 1.06] | $9.20 \times 10^{-18}$<br>$8.54 \times 10^{-6}$<br>0.0004<br>$4.19 \times 10^{-12}$<br>$1.38 \times 10^{-5}$ | *SH2B3*<br>(intron) | pQTL: *BRAP* (blood), *ERP29* (blood), *PPP1CC* (blood) |
| 15 | 38834033 | rs8032939 | T | C | 24.5% | 4 | Biliary<br>Cardiovascular<br>Diabetes<br>Skeletal | 1.07 [1.03, 1.11]<br>1.04 [1.02, 1.06]<br>1.05 [1.03, 1.07]<br>1.09 [1.06, 1.12] | $9.30 \times 10^{-5}$<br>$1.14 \times 10^{-5}$<br>$1.84 \times 10^{-8}$<br>$3.42 \times 10^{-12}$ | *RASGRP1*<br>(intron) | eQTL: *RASGRP1* (spleen, testis), *ENSG00000259345.5* (stomach) |
| 22 | 21979096 | rs11089637 | T | C | 14.8% | 3 | Skeletal<br>Skin<br>Systemic | 1.09 [1.06, 1.13]<br>1.08 [1.05, 1.12]<br>1.07 [0.98, 1.17] | $2.22 \times 10^{-9}$<br>$7.20 \times 10^{-6}$<br>0.121 <sup>a</sup> | *UBE2L3*<br>(downstream/intron) | eQTL: *UBE2L3* (8 tissues), *CCDC116* (22 tissues), *YDJC* (brain, colon), *ENSG00000273342.1* (Artery)<br>sQTL: *UBE2L3* (testis) |

There were two genetic loci that colocalised across ≥3 organs on chromosome 1. The first signal (rs6679356), which colocalised for biliary, skin and skeletal fibrosis, was located in *IL12RB2* and was an eQTL for *IL12RB2* in colon and lung tissue. The second signal colocalised across 4 organs (cardiovascular, diabetes, skeletal and skin) with skeletal fibrosis showing the strongest association (OR=1.56, 95% CI [1.51, 1.61], *p*=8.72×10^−158^). A variant in the credible set (rs2476601) had a very high individual posterior inclusion probability of being causal (PIP=0.752) and is a missense variant in *PTPN22*.

Of the signals that colocalised across ≥3 organs, all but one had consistent direction of effects across organs, i.e., alleles associated with increased risk of fibrosis in one organ were also associated with increased risk of fibrosis in the other organs. The only signal with inconsistent direction of effect was on chromosome 2. For variant rs780093, allele C was associated with increased risk of diabetes (OR=1.07, 95% CI [1.05, 1.09], *p*=7.15×10^−16^) and urinary fibrosis (OR=1.04, 95% CI [1.03, 1.06], *p*=5.21×10^−7^), but decreased risk of liver fibrosis (OR=0.91, 95% CI [0.88, 0.94], *p*=7.18×10^−8^). The credible set for this signal included intronic variants and a missense variant of *GCKR* (though this variant had a low individual posterior inclusion probability of being causal (PIP=0.065).

The signal on chromosome 7, located near *TPI1P2* and *TNPO3*, was strongly associated with biliary, skeletal, skin and systemic fibrosis. The signal on chromosome 8 contained only one variant in the credible set, rs2736340, which is upstream of *BLK*. The signal colocalised with expression of *BLK* in five tissues, *FAM167A* in fibroblasts and RP11-148O21.4 and RP11-148O21.2 in lymphocytes.

The association signal on chromosome 12 colocalised (H_4_≥80%) across five organs (**Figure 2**) with two additional organs (biliary and systemic fibrosis) also showing some evidence of colocalisation (H_4_≥70%, **Supplementary Figure 4**). The strongest associations for the signal were observed for variant rs7310615 for cardiovascular (OR=1.07, 95% CI [1.06, 1.09], *p*=9.20×10^−18^) and skeletal fibrosis (OR=1.08, 95% CI [1.06, 1.11], *p*=4.19×10^−12^). Through fine-mapping of the GWAS results of cardiovascular (the organ with the strongest association for the signal), there were seven variants in the 95% credible set. Of these, two were located in the *SH2B3* gene (including a missense variant) and the other five were intronic to *ATXN2*. The association signal colocalised with pQTL associations of BRAP, ERP29 and PPP1CC in whole blood. Of the nearby genes, rare variants in *SH2B3, ATXN2, CUX2* and *TMEM116* show an association in UK Biobank with diseases included in our list of ‘broadly fibrotic’ diseases (**Supplementary Table 5**). Variant rs7310615 was tested for association with individual fibrotic diseases. In UK Biobank, rs7310615 showed associations with ICD-10 codes I25.1 (atherosclerotic heart disease, OR=1.08, 95% CI [1.06, 1.10], *p*=6.60×10^−7^), I35.0 (aortic stenosis, OR=1.10, 95% CI [1.04, 1.16], *p*=0.0005), I80.2 (phlebitis, OR=1.07, 95% CI [1.02, 1.12], *p*=0.004), M35.0 (sicca syndrome, OR=1.20, 95% CI [1.08, 1.34], *p*=0.001), J60 (coal worker pneumoconiosis, OR=2.30, 95% CI [1.33, 3.96], *p*=0.003), J84.9 (interstitial pulmonary disease, OR=1.22, 95% CI [1.07, 1.38], *p*=0.002), K80.4 (calculus of bile duct, OR=0.81, 95% CI [0.71, 0.94], *p*=0.005), multiple type 1 diabetes-related ICD10 codes and multiple rheumatoid arthritis-related ICD-10 codes (**Supplementary Figure 5**).

**Figure 2:**
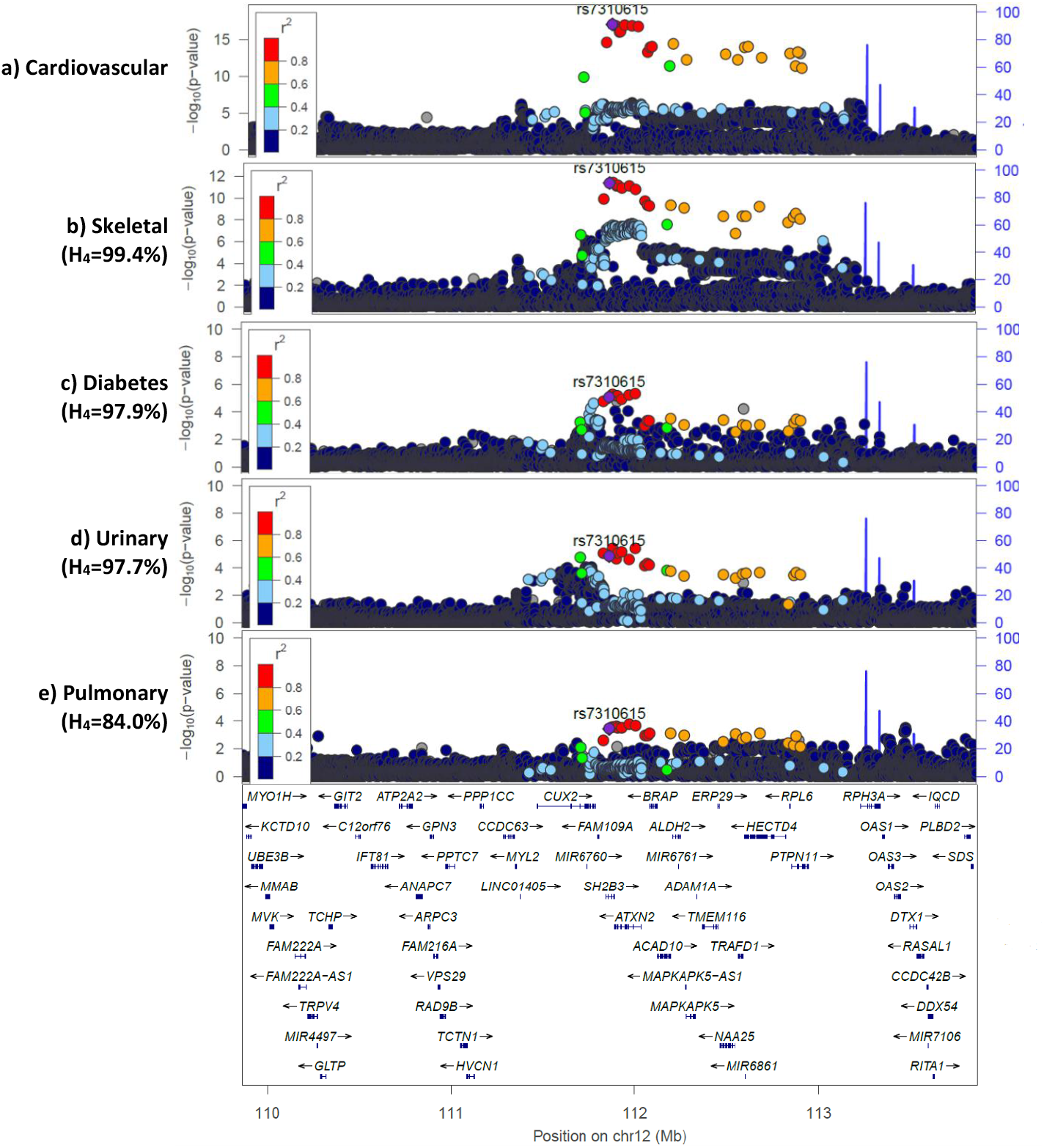
Region plots for the locus that showed evidence of colocalisation across five organs. Each point represents a variant with chromosomal position (GRCh37) on the x axis and −log_10_(*p* value) on the y axis. Variant rs7310615 is coloured purple with other variants coloured by strength of linkage disequilibrium (LD) with rs7310615 based on the 1000 genomes EUR population. Recombination rates are shown by the blue lines and positions of genes are shown at the bottom. H_4_ is the probability of a shared causal variant between that organ and cardiovascular fibrosis. Organs with H_4_≥80% were deemed to have colocalised. Plots created using LocusZoom^42^.

**Figure 3:**
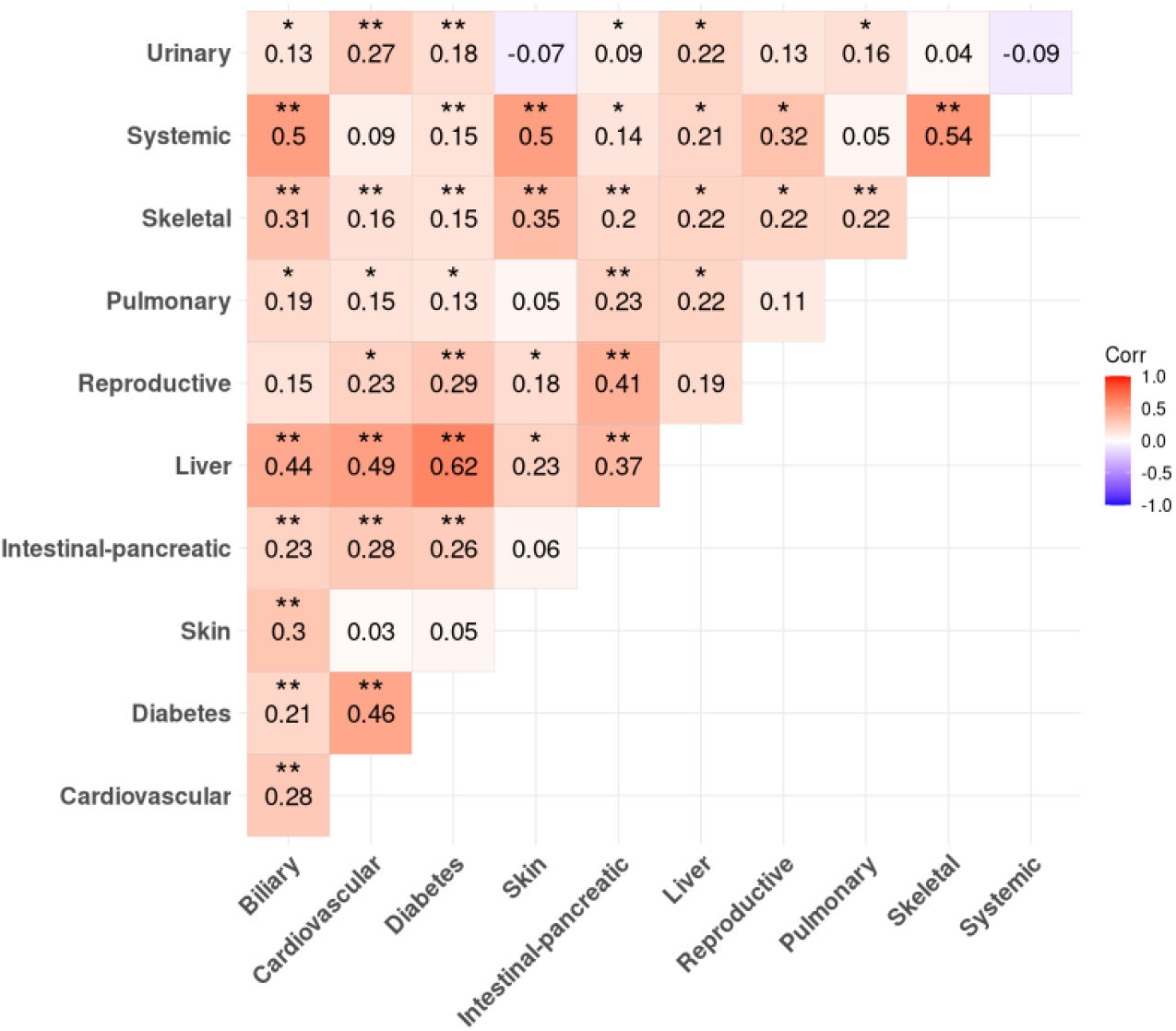
Heatmap of genetic correlations for ‘broadly fibrotic’. Genome-wide genetic correlations were calculated with LDSC using GWAS summary statistics from the European ‘broadly fibrotic’ analyses. Genetic correlations are shown and colours represent the strength of the correlation. ** Denotes correlations significant after Bonferroni correction (*p*<9.09×10^−4^). * Denotes correlations with *p*<0.05.

The signal on chromosome 15 was intronic to *RASGRP1* and colocalised with expression of *RASGRP1* and ENSG00000259345.5. Variants in *RASGRP1* were associated with autoimmune lymphoproliferative syndrome (ALPS) in Orphanet, which may manifest glomerulonephritis, renal insufficiency, pulmonary infiltrates, pulmonary fibrosis, systemic lupus erythematosus, as well as skin neoplasm (diseases we consider fibrotic). The final association signal on chromosome 22 colocalised across skeletal, systemic and skin fibrosis and was located near *UBE2L3*.

Sensitivity analyses testing the signals that colocalised across multiple organs with individual fibrotic diseases can be found in **Supplementary Figures 5-12**.

### Novel associations with fibrosis

Of the 417 genome-wide significant associations with fibrosis in at least one organ, 35 were not reported as associated with a fibrotic disease in GWAS Catalog. Of these, two replicated in the All of Us study (**Supplementary Table 6**).

The first novel replicated variant, rs113477191, was associated with intestinal-pancreatic fibrosis (OR=1.13, 95% CI [1.08, 1.19], *p*=3.67×10^−8^, effect allele frequency (EAF)=2.1%, **Supplementary Figure 13**). rs113477191 is a predicted “deleterious” missense variant in *FAM180A*. Although not previously reported in GWAS Catalog, rare variants in *FAM180A* showed an association with diverticular disease of the intestine (a ‘broadly fibrotic’ intestinal-pancreatic disease) in UK Biobank (**Supplementary Table 5**).

The second novel replicated signal, with sentinel variant rs2580350, was associated with urinary fibrosis (OR=1.06, 95% CI [1.04, 1.07], *p*=1.45×10^−9^, EAF=55.3%, **Supplementary Figure 13**). This signal was intronic to *TFCP2L1* and colocalised with alternative splicing of *TFCP2L1* in 10 tissues (**Supplementary Table 5)**.

### Genetic correlation

Of 55 pairs of ‘broadly fibrotic’ diseases, 27 showed significant positive genetic correlation after Bonferroni correction. Strong genetic correlations were observed for diabetes-liver (r^2^=0.62), skeletal-systemic (r^2^=0.54), systemic-skin (r^2^=0.50), and systemic-biliary (r^2^=0.50). Biliary, cardiovascular, diabetes, intestinal-pancreatic and skeletal fibrosis all had significant positive genetic correlations with ≥7 other organ-systems. For ‘always fibrotic’, no correlations were significant (**Supplementary Figure 14**). When restricting to only UK Biobank, we observed even higher genetic correlations despite little phenotypic overlap (**Supplementary Figures 15-17**). Genetic correlation estimates remained largely unchanged when adjusting for genetics of BMI, CRP and telomere length (**Supplementary Figure 18**).

### Multi-ancestry analyses

Sample sizes for non-European analyses were generally small, apart from diabetes (n=77,569) and skeletal fibrosis (n=11,025) in East Asian ancestry. Of the eight variants that colocalised across ≥3 organs in European analyses, three showed a significant association with fibrosis in one organ in one non-European ancestry group after Bonferroni correction (*p*<6.85×10^−4^, **Supplementary Figure 19**). In each instance, the direction of effect was the same as observed in Europeans.

### Comparison of ‘always fibrotic’ and ‘broadly fibrotic’ definitions

There was high genetic correlation between the two case definitions for the four organs included in both analyses. Heritability estimates were slightly higher for the pulmonary, biliary and skin ‘always fibrotic’ definition (**Supplementary Table 7**).

## Discussion

We have identified eight genetic loci which demonstrated shared genetic determinants of fibrosis across three or more organs, highlighting cross-organ genetic architecture which may underlie fibrotic diseases.

The signal that colocalised across five organs implicates the *SH2B3* gene, which plays a key role in cellular processes such as haematopoiesis, cell migration and signalling of growth factors^43^. *SH2B3* has been previously reported as associated with cardiac inflammation and fibrosis^44, 45^.

The strongest genetic correlation was between diabetes and liver fibrosis and it is estimated that globally one-in-five individuals with type 2 diabetes mellitus develop liver fibrosis^46^. However, from genetic correlations, we cannot determine whether there are shared genetic determinants or whether one disease causes the other (for example, if diabetes causes liver fibrosis). By comparing genetic and phenotypic correlation in UK Biobank, we showed that the high genetic correlations were not likely due to sample overlap. Partial genetic correlation estimates suggest genetic correlation between organs remained broadly unchanged following adjustment for the genetics of some shared heritable risk factors (external factors that cause inflammation, BMI and telomere length). There may be other shared heritable risk factors that explain the high genetic correlations that we have not investigated.

There is no set definition, nor hallmarks, of what diseases should be defined as fibrotic^24^. We based our definition on a published survey; however, even within this survey of expert opinion, there was still disagreement^24^. Therefore, we used two different definitions: a narrow definition where the majority of respondents agreed that diseases were always fibrotic (such as idiopathic pulmonary fibrosis and liver cirrhosis), and a broader definition including diseases where fibrosis may not occur in all individuals or all stages of disease (such as Crohn’s disease or diabetes). For the four organ-systems included in both the ‘always fibrotic’ and ‘broadly fibrotic’ analyses, we observed high genetic correlations between disease definitions. Although LDSC accounts for sample overlap, ‘always fibrotic’ cases were fully nested within the ‘broadly fibrotic’ case definition. The two case definitions for biliary fibrosis had limited overlap (12,937 vs 883 cases) and the ‘always fibrotic’ definition led to far fewer significantly associated loci. However, we still observed a high genetic correlation. This suggests, at least for biliary fibrosis, that using a broader case definition may increase statistical power while retaining underlying genetic architecture for fibrosis.

A limitation of this study is the use of electronic health records (EHR) for defining cases in UK Biobank, where the source of EHR has been shown to affect GWAS results^47^. Some individuals may be misdiagnosed or have suspected diseases recorded in their EHR before a final disease diagnosis is made. However, misdiagnosed codes will likely fall within the same organ. We therefore decided to group diseases by organ to minimise misclassification, reduce multiple testing and maximise statistical power. This makes the assumption that all fibrotic diseases within one organ are driven by the same genetic determinants, which may not be true. For associations identified, we were able to investigate individual fibrotic diseases to determine whether associations are observed for all fibrotic diseases in an organ or driven by one disease in particular.

Secondly, we used the European analyses as our primary discovery analysis which may mean we miss associations that are present in non-European populations, which may also limit generalisability of results. Despite replicating some findings in non-European populations, due to limited sample sizes our effect size estimates were imprecise for most diseases, supporting the need for more diverse studies.

Finally, although we identified shared genetic variants, we cannot confirm whether these genetic variants are involved in the development of fibrosis itself. Furthermore, shared genetic loci were predominantly identified using ‘broadly fibrotic’ case definitions. This may be due to more organs and individuals being included and thereby having larger statistical power, or could be due to these genetic loci not being directly involved in fibrosis. Regardless, we identified shared genetic loci across fibrosis-related diseases.

In summary, we found evidence of shared genetic determinants of fibrotic diseases, both at the genome-wide level and individual genetic loci. Identifying shared biological mechanisms and therapeutic targets that target these mechanisms has the potential to improve patient outcomes over a wide range of diseases. Studying fibrosis across diseases and organs can enable shared learning of mechanism and also the ability to define patient groups likely to be benefit from a new therapy at an early opportunity.

## Data Availability

Access to UK Biobank (https://www.ukbiobank.ac.uk/) and All of Us Research Programme (https://www.researchallofus.org/register/) individual-level data are available to approved researchers upon application or data access request. Genome-wide summary statistics from the single organ analyses will be made publicly available via the EMBL-EBI GWAS Catalog.

## Ethics

UK Biobank genetic and phenotypic data were obtained under UK Biobank application 77050. UK Biobank has ethical approval from the UK National Health Service (NHS) National Research Ethics Service (11/NW/0382). This research also used the All of Us Research Programme data, which can be accessed by registered users from institutions that have a Data Use and Registration Agreement (DURA) in place. All methods were carried out in accordance with the All of Us Data User Code of Conduct, which ensures participant privacy, data security, and respectful use of data. The All of Us Research Program gets ethical approval and regulatory oversight primarily from its centralised All of Us Institutional Review Board (IRB) which operates in accordance with regulations and guidance from the Office for Human Research Protections (OHRP).

## Acknowledgements

We thank all volunteers participating in the UK Biobank and All of Us studies. This research has been conducted using the UK Biobank Resource under Application Number 77050. The study also used resources in the All of Us Research Programme. We also thank authors of studies that have shared GWAS summary statistics for use in this study. This research used the ALICE and SPECTRE High Performance Computing Facility at the University of Leicester.

## Author Contributions

Conceptualisation: LVW, RGJ, RJA, JKQ and HJL; Methodology: LVW, RJA, EJ and OCL; Analysis: EJ, RJA, TH-B, JP, RN, HJP, RW and FA; Data Curation: EJ, GMM and GP; Writing – Original Draft: RJA and EJ; Writing – Review & Editing: all authors; Supervision: RJA, LVW and JKQ; Funding Acquisition: RGJ, LVW, JKQ, RJA and HJL.

## On Behalf of the DEMISTIFI Consortium

Andrew Thorley, Anna Duckworth, Ali-Reza Mohammadi-Nejad, Aloysious Aravinthan, Anthony Harbottle, Armando Mendez Villalon, Chris Scotton, Christopher Denton, Daniel Lea, Dorothee Auer, Ebrima Joof, Eleanor Cox, Elizabeth Eves, Elizabeth Robertson, Emma Blamont, Fasihul Khan, Georgie Massen, Gina Parcesepe, Gisli Jenkins, Gordon Moran, Guruprasad Aithal, Hilary Longhurst, Iain Stewart, Jane Paxton, Jennifer Quint, Karen Piper Hanley, Kate Frost, Leo Casmino, Lisa Chakrabarti, Louise Wain, Margot Roeth, Maria Kaisar, Martin Craig, Michael Nation, Mohammad Alireza Kisomi, Mujdat Zeybel, Neil Guha, Nicholas Selby, Nick Oliver, Nick Selby, Olivia C Leavy, Penny Gowland, Philip Quinlan, Rachel Chambers, Richard Allen, Richard Hubbard, Rob Slack, Rutger Ploeg, Sam Moss, Sara Fawaz, Scott Turner, Shauntelle Quammie, Simon Johnson, Stamatios N Sotiropoulos, Stuart Astbury, Susan Francis, Tom Giles, Valerie Quinn, Wendy Adams, Xin Chen, and Zhendi Gong

## References

1. Hao, M. et al. The pathogenesis of organ fibrosis: Focus on necroptosis. Br. J. Pharmacol. 180, 2862–2879 (2023).

2. Mutsaers, H. A., Merrild, C., Nørregaard, R. & Plana-Ripoll, O. The impact of fibrotic diseases on global mortality from 1990 to 2019. Journal of Translational Medicine 21, 818 (2023).

3. Massen, G. M. et al. Using routinely collected electronic healthcare record data to investigate fibrotic multimorbidity in England. Clinical epidemiology, 433–443 (2024).

4. Lurje, I., Gaisa, N. T., Weiskirchen, R. & Tacke, F. Mechanisms of organ fibrosis: Emerging concepts and implications for novel treatment strategies. Mol. Aspects Med. 92, 101191 (2023).

5. Wynn, T. A. & Ramalingam, T. R. Mechanisms of fibrosis: therapeutic translation for fibrotic disease. Nat. Med. 18, 1028–1040 (2012).

6. Li, Z. I. et al. C-reactive protein promotes acute renal inflammation and fibrosis in unilateral ureteral obstructive nephropathy in mice. Laboratory investigation 91, 837–851 (2011).

7. Zhang, R. et al. C-reactive protein promotes cardiac fibrosis and inflammation in angiotensin II– induced hypertensive cardiac disease. Hypertension 55, 953–960 (2010).

8. Wu, Y., Zheng, G., Zhang, F. & Li, W. Association of high-sensitivity C-reactive protein with hepatic fibrosis in patients with metabolic dysfunction-associated steatotic liver disease. Frontiers in Immunology 16, 1544917 (2025).

9. Zhang, K., Li, A., Zhou, J., Zhang, C. & Chen, M. Genetic association of circulating C-reactive protein levels with idiopathic pulmonary fibrosis: a two-sample Mendelian randomization study. Respiratory Research 24, 7 (2023).

10. Gopalakrishna, H., Fashanu, O. E., Nair, G. B. & Ravendhran, N. Association between body mass index and liver stiffness measurement using transient elastography in patients with non-alcoholic fatty liver disease in a hepatology clinic: a cross sectional study. Translational gastroenterology and hepatology 8, 10 (2023).

11. Kalson, N. S. et al. Reduced telomere length is associated with fibrotic joint disease suggesting that impaired telomere repair contributes to joint fibrosis. PLoS One 13, e0190120 (2018).

12. Duckworth, A. et al. Telomere length and risk of idiopathic pulmonary fibrosis and chronic obstructive pulmonary disease: a mendelian randomisation study. The Lancet Respiratory Medicine 9, 285–294 (2021).

13. Cordell, H. J. et al. International genome-wide meta-analysis identifies new primary biliary cirrhosis risk loci and targetable pathogenic pathways. Nature communications 6, 8019 (2015).

14. Sakaue, S. et al. A cross-population atlas of genetic associations for 220 human phenotypes. Nat. Genet. 53, 1415–1424 (2021).

15. Allen, R. J. et al. Genome-wide association study across five cohorts identifies five novel loci associated with idiopathic pulmonary fibrosis. Thorax (2022).

16. Buch, S. et al. A genome-wide association study confirms PNPLA3 and identifies TM6SF2 and MBOAT7 as risk loci for alcohol-related cirrhosis. Nat. Genet. 47, 1443–1448 (2015).

17. Ishigaki, K. et al. Multi-ancestry genome-wide association analyses identify novel genetic mechanisms in rheumatoid arthritis. Nat. Genet. 54, 1640–1651 (2022).

18. Riesmeijer, S. A. et al. A genome-wide association meta-analysis implicates Hedgehog and Notch signaling in Dupuytren’s disease. Nature Communications 15, 199 (2024).

19. Bentham, J. et al. Genetic association analyses implicate aberrant regulation of innate and adaptive immunity genes in the pathogenesis of systemic lupus erythematosus. Nat. Genet. 47, 1457–1464 (2015).

20. López-Isac, E. et al. GWAS for systemic sclerosis identifies multiple risk loci and highlights fibrotic and vasculopathy pathways. Nature communications 10, 4955 (2019).

21. Garcia-Etxebarria, K. et al. Local genetic variation of inflammatory bowel disease in Basque population and its effect in risk prediction. Scientific Reports 12, 3386 (2022).

22. Wuttke, M. et al. A catalog of genetic loci associated with kidney function from analyses of a million individuals. Nat. Genet. 51, 957–972 (2019).

23. Nauffal, V. et al. Noninvasive assessment of organ-specific and shared pathways in multi-organ fibrosis using T1 mapping. Nat. Med. 30, 1749–1760 (2024).

24. Massen, G. M. et al. Classifying the unclassifiable—a Delphi study to reach consensus on the fibrotic nature of diseases. QJM: An International Journal of Medicine 116, 429–435 (2023).

25. Bycroft, C. et al. The UK Biobank resource with deep phenotyping and genomic data. Nature 562, 203–209 (2018).

26. Shrine, N. et al. New genetic signals for lung function highlight pathways and chronic obstructive pulmonary disease associations across multiple ancestries. Nat Genet 51, 481–493 (2019).

27. Purcell, S. et al. PLINK: A Tool Set for Whole-Genome Association and Population-Based Linkage Analyses. Am J Hum Genet 81, 559–575 (2007).

28. Bulik-Sullivan, B. K. et al. LD Score regression distinguishes confounding from polygenicity in genome-wide association studies. Nat. Genet. 47, 291–295 (2015).

29. Buniello, A. et al. The NHGRI-EBI GWAS Catalog of published genome-wide association studies, targeted arrays and summary statistics 2019. Nucleic Acids Res. 47, D1005–D1012 (2019).

30. Willer, C. J., Li, Y. & Abecasis, G. R. METAL: fast and efficient meta-analysis of genomewide association scans. Bioinformatics 26, 2190–2191 (2010).

31. Yang, J., Lee, S. H., Goddard, M. E. & Visscher, P. M. GCTA: a tool for genome-wide complex trait analysis. The American Journal of Human Genetics 88, 76–82 (2011).

32. Giambartolomei, C. et al. Bayesian test for colocalisation between pairs of genetic association studies using summary statistics. PLoS genetics 10, e1004383 (2014).

33. All of Us Research Program Investigators. The “All of Us” research program. N. Engl. J. Med. 381, 668–676 (2019).

34. McLaren, W. et al. The ensembl variant effect predictor. Genome Biol. 17, 122 (2016).

35. Ng, P. C. & Henikoff, S. SIFT: Predicting amino acid changes that affect protein function. Nucleic Acids Res. 31, 3812–3814 (2003).

36. Rentzsch, P., Witten, D., Cooper, G. M., Shendure, J. & Kircher, M. CADD: predicting the deleteriousness of variants throughout the human genome. Nucleic Acids Res. 47, D886–D894 (2019).

37. Dong, S. et al. Annotating and prioritizing human non-coding variants with RegulomeDB v. 2. Nat. Genet. 55, 724–726 (2023).

38. GTEx Consortium. The GTEx Consortium atlas of genetic regulatory effects across human tissues. Science 369, 1318–1330 (2020).

39. Weinreich, S. S., Mangon, R., Sikkens, J. J., Teeuw, M. & Cornel, M. C. Orphanet: a European database for rare diseases. Ned. Tijdschr. Geneeskd. 152, 518–519 (2008).

40. Wang, Q. et al. Rare variant contribution to human disease in 281,104 UK Biobank exomes. Nature 597, 527–532 (2021).

41. Koscielny, G. et al. The International Mouse Phenotyping Consortium Web Portal, a unified point of access for knockout mice and related phenotyping data. Nucleic Acids Res. 42, D802–D809 (2014).

42. Pruim, R. J. et al. LocusZoom: regional visualization of genome-wide association scan results. Bioinformatics 26, 2336–2337 (2010).

43. Stelzer, G. et al. GeneCards – the human gene database. The GeneCards Suite: From Gene Data Mining to Disease Genome Sequence Analysis. Current protocols in bioinformatics, 1.30.1–1.30.33 (2016).

44. Kuo, C. et al. The longevity-associated SH2B3 (LNK) genetic variant: selected aging phenotypes in 379,758 subjects. The Journals of Gerontology: Series A 75, 1656–1662 (2020).

45. Flister, M. J. et al. SH2B3 is a genetic determinant of cardiac inflammation and fibrosis. Circulation: Cardiovascular Genetics 8, 294–304 (2015).

46. Wongtrakul, W., Niltwat, S., Charatcharoenwitthaya, N., Karaketklang, K. & Charatcharoenwitthaya, P. Global prevalence of advanced fibrosis in patients with type 2 diabetes mellitus: a systematic review and meta-analysis. J. Gastroenterol. Hepatol. 39, 2299–2307 (2024).

47. Isgut, M., Song, K., Ehm, M. G., Wang, M. D. & Davitte, J. Effect of case and control definitions on genome-wide association study (GWAS) findings. Genet. Epidemiol. 47, 394–406 (2023).

