## Supplementary Material for "Genome-wide association studies to identify shared and distinct mechanisms of fibrosis across 12 organ-systems"

### Supplementary Methods

#### Selection of previously published GWAS studies

To identify studies to include in our meta-analysis we followed the following steps:

1. Search GWAS Catalog[1] using the terms "fibrosis", "fibrotic" and all of the disease names included in the consensus list from the previous Delphi study
2. We excluded studies that:
    a. Were not GWAS of a fibrotic disease
    b. Included UK Biobank samples to avoid overlap with our UK Biobank analyses
    c. Did not have available genome-wide summary statistics for download. Where these were not downloadable, we contacted study authors directly and excluded studies where we did not get a response
    d. Did not have effect size, standard error and effect allele recorded in the summary statistics
    e. Were related to disease progression rather than disease susceptibility
    f. We also excluded studies where it was not possible to align alleles
3. If any of the remaining studies had overlapping samples, we kept the study with the largest sample size

Studies were then assigned to one of the following ancestry groups based on the ancestry group reported in the original manuscript; European, South Asian, East Asian and African. If the study only included multi-ancestry results or was performed in a different population then these studies were excluded.

Where necessary, chromosomal positions were converted to build 37 using UCSC LiftOver[2] to ensure all studies were on the same build before meta-analysing. To ensure comparability of results across organ-systems we only included variants included in the European analysis.

A summary of all of the papers identified and included/excluded are in **Supplementary Table 2**.

#### Colocalisation analyses

Colocalisation analyses were performed using the coloc v5.2.3 package in R v4.3.1 following the approach proposed by Giambartolomei et al[3]. In summary, the method compares the summary statistics for a region between two traits and calculates the probability of each of the following five hypotheses:

- $H_0$: No association in the region for either trait
- $H_1$: There is an association for the first trait but not the second
- $H_2$: There is an association for the second trait but not the first
- $H_3$: There is an association for both traits but they are with different independent variants
- $H_4$: There is an association for both traits with a shared causal variant

If the posterior probability of $H_4$ (i.e. that there is a shared association between both traits) was greater than or equal to 80%, then we took this as evidence of colocalisation and concluded there was a shared association between the two fibrotic diseases in this region.

#### Replication analyses using All of Us

For replication analyses, we used whole-genome sequencing data from European ancestry individuals in All of Us (AoU), a longitudinal cohort study involving a large and diverse group of participants (over 868,000) in the USA[4]. The Genomic sequencing, detailed quality control (QC) procedures and genetic ancestry inference carried out by AoU are described elsewhere[5]. We removed samples that failed QC (samples included in the AoU flagged sample list) and samples that had up to 2nd degree relatedness (one of each related pair was removed). We included biallelic autosomal variants with minor allele frequency ≥1%, in Hardy-Weinberg Equilibrium ($p>10^{-50}$). For genetic ancestry inference, we used the AoU generated ancestry predictions from a trained classifier using 16 genetic principal components[5].

Cases were defined as individuals with a fibrotic condition recorded in their electronic health records. To exclude cases recorded using suspected conditions we only used the following sources of electronic health records; outpatient visit, office visit, inpatient visit, emergency room visit, emergency room and inpatient visit, laboratory visit, telehealth, non-hospital institution visit, inpatient hospital, outpatient hospital, hospital, ambulatory clinic/centre, emergency room-hospital and urgent care facility. Diseases were recorded using Systematised Nomenclature of Medicine Clinical Terms (SNOMED-CT).

It has been previously reported that disease prevalences are much higher in American electronic health records than expected. Therefore, as has been performed in previous studies[6, 7] to improve case-definitions, we required individuals to have two records of fibrosis in a particular organ to be considered a case. All individuals who were not cases were considered controls and 10 controls per case (matched by age and sex) were selected for the analysis.

Replication analyses were performed for the novel variants from the discovery GWAS that were not previously reported as associated with fibrosis in GWAS Catalog using a logistic regression model adjusting for 10 genetic principal components using PLINK. Variants were considered significant if they were significant after a Bonferroni correction for the number of novel variants investigated further in All of Us.

#### Functional follow-up of signals

Functional analyses were performed to identify putative causal genes. For genetic loci associated with fibrosis across multiple organs, analyses were performed using GWAS results from the organ with the strongest association.

##### *Variant functional annotation*

We annotated the sentinel variants with Ensembl Variant Effect Predictor (VEP) v.115[8], and we identified potentially causal variants using the Wakefield Bayes factor method to calculate the posterior inclusion probability (PIP) of each variant to define a 95% credible set[9]. We performed a comprehensive functional impact assessment using empirical data from several integrated software tools and datasets for the sentinel SNPs and for each variant in the credible set.

To assess the regulatory potential and rank the functional roles of each SNP, we queried RegulomeDB[10], which provides a score ranging from 0 to 1, with 1 being most likely to be a regulatory variant.

We also annotated regions for functionality based on active chromatin marks, including DNase I hypersensitivity hotspots (DHS), open chromatin peaks, transcription factor footprints, enhancer RNAs (eRNAs), Assay for Transposase-Accessible Chromatin using sequencing (ATACseq), ChIP sequencing (CHIPseq), and epigenetic modification of the DNA packaging protein histones (H3K27ac)[11].

***Quantitative Trait Locus analysis***
We interrogated gene expression and splicing Quantitative Trait Locus (eQTLs; sQTLs) from the Genotype-Tissue Expression Project (GTEx v8) (downloaded from https://www.gtexportal.org/, July 2020)[12] to identify whether variants were associated with expression or splicing of any genes across 54 tissues. We accessed UK Biobank (UKBB) summary statistics of 46,836 individuals from the general population with measurements of plasma protein expression QTL. For genes where the sentinel variant showed an association with gene expression, splicing or protein levels, we used coloc[3] R v4.1.3 package to investigate whether the same causal variant was driving both the genetic association with fibrosis and the association with gene expression, protein or splice variant expression levels. As described above, we report shared signals with a colocalisation probability $H_4 \geq 80\%$.

***Rare variant associations***
We looked for exonic rare variant associations (MAF < 1%) with any 'fibrosis' term/disease within ±500 kb of the sentinel variants using both single-variant and gene-based collapsing tests from 281,104 UK Biobank exomes accessed via the AstraZeneca PheWAS portal[13] (https://azphewas.com/). We used a threshold of $p<5x10^{-6}$ for both single-variant and gene-based tests. For the novel replicated variants we only tested associations with 'broadly fibrotic' diseases in that organ. For the genetic associations that colocalised across three or more organs we tested for the association of rare variants with any 'broadly fibrotic' disease. Fibrotic diseases were searched using the mapped ICD10 code in **Supplementary Table 1**.

***Nearby Mendelian disease and mouse knockout orthologs genes***
Finally, we searched for rare Mendelian disease genes using Orphanet[14] (https://www.orpha.net/), with data downloaded in 2021 (http://www.orphadata.org/data/xml/en_product6.xml & http://www.orphadata.org/data/xml/en_product4.xml), and human orthologs of mouse knockout genes from the International Mouse Phenotyping Consortium[15] (https://www.mousephenotype.org/) that were within ±500 kb of each associated sentinel. For the novel replicated variants we only tested associations with 'broadly fibrotic' diseases in that organ. For the genetic associations that colocalised across three or more organs we tested for the association of rare variants with any 'broadly fibrotic' disease. Fibrotic diseases were searched using the name of the disease in **Supplementary Table 1**.

#### Correlation analyses

***Genetic correlation across fibrotic diseases***
Genome-wide genetic correlation for each pair of organs was calculated using bivariate linkage disequilibrium score regression (LDSC) v1.0.0[16]. The 1000 Genomes Project[17] European dataset was used as the linkage disequilibrium (LD) reference panel. Genetic correlations were not calculated using the lymphatic 'broadly fibrotic' case definition due to the low sample size with LDSC requiring sample sizes > 5,000 to converge.

To assess whether the genetic correlations were driven by case overlap between the organs, we calculated pairwise phenotypic correlations (using the “cor” and “cor_pmat” functions in the ggcorrplot package in R v4.3.1) and Jaccard similarity indices (using the “vegdist” function in the vegan package in R v4.3.1.). All heatmaps for the genetic correlation, phenotypic correlation and Jaccard index were generated using the ggcorrplot package in R v4.3.1.

***Partial genetic correlations***

To assess the extent to which the genetics of identified risk factors (body mass index (BMI), C-reactive protein levels  and telomere length) account for the genetic correlation between pairs of fibrotic diseases, we applied the partialLDSC framework (https://gemini-multimorbidity.github.io/partialLDSC/), an extension of the commonly-used LDSC method[18]. PartialLDSC first calculates a covariance matrix across all phenotypes, computes the genetic correlation between the fibrotic disease-pair, then re-estimates the genetic correlation while conditioning on the specified risk factor(s). A t-test is used to determine whether the partial (conditioned) genetic correlation is significantly different to the original estimate. We initially performed univariate partialLDSC, conditioning on one risk factor at a time. If a fibrotic disease-pair had at least two risk factors that each attenuated the genetic correlation (t-test $p < 0.05$), we subsequently conducted multivariate partialLDSC, jointly conditioning on all risk factors with a nominally significant attenuation. Finally, we used ggplot2 in R to generate forest plots illustrating the attenuation of genetic correlations before and after conditioning.

#### Supplementary References

1. Buniello, A. *et al*. The NHGRI-EBI GWAS Catalog of published genome-wide association studies, targeted arrays and summary statistics 2019. *Nucleic Acids Res.* **47**, D1005–D1012 (2019).

2. Hinrichs, A. S. *et al*. The UCSC genome browser database: update 2006. *Nucleic Acids Res.* **34**, D590–D598 (2006).

3. Giambartolomei, C. *et al*. Bayesian test for colocalisation between pairs of genetic association studies using summary statistics. *PLoS genetics* **10**, e1004383 (2014).

4. All of Us Research Program Investigators. The "All of Us" research program. *N. Engl. J. Med.* **381**, 668–676 (2019).

5. Biobank, Mayo Blegen Ashley L. 18 Wirkus Samantha J. 18 Wagner Victoria A. 18 Meyer Jeffrey G. 18 Cicek Mine S. 10 18 & All of Us Research Demonstration Project Teams Choi Seung Hoan 14 http://orcid. org/0000-0002-0322-8970 Wang Xin 14 http://orcid. org/0000-0001-6042-4487 Rosenthal Elisabeth A. 15. Genomic data in the all of us research program. *Nature* **627**, 340–346 (2024).

6. Li, T. *et al*. Long-term medical costs and resource utilization in systemic lupus erythematosus and lupus nephritis: a five-year analysis of a large Medicaid population. *Arthritis Care & Research: Official Journal of the American College of Rheumatology* **61**, 755–763 (2009).

7. Karve, S. *et al*. Healthcare utilization and comorbidity burden among children and young adults in the United States with systemic lupus erythematosus or inflammatory bowel disease. *J. Pediatr.* **161**, 662–670. e2 (2012).

8. Yates, A. *et al*. Ensembl 2016. *Nucleic Acids Res.* **44**, 710 (2016).

9. Wakefield, J. Bayes factors for genome-wide association studies: comparison with P-values. *Genet. Epidemiol.* **33**, 79–86 (2009).

10. Dong, S. *et al*. Annotating and prioritizing human non-coding variants with RegulomeDB v. 2. *Nat. Genet.* **55**, 724–726 (2023).

11. Biddie, S. C., Weykopf, G., Hird, E. F., Friman, E. T. & Bickmore, W. A. DNA-binding factor footprints and enhancer RNAs identify functional non-coding genetic variants. *Genome Biol.* **25**, 208 (2024).

12. Lonsdale, J. *et al*. The genotype-tissue expression (GTEx) project. *Nat. Genet.* **45**, 580–585 (2013).

13. Wang, Q. *et al*. Rare variant contribution to human disease in 281,104 UK Biobank exomes. *Nature* **597**, 527–532 (2021).

14. Weinreich, S. S., Mangon, R., Sikkens, J. J., Teeuw, M. & Cornel, M. C. Orphanet: a European database for rare diseases. *Ned. Tijdschr. Geneeskd.* **152**, 518–519 (2008).

15. Koscielny, G. *et al*. The International Mouse Phenotyping Consortium Web Portal, a unified point of access for knockout mice and related phenotyping data. *Nucleic Acids Res.* **42**, D802–D809 (2014).

16. Bulik-Sullivan, B. K. *et al*. LD Score regression distinguishes confounding from polygenicity in genome-wide association studies. *Nat. Genet.* **47**, 291–295 (2015).

17. 1000 Genomes Project Consortium. A global reference for human genetic variation. *Nature* **526**, 68–74 (2015).

18. Mounier, N. *et al*. Genetics identifies obesity as a shared risk factor for co-occurring multiple long-term conditions. *medRxiv*, 2024.07. 10.24309772 (2024).

### Supplementary Tables

#### Supplementary Table 1: List of diseases and mapped ICD10 and SNOMED codes

See Excel file titled Supplementary_Table_1_List_of_diseases_and_mapped_ICD10_and_SNOMED_codes.xlsx. Summary of columns in the Excel file:

- **Organ**: The organ category under which the disease was grouped.
- **Fibrotic Disease (from Delphi Survey)**: The list of diseases that reached consensus as fibrotic in the Delphi survey and were included in this study.
- **Always Fibrotic Percentage**: Percentage of clinician respondents that agreed the disease is always fibrotic in the Delphi survey.
- **SNOMED Code**: The Systematised Nomenclature of Medicine Clinical Terms (SNOMED-CT) code the disease was mapped to.
- **Term**: The SNOMED description for the disease.
- **ICD10 Code**: The International Classification of Disease 10th Revision (ICD-10) code the disease was mapped to.
- **Description**: The ICD10 description of the disease.

**Supplementary Table 2: Summary of previously published studies of fibrotic diseases and selection of studies for meta-analysis**
Summary of all studies identified during screening and exclusion reason if not included. Studies that were included in the meta-analysis are highlighted in green.

| Organ | Author (year) | DOI | Disease | Always or broadly fibrotic | Ancestry | Cases | Controls | Summary statistics available | Include/ exclude | Reason for exclusion |
|---|---|---|---|---|---|---|---|---|---|---|
| Biliary | Jiang L (2021) | doi.org/10.1038/s41588-021-00954-4 | Cholangitis | Broadly | European | 359 | 455,989 | Yes | Exclude | Includes UK Biobank |
| Biliary | Backman JD (2021) | doi.org/10.1038/s41586-021-04103-z | ICD10 K83.0: Cholangitis | Broadly | European | 750 | 387,180 | Yes | Exclude | Includes UK Biobank |
| Biliary | Backman JD (2021) | doi.org/10.1038/s41586-021-04103-z | ICD10 K83.1: Obstruction of bile duct | Broadly | European | 1,070 | 386,859 | Yes | Exclude | Includes UK Biobank |
| Biliary | Cordell HJ (2015) | doi.org/10.1038/ncomms9019 | Primary biliary cholangitis | Broadly | European | 2,764 | 10,475 | Yes | Exclude | Overlaps with larger study included in analysis |
| Biliary | Cordell HJ (2021) | doi.org/10.1016/j.jhep.2021.04.055 | Primary biliary cholangitis | Broadly | European | 8,021 | 16,489 | Yes | Include | - |
| Biliary | Liu JZ (2012) | doi.org/10.1038/ng.2395 | Primary biliary cirrhosis | Broadly | European | 2,861 | 8,514 | Yes | Exclude | Overlaps with larger study included in analysis |
| Biliary | Nakamura M (2012) | doi.org/10.1016/j.ajhg.2012.08.010 | Primary biliary cholangitis | Broadly | East Asian | 487 | 476 | Yes | Include | - |
| Biliary | Jiang L (2021) | doi.org/10.1038/s41588-021-00954-4 | Primary biliary cirrhosis | Broadly | European | 121 | 456,227 | Yes | Exclude | Includes UK Biobank |
| Biliary | Ji SG (2016) | doi.org/10.1038/ng.3745 | Primary sclerosing cholangitis | Broadly | European | 2,871 | 12,019 | Yes | Exclude | Overlaps with larger study included in analysis |
| Cardiovascular | Jiang L (2021) | doi.org/10.1038/s41588-021-00954-4 | Atherosclerosis of the extremities | Broadly | European | 635 | 455,713 | Yes | Exclude | Includes UK Biobank |
| Cardiovascular | Backman JD (2021) | doi.org/10.1038/s41586-021-04103-z | ICD10 I70.2: Atherosclerosis of native arteries of the extremities | Broadly | European | 1,065 | 386,835 | Yes | Exclude | Includes UK Biobank |

| Organ | Author (year) | DOI | Disease | Always or broadly fibrotic | Ancestry | Cases | Controls | Summary statistics available | Include/ exclude | Reason for exclusion |
|---|---|---|---|---|---|---|---|---|---|---|
| Cardiovascular | Backman JD (2021) | doi.org/10.1038/s41586-021-04103-z | ICD10 I65.2: Occlusion and stenosis of carotid artery | Broadly | European | 1,229 | 386,700 | Yes | Exclude | Includes UK Biobank |
| Cardiovascular | Zhou J (2020) | doi.org/10.1371/journal.pone.0238304 | Coronary artery disease | Non-fibrotic | European | 2,824 (quantitative trait) | | Yes | Exclude | Non-fibrotic disease |
| Cardiovascular | Schunkert H (2011) | doi.org/10.1038/ng.784 | Coronary artery disease | Non-fibrotic | European | 22,233 | 64,762 | Yes | Exclude | Non-fibrotic disease |
| Cardiovascular | Matsunaga H (2020) | doi.org/10.1161/circgen.119.002670 | Coronary artery disease | Non-fibrotic | East Asian | 15,302 | 36,140 | Yes | Exclude | Non-fibrotic disease |
| Cardiovascular | Sakaue S (2021) | doi.org/10.1038/s41588-021-00931-x | Behcets disease | Broadly | East Asian | 78 | 171,966 | Yes | Exclude | Overlaps with larger study included in analysis |
| Cardiovascular | Sakaue S (2021) | doi.org/10.1038/s41588-021-00931-x | Behcets disease | Broadly | European | 27 | 317,225 | Yes | Exclude | Includes UK Biobank |
| Cardiovascular | Ortiz-Fernández L (2016) | doi.org/10.1371/journal.pone.0161305 | Behcets disease | Broadly | European | 278 | 1,517 | No | Exclude | Summary statistics not available |
| Cardiovascular | Sakaue S (2021) | doi.org/10.1038/s41588-021-00931-x | Dilated cardiomyopathy | Non-fibrotic | East Asian | 417 | 177,745 | Yes | Exclude | Non-fibrotic disease |
| Cardiovascular | Sakaue S (2021) | doi.org/10.1038/s41588-021-00931-x | Dilated cardiomyopathy | Non-fibrotic | European | 1444 | 353,937 | Yes | Exclude | Non-fibrotic disease |
| Cardiovascular | Backman JD (2021) | doi.org/10.1038/s41586-021-04103-z | Dilated cardiomyopathy | Non-fibrotic | European | 858 | 387,072 | Yes | Exclude | Non-fibrotic disease |
| Cardiovascular | Sakaue S (2021) | doi.org/10.1038/s41588-021-00931-x | Hypertrophic cardiomyopathy | Non-fibrotic | East Asian | 383 | 177,745 | Yes | Exclude | Non-fibrotic disease |
| Cardiovascular | Sakaue S (2021) | doi.org/10.1038/s41588-021-00931-x | Hypertrophic cardiomyopathy | Non-fibrotic | European | 507 | 489,220 | Yes | Exclude | Includes UK Biobank |
| Cardiovascular | Sakaue S (2021) | doi.org/10.1038/s41588-021-00931-x | Ventricular arrhythmia | Broadly | East Asian | 1,673 | 155,540 | Yes | Include | - |
| Cardiovascular | Sakaue S (2021) | doi.org/10.1038/s41588-021-00931-x | Ventricular arrhythmia | Broadly | European | 1018 | 327,198 | Yes | Exclude | Includes UK Biobank |
| Cardiovascular | Zhou J (2020) | doi.org/10.1371/journal.pone.0238304 | Cardiomyopathy | Non-fibrotic | European | 2,824 (quantitative trait) | | Yes | Exclude | Non-fibrotic disease |
| Cardiovascular | Zhou J (2020) | doi.org/10.1371/journal.pone.0238304 | Arrhythmia | Broadly | European | 89 (quantitative trait) | | Yes | Exclude | Unable to align alleles |

| Organ | Author (year) | DOI | Disease | Always or broadly fibrotic | Ancestry | Cases | Controls | Summary statistics available | Include/ exclude | Reason for exclusion |
|---|---|---|---|---|---|---|---|---|---|---|
| Cardiovascular | Zhou W (2022) | doi.org/10.1016/j.xgen.2022.100192 | Hypertrophic cardiomyopathy | Non-fibrotic | East Asian | 1,110 | 268,962 | Yes | Exclude | Non-fibrotic disease |
| Cardiovascular | Villard E (2011) | doi.org/10.1093/eurheartj/ehr105 | Dilated cardiomyopathy | Non-fibrotic | European | 1,179 | 1,108 | No | Exclude | Summary statistics not available |
| Cardiovascular | Meder B (2013) | doi.org/10.1093/eurheartj/eht251 | Dilated cardiomyopathy | Non-fibrotic | European | 909 | 2,120 | No | Exclude | Summary statistics not available |
| Cardiovascular | Garnier S (2021) | 10.1093/eurheartj/ehab030 | Dilated cardiomyopathy | Non-fibrotic | European | 2,651 | 4,329 | No | Exclude | Summary statistics not available |
| Pulmonary | Aksit MA (2022) | 10.1016/j.ajhg.2022.09.004 | Cystic fibrosis-related diabetes | Non-fibrotic | European | 1,396 | 2,632 | Yes | Exclude | Non-fibrotic disease |
| Pulmonary | Gu Y (2009) | doi.org/10.1038/nature07811 | Cystic fibrosis severity | Broadly | European | 320 | NA | No | Exclude | Summary statistics not available<br><br>Not a disease susceptibility GWAS |
| Pulmonary | Wright FA (2011) | 10.1038/ng.838 | Cystic fibrosis severity | Broadly | European | 2317 | NA | No | Exclude | Summary statistics not available<br><br>Not a disease susceptibility GWAS |
| Diabetes | Zhou J (2020) | 10.1371/journal.pone.0238304 | Diabetes mellitus | Broadly | European | 2,823 (quantitative trait) | | Yes | Exclude | Unable to align alleles<br><br>Not a disease susceptibility GWAS |
| Diabetes | Mansour Aly D (2021) | 10.1038/s41588-021-00948-2 | Severe insulin-deficient type 2 diabetes | Non-fibrotic | European | 1,193 | 2,744 | Yes | Exclude | Non-fibrotic disease |
| Diabetes | Mansour Aly D (2021) | 10.1038/s41588-021-00948-2 | Type 2 diabetes | Broadly | European | 9,486 | 2,744 | Yes | Include | - |
| Diabetes | Backman JD (2021) | doi.org/10.1038/s41586-021-04103-z | Diabetes | Broadly | European | 17,927 | 313,827 | Yes | Exclude | Includes UK Biobank |
| Diabetes | Loh M (2022) | doi.org/10.1038/s42003-022-03248-5 | Type ii diabetes | Broadly | South Asian | 16,677 | 33,856 | Yes | Exclude | Includes UK Biobank |

| Organ | Author (year) | DOI | Disease | Always or broadly fibrotic | Ancestry | Cases | Controls | Summary statistics available | Include/ exclude | Reason for exclusion |
|---|---|---|---|---|---|---|---|---|---|---|
| Diabetes | Lee CJ | 10.1038/s42003-022-04168-0 | Type 2 diabetes | Broadly | East Asian | 3,844 | 59,333 | Yes | Exclude | Overlaps with larger study included in analysis |
| Diabetes | Barrett JC (2009) | 10.1038/ng.381 | Type 1 diabetes | Broadly | European | 7,514 | 9,045 | Yes | Exclude | Overlaps with larger study included in analysis |
| Diabetes | Onengut-Gumuscu S (2015) | doi.org/10.1038/ng.3245 | Type 1 diabetes | Broadly | European | 6,683 | 12,173 | Yes | Exclude | Overlaps with larger study included in analysis |
| Diabetes | Onengut-Gumuscu S (2015) | doi.org/10.1038/ng.3245 | Type 1 diabetes | Broadly | European | 6,683 | 12,173 | Yes | Exclude | Overlaps with larger study included in analysis |
| Diabetes | Forgetta V (2020) | doi.org/10.2337/db19-0831 | Type 1 diabetes | Broadly | European | 9,266 | 15,574 | Yes | Exclude | Overlaps with larger study included in analysis |
| Diabetes | Qu HQ (2021) | doi.org/10.1038/s42003-021-02368-8 | Type 1 diabetes with low genetic risk score | Broadly | European | 957 | 12,323 | Yes | Exclude | Not a disease susceptibility GWAS |
| Diabetes | Inshaw JRJ (2021) | doi.org/10.1007/s00125-021-05428-0 | Type 1 diabetes | Broadly | European | 7,467 | 10,218 | Yes | Exclude | Overlaps with larger study included in analysis |
| Diabetes | Chen J (2019) | doi.org/10.1007/s00125-019-4880-7 | Type 2 diabetes | Broadly | African | 2,633 | 1,714 | Yes | Include | - |
| Diabetes | Bonàs-Guarch S (2018) | doi.org/10.1038/s41467-017-02380-9 | Type 2 diabetes | Broadly | European | 12,931 | 57,196 | Yes | Include | - |
| Diabetes | Suzuki K (2019) | doi.org/10.1038/s41588-018-0332-4 | Type 2 diabetes | Broadly | East Asian | 36,614 | 155,150 | Yes | Exclude | Overlaps with larger study included in analysis |
| Diabetes | Spracklen CN (2020) | doi.org/10.1038/s41586-020-2263-3 | Type 2 diabetes | Broadly | East Asian | 77,418 | 356,122 | Yes | Include | - |
| Skin | Rothwell S (2023) | doi.org/10.1002/art.42434 | Dermatomyositis | Broadly | European | 817 | 10,260 | No | Exclude | Summary statistics not available |

| Organ | Author (year) | DOI | Disease | Always or broadly fibrotic | Ancestry | Cases | Controls | Summary statistics available | Include/ exclude | Reason for exclusion |
|---|---|---|---|---|---|---|---|---|---|---|
| Skin | Riesmeijer (2024) | www.nature.com/articles/s41467-023-44451-0#Sec31 | Dupuytrens disease | Always | European | 7,584 | 28,343 | Yes | Include | - |
| Skin | Julia A (2018) | 10.1186/s13075-018-1604-1 | Systemic lupus erythematosus | Broadly | European | 907 | 1,524 | Yes | Include | - |
| Skin | Langefeld CD (2017) | 10.1038/ncomms16021 | Systemic lupus erythematosus | Broadly | European | 6,748 | 11,516 | Yes | Exclude | Effect size estimates missing |
| Skin | Backman JD (2021) | doi.org/10.1038/s41586-021-04103-z | ICD10 M72.0: Palmar fascial fibromatosis [Dupuytren] | Always | European | 4,263 | 383,667 | Yes | Exclude | Includes UK Biobank |
| Skin | Backman JD (2021) | doi.org/10.1038/s41586-021-04103-z | ICD10 L90.5: Scar conditions and fibrosis of skin | Always | European | 2,723 | 385,128 | Yes | Exclude | Includes UK Biobank |
| Skin | Bentham J (2015) | doi.org/10.1038/ng.3434 | Systemic lupus erythematosus | Broadly | European | 5,201 | 9,066 | Yes | Include | - |
| Skin | Wang YF (2021) | doi.org/10.1038/s41467-021-21049-y | Systemic lupus erythematosus | Broadly | East Asian | 4,222 | 8,431 | Yes | Include | - |
| Skin | Song Q (2021) | doi.org/10.1093/rheumatology/keab016 | Systemic lupus erythematosus | Broadly | East Asian | 512 | 994 | Yes | Exclude | Overlaps with larger study included in analysis |
| Skin | Backman JD (2021) | doi.org/10.1038/s41586-021-04103-z | Systemic lupus erythematosis | Broadly | European | 523 | 331,231 | Yes | Exclude | Includes UK Biobank |
| Intestinal-pancreas | Sakaue S (2021) | doi.org/10.1038/s41588-021-00931-x | Acute pancreatitis | Broadly | East Asian | 827 | 177,471 | Yes | Include | - |
| Intestinal-pancreas | Sakaue S (2021) | doi.org/10.1038/s41588-021-00931-x | Acute pancreatitis | Broadly | European | 3,798 | 476,104 | Yes | Exclude | Includes UK Biobank |
| Intestinal-pancreas | Blackman JD (2021) | doi.org/10.1038/s41586-021-04103-z | ICD10 K85: Acute pancreatitis | Broadly | European | 2,460 | 385,091 | Yes | Exclude | Includes UK Biobank |
| Intestinal-pancreas | Bourgault J (2023) | doi.org/10.1053/j.gastro.2023.01.028 | Acute pancreatitis | Broadly | European | 10,630 | 844,679 | Yes | Exclude | Includes UK Biobank |
| Intestinal-pancreas | Franke A (2010) | doi.org/10.1038/ng.717 | Crohn's disease | Broadly | European | 6,333 | 15,056 | Yes | Exclude | Summary statistics are all NA |

| Organ | Author (year) | DOI | Disease | Always or broadly fibrotic | Ancestry | Cases | Controls | Summary statistics available | Include/ exclude | Reason for exclusion |
|---|---|---|---|---|---|---|---|---|---|---|
| Intestinal-pancreas | Liu JZ (2015) | doi.org/10.1038/ng.3359 | Crohn's disease | Broadly | Multi-ancestry | 5,956 | 14,927 | Yes | Exclude | Multi-ancestry analysis |
| Intestinal-pancreas | Garcia-Etxebarria K (2022) | doi.org/10.1038/s41598-022-07401-2 | Crohn's disease | Broadly | European | 284 | 935 | Yes | Include | - |
| Intestinal-pancreas | Blackman JD (2021) | doi.org/10.1038/s41586-021-04103-z | Crohn's disease | Broadly | European | 1,324 | 330,430 | Yes | Exclude | Includes UK Biobank |
| Intestinal-pancreas | Jung S (2022) | doi.org/10.1093/hmg/ddac101 | Crohn's disease | Broadly | East Asian | 1,621 | 4,419 | Yes | Exclude | Effect size estimates missing |
| Intestinal-pancreas | Sakaue S (2021) | doi.org/10.1038/s41588-021-00931-x | Chronic pancreatitis | Broadly | East Asian | 457 | 177,471 | Yes | Exclude | Overlaps with larger study included in analysis |
| Intestinal-pancreas | Sakaue S (2021) | doi.org/10.1038/s41588-021-00931-x | Chronic pancreatitis | Broadly | European | 1,424 | 476,104 | Yes | Exclude | Includes UK Biobank |
| Intestinal-pancreas | Blackman JD (2021) | doi.org/10.1038/s41586-021-04103-z | ICD10 K52.8: Other specified noninfective gastroenteritis and colitis | Broadly | European | 650 | 387,279 | Yes | Exclude | Includes UK Biobank |
| Intestinal-pancreas | Blackman JD (2021) | doi.org/10.1038/s41586-021-04103-z | ICD10 A09.9: Gastroenteritis and colitis of unspecified origin | Broadly | European | 8,538 | 379,392 | Yes | Exclude | Includes UK Biobank |
| Intestinal-pancreas | Blackman JD (2021) | doi.org/10.1038/s41586-021-04103-z | ICD10 K86.1: Other chronic pancreatitis | Broadly | European | 535 | 387,394 | Yes | Exclude | Includes UK Biobank |
| Intestinal-pancreas | Sakaue S (2021) | doi.org/10.1038/s41588-021-00931-x | Ulcerative colitis | Broadly | East Asian | 314 | 178,375 | Yes | Exclude | Overlaps with larger study included in analysis |
| Intestinal-pancreas | Sakaue S (2021) | doi.org/10.1038/s41588-021-00931-x | Ulcerative colitis | Broadly | European | 5,371 | 412,561 | Yes | Exclude | Includes UK Biobank |
| Intestinal-pancreas | Garcia-Etxebarria K (2022) | doi.org/10.1038/s41598-022-07401-2 | Ulcerative colitis | Broadly | European | 208 | 935 | Yes | Exclude | Overlaps with larger study included in analysis |
| Intestinal-pancreas | Blackman JD (2021) | doi.org/10.1038/s41586-021-04103-z | Ulcerative colitis | Broadly | European | 2,363 | 329,391 | Yes | Exclude | Includes UK Biobank |

| Organ | Author (year) | DOI | Disease | Always or broadly fibrotic | Ancestry | Cases | Controls | Summary statistics available | Include/ exclude | Reason for exclusion |
|---|---|---|---|---|---|---|---|---|---|---|
| Liver | Buch S (2015) | 10.1038/ng.3417 | Alcoholic liver cirrhosis | Always | European | 712 | 1,426 | Yes | Include | - |
| Liver | Buch S (2015) | 10.1038/ng.3417 | Alcoholic liver cirrhosis ( Secondary adjusted analysis) | Always | European | 712 | 1,426 | Yes | Exclude | Overlaps with larger study included in analysis |
| Liver | Sakaue S (2021) | doi.org/10.1038/s41588-021-00931-x | Chronic hepatitis B virus infection | Non-fibrotic | East Asian | 2,234 | 169,588 | Yes | Exclude | Non-fibrotic disease |
| Liver | Sakaue S (2021) | doi.org/10.1038/s41588-021-00931-x | Chronic hepatitis B virus infection | Non-fibrotic | European | 145 | 351,740 | Yes | Exclude | Includes UK Biobank<br><br>Non-fibrotic disease |
| Liver | Sakaue S (2021) | doi.org/10.1038/s41588-021-00931-x | Chronic hepatitis C virus infection | Broadly | East Asian | 7110 | 169,588 | Yes | Exclude | Overlaps with larger study included in analysis |
| Liver | Sakaue S (2021) | doi.org/10.1038/s41588-021-00931-x | Chronic hepatitis C virus infection | Broadly | European | 273 | 351,740 | Yes | Exclude | Includes UK Biobank |
| Liver | Sakaue S (2021) | doi.org/10.1038/s41588-021-00931-x | Cirrhosis of liver | Always | East Asian | 2,551 | 176,175 | Yes | Include | - |
| Liver | Sakaue S (2021) | doi.org/10.1038/s41588-021-00931-x | Cirrhosis of liver | Always | European | 122 | 347,284 | Yes | Exclude | Includes UK Biobank |
| Liver | Backman JD (2021) | doi.org/10.1038/s41586-021-04103-z | Cirrhosis of liver | Always | European | 1,995 | 381,355 | Yes | Exclude | Includes UK Biobank |
| Liver | Backman JD (2021) | doi.org/10.1038/s41586-021-04103-z | Fibrosis and cirrhosis of liver | Always | European | 1,292 | 386,463 | Yes | Exclude | Includes UK Biobank |
| Liver | Jiang L (2021) | 10.1038/s41588-021-00954-4 | Cirrhosis of liver | Always | European | 240 | 456,108 | Yes | Exclude | Includes UK Biobank |
| Liver | Namjou B (2019) | 10.1186/s12916-019-1364-z | Liver fibrosis in non-alcoholic fatty liver disease | Always | European | 235 (quantitative trait) | | Yes | Exclude | Not a disease susceptibility GWAS |
| Liver | Namjou B (2019) | 10.1186/s12916-019-1364-z | Non-alcoholic fatty liver disease | Broadly | European | 1106 | 8,571 | Yes | Include | - |
| Liver | Anstee QM (2020) | 10.1016/j.jhep.2020.04.003 | Non-alcoholic fatty liver disease | Broadly | European | 1,483 | 17,781 | Yes | Include | - |
| Liver | Fairfield CJ (2022) | 10.1002/hep4.1805 | Non-alcoholic fatty liver disease | Broadly | European | 4761 | 373,227 | Yes | Exclude | Includes UK Biobank |

| Organ | Author (year) | DOI | Disease | Always or broadly fibrotic | Ancestry | Cases | Controls | Summary statistics available | Include/ exclude | Reason for exclusion |
|---|---|---|---|---|---|---|---|---|---|---|
| Liver | Ghodsian N (2021) | 10.1016/j.xcrm.2021.100437 | Non-alcoholic fatty liver disease | Broadly | European | 8434 | 770,180 | Yes | Exclude | Includes UK Biobank |
| Liver | Beaudoin JJ (2017) | 10.1080/00365521.2017.1359664 | Alcoholic hepatitis | Not-fibrotic | European | 90 | 93 | No | Exclude | Summary statistics not available<br><br>Non-fibrotic disease |
| Liver | Schwantes-An TH (2021) | doi.org/10.1002/hep.31535 | Alcohol-related cirrhosis | Always | European | 1128 | 849 | Yes | Exclude | Unable to align alleles |
| Liver | Speliotes EK (2011) | doi.org/10.1371/journal.pgen.1001324 | Non-alcoholic fatty liver disease | Broadly | European | 880 | 6,296 | No | Exclude | Summary statistics not available |
| Lymphatic | Jiang L (2021) | doi.org/10.1038/s41588-021-00954-4 | ICD10 C81.1: Nodular sclerosis classical Hodgkin | Broadly | European | 126 | 456,222 | Yes | Exclude | Includes UK Biobank |
| Lymphatic | Sud A (2017) | doi.org/10.1038/s41467-017-00320-1 | Nodular sclerosis Hodgkin lymphoma | Broadly | European | 1,278 | 14,325 | No | Exclude | Summary statistics not available |
| Pulmonary | Sakaue S (2021) | doi.org/10.1038/s41588-021-00931-x | Interstitial lung disease | Broadly | East Asian | 1,046 | 176,974 | Yes | Exclude | Overlaps with larger study included in analysis |
| Pulmonary | Sakaue S (2021) | doi.org/10.1038/s41588-021-00931-x | Interstitial lung disease | Broadly | European | 2,267 | 467,560 | Yes | Exclude | Includes UK Biobank |
| Pulmonary | Sakaue S (2021) | doi.org/10.1038/s41588-021-00931-x | Pneumoconiosis | Broadly | East Asian | 85 | 176,974 | Yes | Exclude | Overlaps with larger study included in analysis |
| Pulmonary | Sakaue S (2021) | doi.org/10.1038/s41588-021-00931-x | Pneumoconiosis | Broadly | European | 433 | 478,607 | Yes | Exclude | Includes UK Biobank |
| Pulmonary | Jiang L (2021) | doi.org/10.1038/s41588-021-00954-4 | Pulmonary fibrosis | Always | European | 363 | 455,985 | Yes | Exclude | Includes UK Biobank |
| Pulmonary | Sakaue S (2021) | doi.org/10.1038/s41588-021-00931-x | Pulmonary fibrosis | Always | East Asian | 126 | 176,974 | Yes | Exclude | Overlaps with larger study included in analysis |
| Pulmonary | Sakaue S (2021) | doi.org/10.1038/s41588-021-00931-x | Pulmonary fibrosis | Always | European | 1566 | 467,560 | Yes | Exclude | Includes UK Biobank |

| Organ | Author (year) | DOI | Disease | Always or broadly fibrotic | Ancestry | Cases | Controls | Summary statistics available | Include/ exclude | Reason for exclusion |
|---|---|---|---|---|---|---|---|---|---|---|
| Pulmonary | Allen R (2022) | doi.org/10.1136/thoraxjnl-2021-218577 | Idiopathic pulmonary fibrosis | Always | European | 4125 | 20,464 | Yes | Include | - |
| Pulmonary | Duckworth A (2021) | doi.org/10.1016/s2213-2600(20)30364-7 | Idiopathic pulmonary fibrosis | Always | European | 1369 | 435,866 | Yes | Exclude | Includes UK Biobank |
| Pulmonary | Partanen JJ (2022) | doi.org/10.1016/j.xgen.2022.100181 | Idiopathic pulmonary fibrosis | Always | African | 169 | 8,368 | Yes | Include | - |
| Pulmonary | Partanen JJ (2022) | doi.org/10.1016/j.xgen.2022.100181 | Idiopathic pulmonary fibrosis | Always | East Asian | 1,210 | 254,409 | Yes | Include | - |
| Pulmonary | Fingerlin TE (2013) | doi.org/10.1038/ng.2609 | Idiopathic pulmonary fibrosis | Always | European | 1,161 | 4,683 | Yes | Exclude | Overlaps with larger study included in analysis |
| Pulmonary | Partanen JJ (2022) | doi.org/10.1016/j.xgen.2022.100181 | Idiopathic pulmonary fibrosis | Always | European | 6,743 | 1,056,639 | Yes | Exclude | Includes UK Biobank |
| Reproductive | Sobalska-Kwapis M (2017) | doi.org/10.1016/j.ejogrb.2017.08.037 | Endometriosis | Broadly | European | 171 | 2,934 | No | Exclude | Summary statistics not available |
| Reproductive | Backman JD (2021) | doi.org/10.1038/s41586-021-04103-z | ICD10 N80: Endometriosis | Broadly | European | 3,142 | 174,752 | Yes | Exclude | Includes UK Biobank |
| Reproductive | Rahmioglu N (2023) | doi.org/10.1038/s41588-023-01323-z | Endometriosis | Broadly | Multi-ancestry | 23,360 | 450,668 | Yes | Exclude | Multi-ancestry analysis |
| Reproductive | Rahmioglu N (2023) | doi.org/10.1038/s41588-023-01323-z | Endometriosis | Broadly | European | 21,779 | 449,087 | Yes | Exclude | Includes UK Biobank |
| Reproductive | Backman JD (2021) | doi.org/10.1038/s41586-021-04103-z | ICD10 N50.8: Other specified disorders of male genital organs | Broadly | European | 3,016 | 171,235 | Yes | Exclude | Includes UK Biobank |
| Skeletal | Stahl EA (2010) | doi.org/10.1038/ng.582 | Rheumatoid arthritis | Broadly | European | 5,539 | 20,169 | Yes | Exclude | Overlaps with larger study included in analysis |
| Skeletal | Okada Y (2014) | doi.org/10.1038/nature12873 | Rheumatoid arthritis | Broadly | Multi-ancestry | 19,234 | 60,565 | Yes | Exclude | Multi-ancestry analysis |
| Skeletal | Eyre S (2012) | doi.org/10.1038/ng.2462 | Rheumatoid arthritis | Broadly | European | 13,838 | 33,742 | Yes | Exclude | Overlaps with larger study included in analysis |

| Organ | Author (year) | DOI | Disease | Always or broadly fibrotic | Ancestry | Cases | Controls | Summary statistics available | Include/ exclude | Reason for exclusion |
|---|---|---|---|---|---|---|---|---|---|---|
| Skeletal | Ha E (2021) | doi.org/10.1136/annrheumdis-2020-219065 | Rheumatoid arthritis | Broadly | Multi-ancestry | 22,628 | 288,664 | Yes | Exclude | Multi-ancestry analysis |
| Skeletal | Sakaue S (2021) | doi.org/10.1038/s41588-021-00931-x | Rheumatoid arthritis | Broadly | East Asian | 5,348 | 173,268 | Yes | Exclude | Overlaps with larger study included in analysis |
| Skeletal | Backman JD (2021) | doi.org/10.1038/s41586-021-04103-z | Rheumatoid arthritis | Broadly | European | 8,561 | 295,029 | Yes | Exclude | Includes UK Biobank |
| Skeletal | Ishigaki K (2022) | doi.org/10.1038/s41588-022-01213-w | Rheumatoid arthritis | Broadly | Multi-ancestry | 35,871 | 240,149 | Yes | Exclude | Multi-ancestry analysis |
| Skeletal | Ishigaki K (2022) | doi.org/10.1038/s41588-022-01213-w | Rheumatoid arthritis | Broadly | European | 22,350 | 74,823 | Yes | Include | - |
| Skeletal | Ishigaki K (2022) | doi.org/10.1038/s41588-022-01213-w | Rheumatoid arthritis | Broadly | East Asian | 11,025 | 162,608 | Yes | Include | - |
| Systemic | Jiang L (2021) | 10.1038/s41588-021-00954-4 | Systemic sclerosis | Broadly | European | 104 | 456,244 | Yes | Exclude | Includes UK Biobank |
| Systemic | López-Isac E (2019) | doi.org/10.1038/s41467-019-12760-y | Systemic sclerosis | Broadly | European | 9,095 | 17,584 | Yes | Include | - |
| Systemic | Márquez A (2018) | doi.org/10.1186/s13073-018-0604-8 | Systemic sclerosis | Broadly | European | 3,477 | 22,308 | No | Exclude | Summary statistics not available |
| Systemic | Taylor KE (2017) | doi.org/10.1002/art.40040 | Sjögren's syndrome | Broadly | Multi-ancestry | 1,405 | 4,747 | Yes | Exclude | Multi-ancestry analysis |
| Systemic | Taylor KE (2017) | doi.org/10.1002/art.40040 | Sjögren's syndrome | Broadly | East Asian | 460 | 1,125 | Yes | Exclude | Standard error estimates missing |
| Systemic | Taylor KE (2017) | doi.org/10.1002/art.40040 | Sjögren's syndrome | Broadly | European | 585 | 1,546 | Yes | Include | - |
| Systemic | Glanville KP (2021) | doi.org/10.1016/j.bpsgos.2021.03.002 | Sjögren's syndrome | Broadly | European | 647 | 324,074 | Yes | Exclude | Includes UK Biobank |
| Systemic | Sakaue S (2021) | doi.org/10.1038/s41588-021-00931-x | Sjögren's syndrome | Broadly | East Asian | 303 | 175,599 | Yes | Include | - |
| Systemic | Sakaue S (2021) | doi.org/10.1038/s41588-021-00931-x | Sjögren's syndrome | Broadly | European | 1,296 | 482,717 | Yes | Exclude | Includes UK Biobank |
| Systemic | Jiang L (2021) | 10.1038/s41588-021-00954-4 | Sicca syndrome (PheCode 709.2) | Broadly | European | 99 | 456,249 | Yes | Exclude | Includes UK Biobank |

| Organ | Author (year) | DOI | Disease | Always or broadly fibrotic | Ancestry | Cases | Controls | Summary statistics available | Include/ exclude | Reason for exclusion |
|---|---|---|---|---|---|---|---|---|---|---|
| Systemic | Backman JD (2021) | doi.org/10.1038/s41586-021-04103-z | ICD10 M35.0: Sicca syndrome | Broadly | European | 711 | 387,080 | Yes | Exclude | Includes UK Biobank |
| Urinary | Backman JD (2021) | doi.org/10.1038/s41586-021-04103-z | ICD10 N03: Chronic nephritic syndrome | Broadly | European | 978 | 386,890 | Yes | Exclude | Includes UK Biobank |
| Urinary | Sakaue S (2021) | doi.org/10.1038/s41588-021-00931-x | Polycystic Kidney Disease | Broadly | East Asian | 510 | 178,216 | Yes | Include | - |
| Urinary | Sakaue S (2021) | doi.org/10.1038/s41588-021-00931-x | Polycystic Kidney Disease | Broadly | European | 424 | 355,431 | Yes | Exclude | Includes UK Biobank |
| Urinary | Wuttke M (2019) | doi.org/10.1038/s41588-019-0407-x | Chronic kidney disease | Broadly | Multi-ancestry | 64,164 | 561,055 | Yes | Exclude | Multi-ancestry analysis |
| Urinary | Hishida A (2018) | doi.org/10.1159/000488946 | Chronic kidney disease | Broadly | East Asian | 939 | 10,344 | No | Exclude | Summary statistics not available |
| Urinary | Yun S (2019) | doi.org/10.1007/s10157-019-01731-8 | Chronic kidney disease | Broadly | East Asian | 281 | 3,336 | No | Exclude | Summary statistics not available |
| Urinary | Wuttke M (2016) | 10.1093/ndt/gfv342 | Chronic kidney disease | Broadly | European | 658 | 1,347 | No | Exclude | Summary statistics not available |
| Urinary | Pattaro C (2016) | doi.org/10.1038/ncomms10023 | Chronic kidney disease | Broadly | European | 12,385 | 104,780 | Yes | Exclude | Overlaps with larger study included in analysis |
| Urinary | Wojcik GL (2019) | doi.org/10.1038/s41586-019-1310-4 | Chronic kidney disease | Broadly | Multi-ancestry | 4,154 | 41,573 | Yes | Exclude | Multi-ancestry analysis |
| Urinary | Wuttke M (2019) | doi.org/10.1038/s41588-019-0407-x | Chronic kidney disease | Broadly | European | 41,395 | 439,303 | Yes | Include | - |
| Urinary | Backman JD (2021) | doi.org/10.1038/s41586-021-04103-z | ICD10 N18: Chronic kidney disease | Broadly | European | 10,144 | 372,699 | Yes | Exclude | Includes UK Biobank |

#### Supplementary Table 3: Sample sizes of analyses across all ancestry groups

| | Ancestry | Broadly fibrotic | | | | | | Always fibrotic | | | | | |
|---|---|---|---|---|---|---|---|---|---|---|---|---|---|
| | | Individual studies | | | Sample size of meta-analysis | | | Individual studies | | | Sample size of meta-analysis | | |
| | | Source: disease | Cases | Controls | Cases | Controls | Total | Source: disease | Cases | Controls | Cases | Controls | Total |
| Biliary | European | UK Biobank: Biliary broadly fibrotic | 4,916 | 49,160 | **12,937** | **65,649** | **78,586** | UK Biobank: Biliary always fibrotic | 883 | 8,830 | **883** | **8,830** | **9,713** |
| | | Cordell (2021): Primary biliary cholangitis | 8,021 | 16,489 | | | | | | | | | |
| | East Asian | Nakamura (2012): Primary biliary cholangitis | 487 | 476 | **487** | **476** | **963** | - | - | - | - | - | - |
| | South Asian | UK Biobank: Biliary broadly fibrotic | 89 | 890 | **89** | **890** | **979** | - | - | - | - | - | - |
| | African | UK Biobank: Biliary broadly fibrotic | 33 | 330 | **33** | **330** | **363** | - | - | - | - | - | - |
| Cardiovascular | European | UK Biobank: Cardiovascular broadly fibrotic | 33,671 | 336,710 | **33,671** | **336,710** | **370,381** | - | - | - | - | - | - |
| | East Asian | Sakaue (2021): Ventricular arrhythmia | 1,673 | 155,540 | **1,673** | **155,540** | **157,213** | - | - | - | - | - | - |
| | South Asian | UK Biobank: Cardiovascular broadly fibrotic | 1,194 | 8,844 | **1,194** | **8,844** | **10,038** | - | - | - | - | - | - |
| | African | UK Biobank: Cardiovascular broadly fibrotic | 334 | 3,440 | **344** | **3,440** | **3,784** | - | - | - | - | - | - |
| Diabetes | European | UK Biobank: Diabetes broadly fibrotic | 24,907 | 249,070 | **47,324** | **309,010** | **356,334** | - | - | - | - | - | - |
| | | Mansour Aly (2021): Type 2 diabetes | 9,486 | 2,744 | | | | | | | | | |
| | | Bonàs-Guarch (2018): Type 2 diabetes | 12,931 | 57,196 | | | | | | | | | |
| | East Asian | UK Biobank: Diabetes broadly fibrotic | 151 | 1,510 | **77,569** | **357,632** | **435,201** | - | - | - | - | - | - |
| | | Spracklen (2020): Type 2 diabetes | 77,418 | 356,122 | | | | | | | | | |
| | South Asian | UK Biobank: Diabetes broadly fibrotic | 1,856 | 8,182 | **1,856** | **8,182** | **10,038** | - | - | - | - | - | - |
| | African | UK Biobank: Diabetes broadly fibrotic | 911 | 6,551 | **3,544** | **8,265** | **11,809** | - | - | - | - | - | - |
| | | Chen (2019): Type 2 diabetes | 2,633 | 1,714 | | | | | | | | | |
| | European | UK Biobank: Intestinal-pancreatic broadly fibrotic | 49,710 | 368,292 | **49,994** | **369,227** | **419,221** | - | - | - | - | - | - |

| | | | | | | | | | | | | | |
|---|---|---|---|---|---|---|---|---|---|---|---|---|---|
| | | Garcia-Etxebarria (2022): Crohn's disease | 284 | 935 | | | | | | | | | |
| **Intestinal-pancreatic** | **East Asian** | UK Biobank: Intestinal-pancreatic broadly fibrotic | 65 | 650 | **892** | **178,121** | **179,013** | - | - | - | - | - | - |
| | | Sakaue (2021): Acute pancreatitis | 827 | 177,471 | | | | | | | | | |
| | **South Asian** | UK Biobank: Intestinal-pancreatic broadly fibrotic | 806 | 8,060 | **806** | **8,060** | **8,866** | - | - | - | - | - | - |
| | **African** | UK Biobank: Intestinal-pancreatic broadly fibrotic | 480 | 4,800 | **480** | **4,800** | **5,280** | - | - | - | - | - | - |
| **Liver** | **European** | UK Biobank: Liver broadly fibrotic | 4,437 | 44,370 | **7,738** | **72,148** | **79,886** | UK Biobank: Liver always fibrotic | 1,414 | 14,140 | **2,126** | **15,566** | **17,692** |
| | | Buch (2015): Alcohol-related liver cirrhosis | 712 | 1,426 | | | | | | | | | |
| | | Namjou (2019): Non-alcoholic fatty liver disease | 1,106 | 8,571 | | | | Buch (2015): Alcohol-related liver cirrhosis | 712 | 1,426 | | | |
| | | Anstee (2020): Non-alcoholic fatty liver disease | 1,483 | 17,781 | | | | | | | | | |
| | **East Asian** | Sakaue (2021): Cirrhosis of liver | 2,551 | 176,175 | **2,551** | **176,175** | **178,726** | Sakaue (2021): Cirrhosis of liver | 2,551 | 176,175 | **2,551** | **176,175** | **178,726** |
| | **South Asian** | UK Biobank: Liver broadly fibrotic | 138 | 1,380 | **138** | **1,380** | **1,518** | - | - | - | - | - | - |
| | **African** | UK Biobank: Liver broadly fibrotic | 71 | 710 | **71** | **710** | **781** | - | - | - | - | - | - |
| **Lymphatic** | **European** | UK Biobank: Lymphatic broadly fibrotic | 334 | 3,340 | **334** | **3,340** | **3,674** | - | - | - | - | - | - |
| | **East Asian** | - | - | - | - | - | - | - | - | - | - | - | - |
| | **South Asian** | - | - | - | - | - | - | - | - | - | - | - | - |
| | **African** | - | - | - | - | - | - | - | - | - | - | - | - |
| **Pulmonary** | **European** | UK Biobank: Pulmonary broadly fibrotic | 2,598 | 25,980 | **6,723** | **46,444** | **53,167** | UK Biobank: Pulmonary always fibrotic | 1,764 | 17,640 | **5,889** | **38,104** | **43,993** |
| | | Allen (2022): Idiopathic pulmonary fibrosis | 4,125 | 20,464 | | | | Allen (2022): Idiopathic pulmonary fibrosis | 4,125 | 20,464 | | | |
| | **East Asian** | Partanen (2022): Idiopathic pulmonary fibrosis | 1,210 | 254,409 | **1,210** | **254,409** | **255,619** | Partanen (2022): Idiopathic pulmonary fibrosis | 1,210 | 254,409 | **1,210** | **254,409** | **255,619** |
| | **South Asian** | UK Biobank: Pulmonary broadly fibrotic | 57 | 570 | **57** | **570** | **627** | - | - | - | - | - | - |

| | | | | | | | | | | | | | |
|---|---|---|---|---|---|---|---|---|---|---|---|---|---|
| | **African** | Partanen (2022): Idiopathic pulmonary fibrosis | 169 | 8,368 | **169** | **8,368** | **8,537** | Partanen (2022): Idiopathic pulmonary fibrosis | 169 | 8,368 | **169** | **8,368** | **8,537** |
| **Reproductive** | **European** | UK Biobank: Reproductive broadly fibrotic | 7,813 | 78,130 | **7,813** | **78,130** | **85,943** | - | - | - | - | - | - |
| | **East Asian** | UK Biobank: Reproductive broadly fibrotic | 33 | 330 | **33** | **330** | **363** | - | - | - | - | - | - |
| | **South Asian** | UK Biobank: Reproductive broadly fibrotic | 190 | 1,900 | **190** | **1,900** | **2,090** | - | - | - | - | - | - |
| | **African** | UK Biobank: Reproductive broadly fibrotic | 148 | 1,480 | **148** | **1,480** | **1,628** | - | - | - | - | - | - |
| **Skeletal** | **European** | UK Biobank: Skeletal broadly fibrotic | 6,516 | 65,160 | **28,886** | **139,983** | **168,869** | - | - | - | - | - | - |
| | | Ishigaki (2022): Rheumatoid arthritis | 22,350 | 74,823 | | | | | | | | | |
| | **East Asian** | Ishigaki (2022): Rheumatoid arthritis | 11,025 | 162,608 | **11,025** | **162,608** | **173,633** | - | - | - | - | - | - |
| | **South Asian** | UK Biobank: Skeletal broadly fibrotic | 205 | 2,050 | **205** | **2,050** | **2,255** | - | - | - | - | - | - |
| | **African** | UK Biobank: Skeletal broadly fibrotic | 87 | 870 | **87** | **870** | **957** | - | - | - | - | - | - |
| **Skin** | **European** | UK Biobank: Skin broadly fibrotic | 6,593 | 65,930 | **20,285** | **104,863** | **125,148** | UK Biobank: Skin always fibrotic | 6,126 | 61,260 | **13,710** | **89,603** | **103,313** |
| | | Riesmeijer (2024): Dupuytren’s disease | 7,584 | 28,343 | | | | | | | | | |
| | | Julia (2018): Systemic lupus erythematosus) | 907 | 1,524 | | | | Riesmeijer (2024): Dupuytren’s disease | 7,584 | 28,343 | | | |
| | | Bentham (2015): Systemic lupus erythematosus | 5,201 | 9,066 | | | | | | | | | |
| | **East Asian** | Wang (2021): Systemic lupus erythematosus | 4,222 | 8,431 | **4,222** | **8,431** | **12,653** | - | - | - | - | - | - |
| | **South Asian** | UK Biobank: Skin broadly fibrotic | 93 | 930 | **93** | **930** | **1,023** | - | - | - | - | - | - |
| | **African** | UK Biobank: Skin broadly fibrotic | 66 | 660 | **66** | **660** | **726** | - | - | - | - | - | - |
| **Systemic** | **European** | UK Biobank: Systemic broadly fibrotic | 1,620 | 16,200 | **11,300** | **35,330** | **46,630** | - | - | - | - | - | - |
| | | López-Isac (2019): Systemic sclerosis | 9,095 | 17,584 | | | | | | | | | |
| | | Taylor (2017): Sjögren's syndrome | 585 | 1,546 | | | | | | | | | |

| | | | | | | | | | | | | | |
|---|---|---|---|---|---|---|---|---|---|---|---|---|---|
| | **East Asian** | Sakaue (2021): Sjögren's syndrome | 303 | 175,599 | **303** | **175,599** | **175,902** | - | - | - | - | - | - |
| | **South Asian** | UK Biobank: Systemic broadly fibrotic | 48 | 480 | **48** | **480** | **528** | - | - | - | - | - | - |
| | **African** | UK Biobank: Systemic broadly fibrotic | 42 | 420 | **42** | **420** | **462** | - | - | - | - | - | - |
| **Urinary** | **European** | UK Biobank: Urinary broadly fibrotic | 6,011 | 60,110 | **47,406** | **499,413** | **546,819** | - | - | - | - | - | - |
| | | Wuttke (2019): Chronic kidney disease | 41,395 | 439,303 | | | | | | | | | |
| | **East Asian** | Sakaue (2021): Polycystic Kidney Disease | 510 | 178,216 | **510** | **178,216** | **178,726** | - | - | - | - | - | - |
| | **South Asian** | UK Biobank: Urinary broadly fibrotic | 208 | 2,080 | **208** | **2,080** | **2,288** | - | - | - | - | - | - |
| | **African** | UK Biobank: Urinary broadly fibrotic | 161 | 1,610 | **161** | **1,610** | **1,771** | - | - | - | - | - | - |

#### Supplementary Table 4: Colocalisation results

Summary of $H_4$ results from colocalisation analyses. The organ the variant was most significant in is denoted by "ref" and all values are the colocalisation for the association signal between each organ and the "ref" organ. Values deemed to have colocalised ($H_4 \geq 80\%$) are coloured in grey. Organs with potential colocalisation ($H_4 \geq 50\%$) are shown in red text.

| Sentinel variant (nearest gene) | Organ-system | | | | | | | | | | | | Number of organs colocalised |
|---|---|---|---|---|---|---|---|---|---|---|---|---|---|
| | Biliary | Cardiovascular | Diabetes | Intestinal-pancreas | Liver | Lymphatic | Pulmonary | Reproductive | Skeletal | Skin | Systemic | Urinary | |
| rs7310615 (*SH2B3*) | 79.1% | ref | 97.9% | 0.3% | 3.5% | 7.8% | 84.0% | 0.8% | 99.4% | 0.9% | 71.3% | 97.7% | 5 |
| rs13246321 (*TPI1P2*) | 91.3% | 0.5% | 0.6% | 5.5% | 1.0% | 3.6% | 40.8% | 6.2% | 93.1% | 94.0% | ref | 0.5% | 4 |
| rs2476601 (*PTPN22*) | 7.7% | 84.5% | 99.9% | 0.6% | 2.0% | 52.7% | 1.6% | 1.3% | ref | 99.2% | 22.8% | 4.0% | 4 |
| rs8032939 (*RASGRP1*) | 84.1% | 96.1% | 95.3% | 0.2% | 2.4% | 2.1% | 33.0% | 0.8% | ref | 2.7% | 64.7% | 0.0% | 4 |
| rs780093 (*GCKR*) | 1.0% | 3.3% | ref | 0.0% | 98.6% | 2.8% | 0.4% | 0.5% | 0.5% | 0.6% | 0.7% | 99.6% | 3 |
| rs2736340 (*BLK*) | 0.8% | 0.3% | 0.3% | 0.4% | 1.1% | 3.3% | 1.0% | 1.4% | 98.4% | 98.8% | ref | 3.7% | 3 |
| rs6679356 (*IL12RB2*) | ref | 0.5% | 0.5% | 0.2% | 0.7% | 14.8% | 2.3% | 0.7% | 99.1% | 99.2% | 0.4% | 0.5% | 3 |
| rs11089637 (*UBE2L3*) | 3.6% | 0.6% | 0.4% | 2.5% | 1.4% | 3.3% | 1.9% | 1.0% | ref | 90.6% | 94.1% | 0.6% | 3 |
| rs772106113 (*IL12A-AS1*) | ref | 0.4% | 0.4% | 0.3% | 0.7% | 3.7% | 0.9% | 1.7% | 74.3% | 42.6% | 95.5% | 0.5% | 2 |
| rs2069235 (*SYNGR1*) | 98.2% | 0.4% | 0.4% | 0.1% | 1.5% | 3.2% | 1.3% | 1.1% | ref | 0.9% | 1.2% | 0.7% | 2 |
| rs4853459 (*STAT4*) | 0.0% | 0.5% | 4.1% | 0.5% | 1.0% | 3.3% | 4.4% | 2.7% | 93.9% | ref | 0.0% | 0.5% | 2 |
| rs113897057 (*GSDMB*) | ref | 2.6% | 0.4% | 0.3% | 1.0% | 2.8% | 1.2% | 0.8% | 49.4% | 1.1% | 95.1% | 0.5% | 2 |
| rs769449 (*APOE*) | 33.8% | ref | 0.0% | 2.6% | 96.4% | 6.1% | 68.8% | 0.6% | 0.8% | 0.9% | 3.4% | 1.0% | 2 |
| rs643434 (*ABO*) | 3.4% | 50.6% | 94.8% | ref | 1.1% | 2.8% | 1.0% | 38.0% | 0.5% | 0.7% | 6.0% | 0.7% | 2 |
| rs7117261 (*CXCR5*) | ref | 0.6% | 0.6% | 1.3% | 6.0% | 4.4% | 1.1% | 0.9% | 97.8% | 1.7% | 9.0% | 0.4% | 2 |
| rs10401969 (*SUGP1*) | 1.6% | 1.0% | 96.6% | 0.6% | ref | 23.2% | 1.6% | 1.6% | 0.1% | 0.0% | 6.7% | 2.4% | 2 |
| rs13268508 (*HSF1*) | 1.1% | 0.4% | 97.6% | 0.7% | 1.0% | 7.9% | 5.9% | 0.9% | 0.5% | ref | 1.5% | 1.2% | 2 |
| rs11982901 (*IRF5*) | ref | 0.4% | 0.4% | 2.4% | 4.1% | 2.8% | 19.1% | 3.4% | 96.1% | 0.0% | 0.3% | 0.5% | 2 |
| rs1458038 (*FGF5*) | 0.7% | 98.2% | 0.8% | 4.0% | 1.0% | 3.0% | 1.4% | 0.9% | 0.7% | 1.2% | 2.3% | ref | 2 |
| rs16879645 (*ELMO1*) | ref | 20.8% | 0.6% | 8.2% | 1.1% | 7.9% | 1.4% | 1.2% | 95.8% | 0.0% | 38.8% | 4.6% | 2 |
| rs28601761 (*TRIB1AL*) | 0.8% | 82.3% | 1.3% | 0.3% | ref | 2.7% | 6.9% | 0.7% | 0.6% | 1.7% | 6.9% | 6.5% | 2 |
| rs35382826 (*LINC03004*) | 98.8% | 2.3% | 0.0% | 13.8% | 0.9% | 2.9% | 6.7% | 1.7% | ref | 0.1% | 1.4% | 2.3% | 2 |
| rs3792783 (*TNIP1*) | 19.2% | 0.5% | 1.4% | 1.1% | 1.4% | 5.4% | 3.8% | 1.1% | 12.0% | 97.5% | ref | 1.0% | 2 |
| rs4782661 (*CRISPLD2*) | 0.8% | 2.5% | 0.4% | ref | 0.7% | 2.2% | 1.1% | 0.8% | 0.6% | 11.2% | 1.3% | 97.0% | 2 |
| rs765299456 (*ABCG5*) | ref | 89.1% | 0.0% | 0.9% | 1.5% | 3.7% | 2.6% | 1.6% | 1.0% | 2.3% | 4.0% | 0.6% | 2 |
| rs7528321 (*PRDX6*) | 2.5% | 0.4% | 0.2% | 0.8% | 1.2% | 4.5% | 1.0% | 1.0% | 90.8% | 18.7% | ref | 0.9% | 2 |
| rs881858 (*VEGFA*) | 1.7% | 0.7% | 95.5% | 0.2% | 3.0% | 20.5% | 0.5% | 0.8% | 0.5% | 0.0% | 1.5% | ref | 2 |
| rs2297550 (*IKBKE*) | 2.3% | 0.5% | 98.6% | 0.5% | 1.7% | 7.7% | 0.2% | 4.7% | 0.0% | ref | 2.1% | 1.1% | 2 |
| rs2293370 (*TIMMDC1*) | ref | 0.8% | 0.4% | 0.6% | 0.9% | 3.6% | 2.4% | 0.9% | 7.3% | 3.4% | 98.0% | 0.6% | 2 |

#### Supplementary Table 5: Summary of functional follow-up analyses

See Excel file titled Supplementary_Table_5_Functional_follow_up_of_signals_of_interest.xlsx. The sheet contains the following columns:

- **Type**: Type of study (novel & replicated signal or colocalised ≥ 3 organs signal)
- **Organ**: Organ where the signal was showed the strongest association
- **rsid**: SNP identifier from dbSNP
- **chr**: Chromosome
- **position_b37**: Genomic position in GRCh37/hg19
- **position_b38**: Genomic position in GRCh38/hg38
- **reference_allele**: Reference allele
- **effect_allele**: Effect allele
- **eaf**: Frequency of the effect allele
- **beta**: Effect size
- **se**: Standard error
- **p**: P-value
- **location_vep**: Variant location annotated by VEP (Variant Effect Predictor)
- **nearest_gene_vep**: Nearest gene to the variant according to VEP
- **SIFT_vep**: Functional prediction from SIFT (tolerated or deleterious)
- **PolyPhen_vep**: Functional prediction from PolyPhen (benign, possibly damaging, probably damaging)
- **CADD_PHRED_vep**: CADD score predicting deleteriousness
- **Fine-mapping (PIP)**: Fine-mapping results (posterior inclusion probability)
- **RegulomeDB_(score)**: RegulomeDB score
- **epigenMark**: Epigenetic marks associated with the variant
- **eQTL_GTEx_V8_coloc**: Colocalisation with gene expression QTL from GTEx V8
- **sQTL_GTEx_V8_coloc**: Colocalisation with splicing QTL from GTEx V8
- **CISpQTLs**: Variants affecting protein levels in cis (nearby genes)
- **CISpQTLs_coloc**: Colocalisation with cis protein levels QTL
- **mouseKO_genes**: Genes studied in mouse knockout models
- **rareHPO_genes**: Genes linked to rare phenotypes in Orphanet
- **AZ_Gene-level-results**: Gene-level results from AstraZeneca PheWAS portal
- **AZ_Variant-level-results**: Variant-level results from AstraZeneca PheWAS portal

#### Supplementary Table 6: Sample sizes in All of Us replication analyses

| | Broadly fibrotic | | | Always fibrotic | | |
|---|---|---|---|---|---|---|
| | Cases | Controls | Total | Cases | Controls | Total |
| **Biliary** | 2,171 | 21,710 | 23,881 | 213 | 2,130 | 2,343 |
| **Cardiovascular** | 16,524 | 147,872 | 164,396 | - | - | - |
| **Diabetes** | 19, 210 | 145,186 | 164,396 | - | - | - |
| **Intestinal-pancreatic** | 5,799 | 57,990 | 63,789 | - | - | - |
| **Liver** | 3,955 | 39,550 | 43,505 | 1,939 | 19,390 | 21,329 |
| **Lymphatic** | 213 | 2,130 | 2,343 | - | - | - |
| **Pulmonary** | 1,480 | 14,800 | 16,280 | 1,300 | 13,000 | 14,300 |
| **Reproductive** | 1,428 | 14,280 | 15,708 | - | - | - |
| **Skeletal** | 4,676 | 46,760 | 51,436 | - | - | - |
| **Skin** | 5,755 | 57,550 | 63,305 | 4,704 | 47,040 | 51,744 |
| **Systemic** | 2,906 | 29,060 | 31,966 | - | - | - |
| **Urinary** | 8,680 | 86,800 | 95,480 | - | - | - |

**Supplementary Table 7: Heritability and genetic correlations between 'always fibrotic' and 'broadly fibrotic' case-definitions**

| | Heritability using 'always fibrotic' case definition (Heritability [95% CI]) | Heritability using 'broadly fibrotic' case definition (Heritability [95% CI]) | Genetic correlation between 'always fibrotic' and 'broadly fibrotic' case definitions (Correlation [95% CI]) |
|---|---|---|---|
| **Biliary** | 5.8% [0%, 15.4%] | 4.1% [2.3%, 5.9%] | 0.67 [0.13, 1] |
| **Liver** | 4.5% [0%, 9.9%] | 4.4% [2.3%, 6.5%] | 0.90 [0.52, 1] |
| **Pulmonary** | 6.0% [0.6%, 11.5%] | 3.2% [0%, 6.5%] | 0.89 [0.72, 1] |
| **Skin** | 6.0% [3.4%, 8.5%] | 5.1% [2.9%, 7.3%] | 0.99 [0.98, 1] |

#### Supplementary Figures

##### Supplementary Figure 1: Flow chart showing selection of previously published studies for inclusion in meta-analyses

Note, one study may have performed multiple analyses and therefore contribute more than one set of summary statistics.

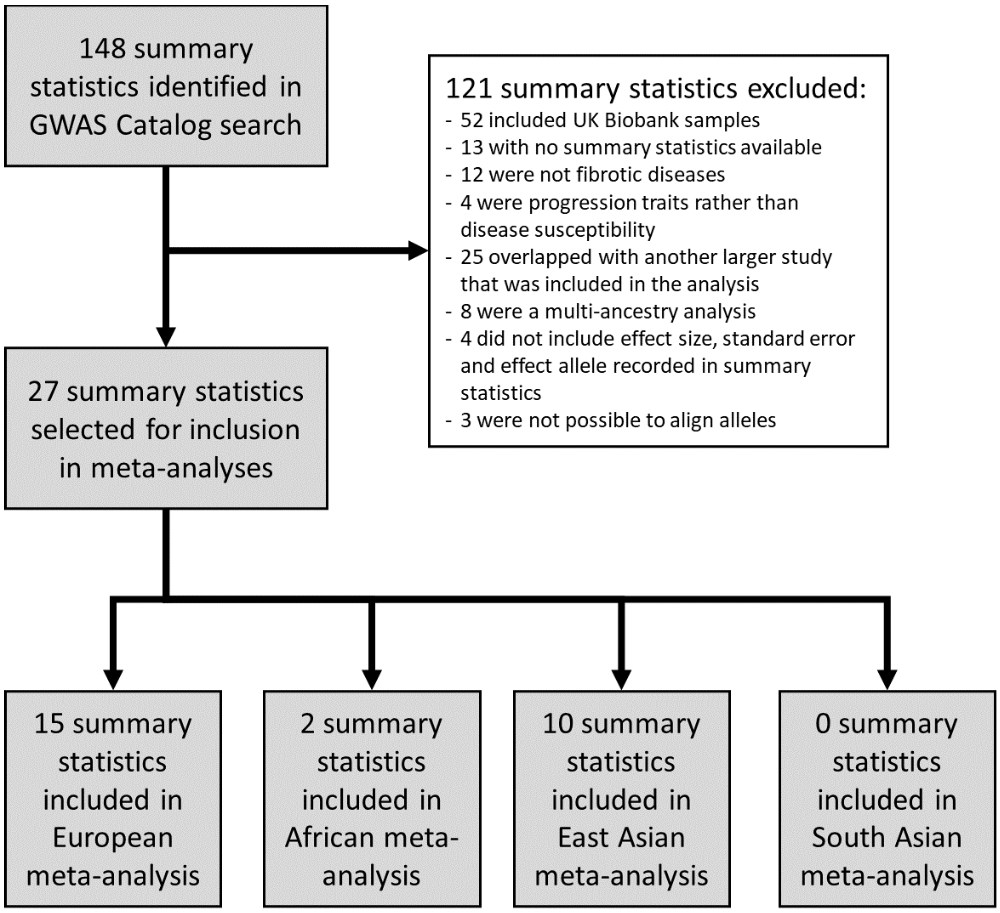

**Supplementary Figure 2: Manhattan plots for European genome-wide meta-analyses**

Each point represents a single genetic variant with position on the x axis and $-\log_{10}$(p-value) on the y axis. The red line denotes the genome-wide significance threshold of $5\times10^{-8}$.

**a) Biliary (always fibrotic)**

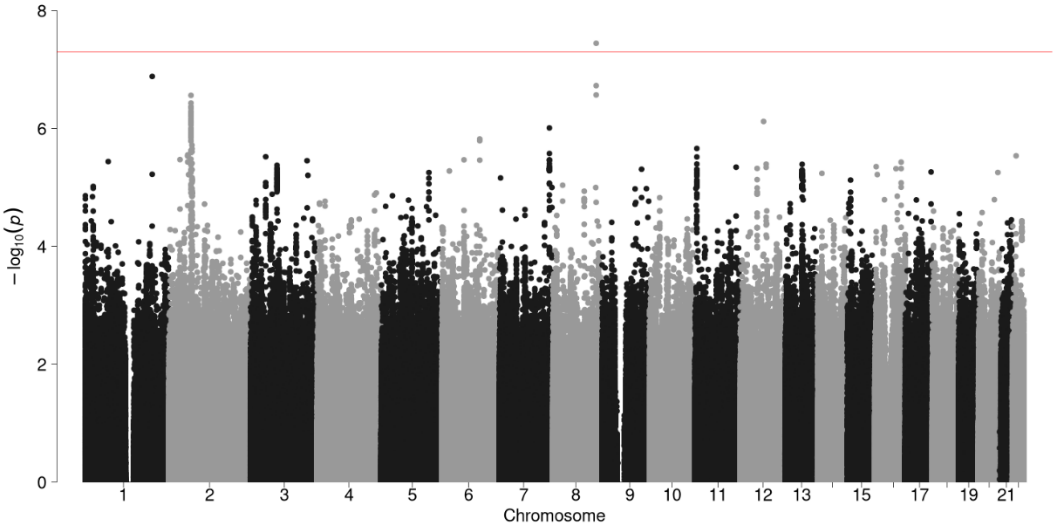

**b) Biliary (broadly fibrotic)**

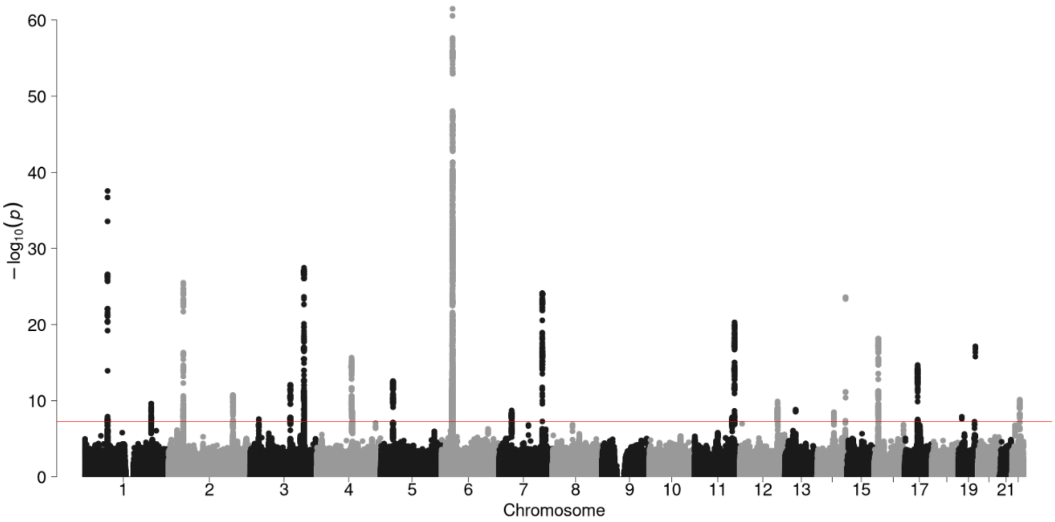

**c) Cardiovascular (broadly fibrotic)**

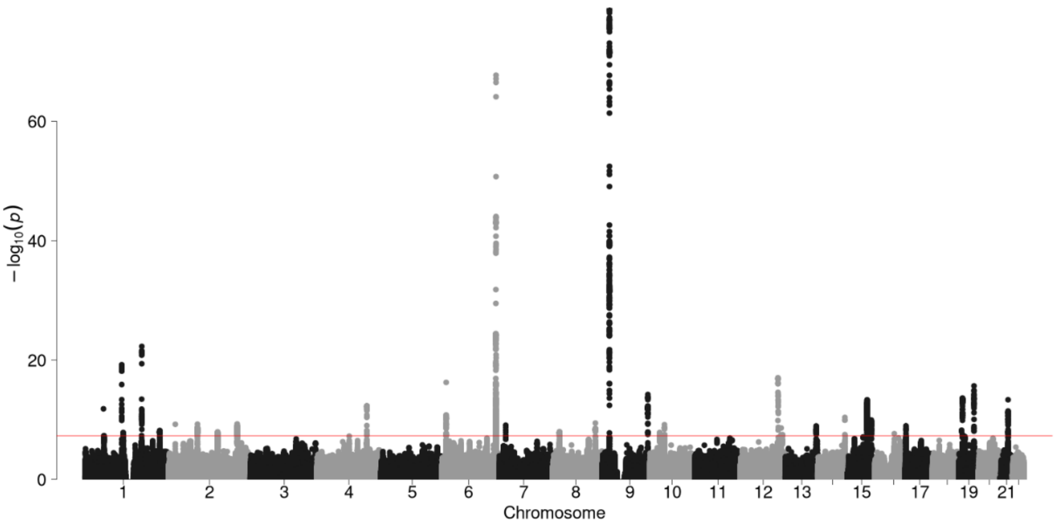

**d) Diabetes (broadly fibrotic)**

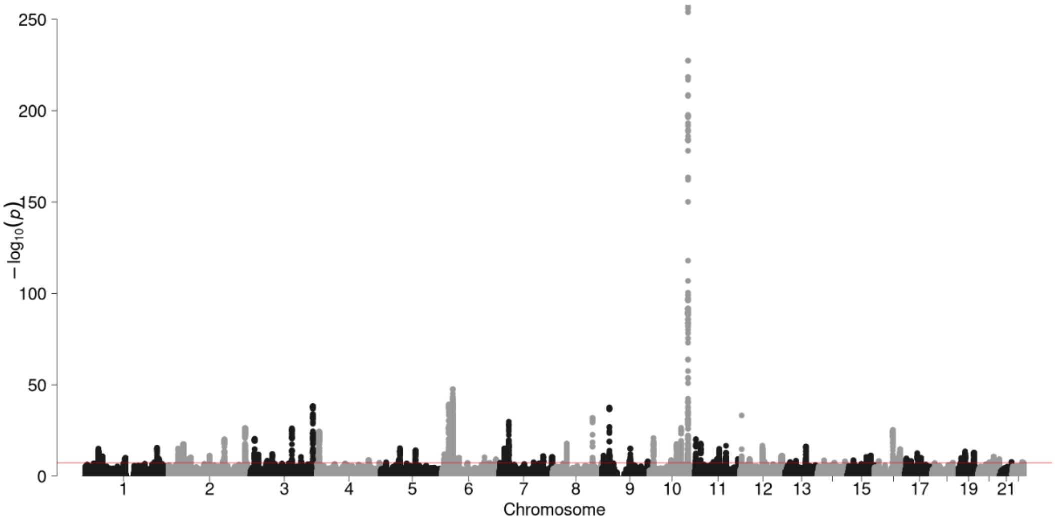

**e) Intestinal-pancreatic (broadly fibrotic)**

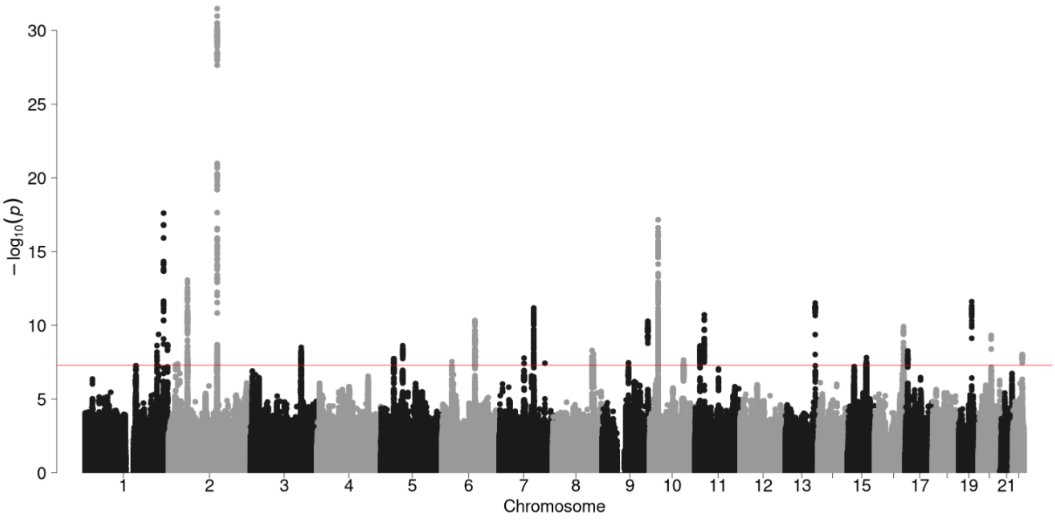

**f) Liver (always fibrotic)**

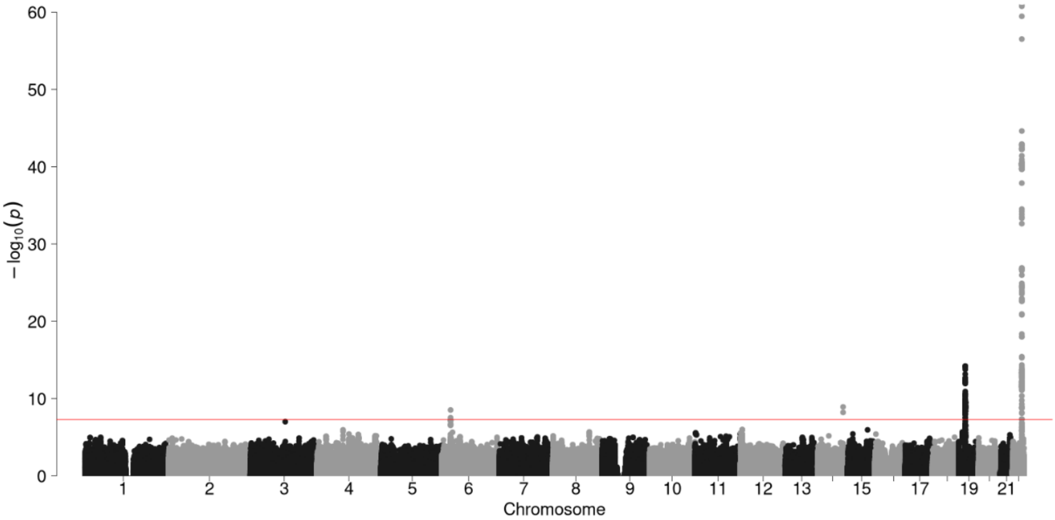

**g) Liver (broadly fibrotic)**

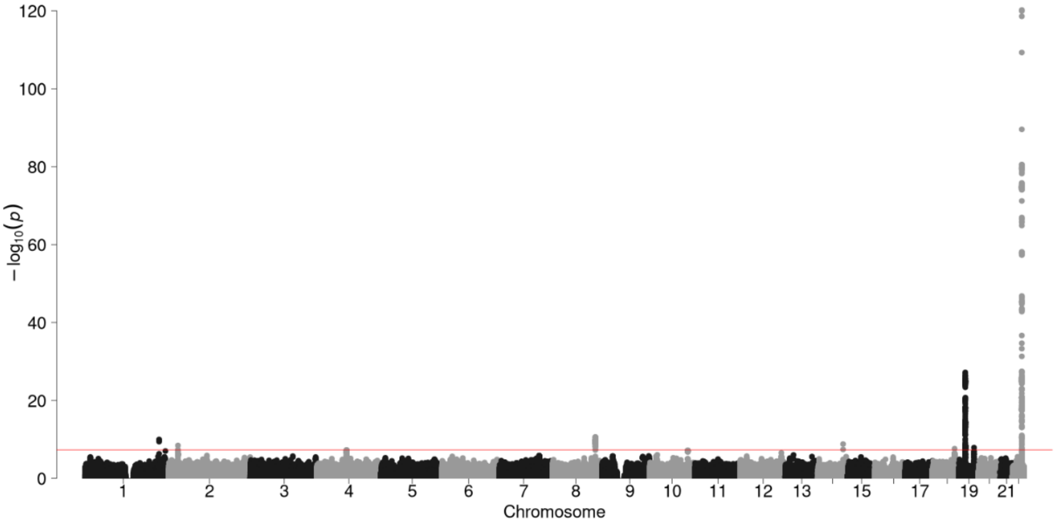

**h) Lymphatic (broadly fibrotic)**

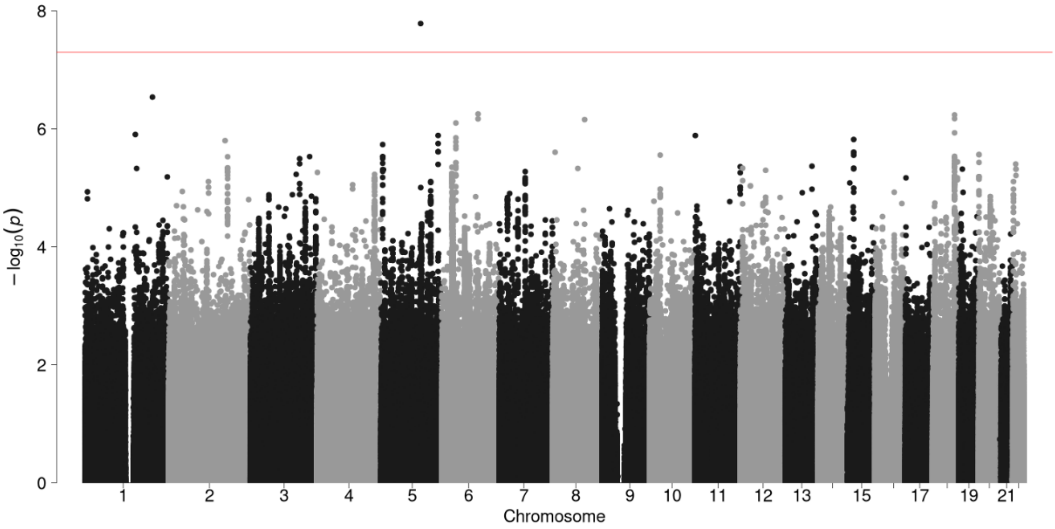

**i) Pulmonary (always fibrotic)**

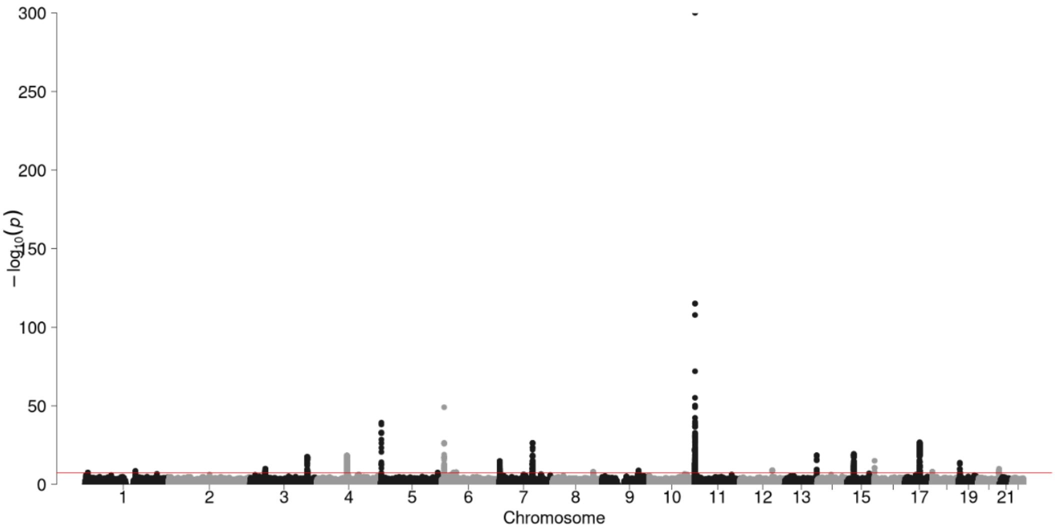

**j) Pulmonary (broadly fibrotic)**

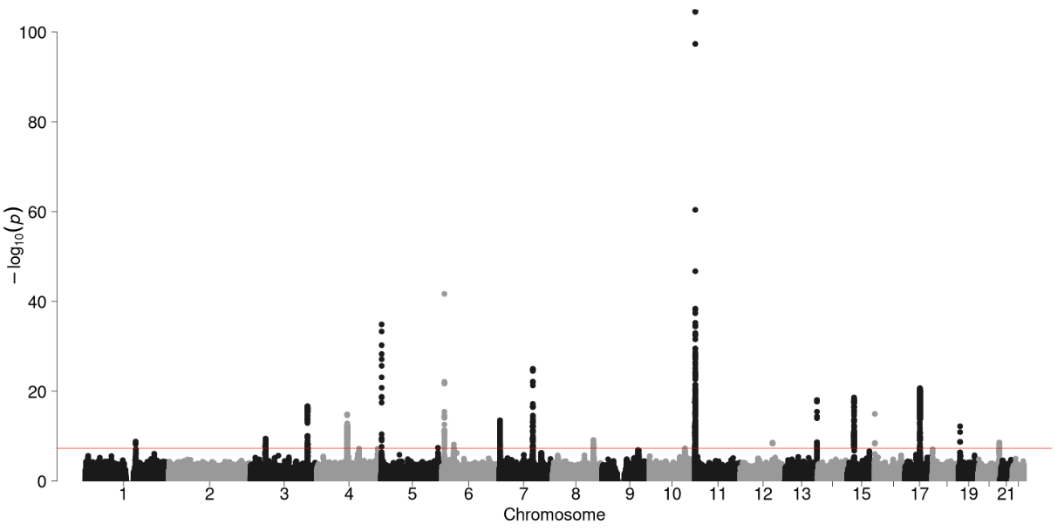

**k) Reproductive (broadly fibrotic)**

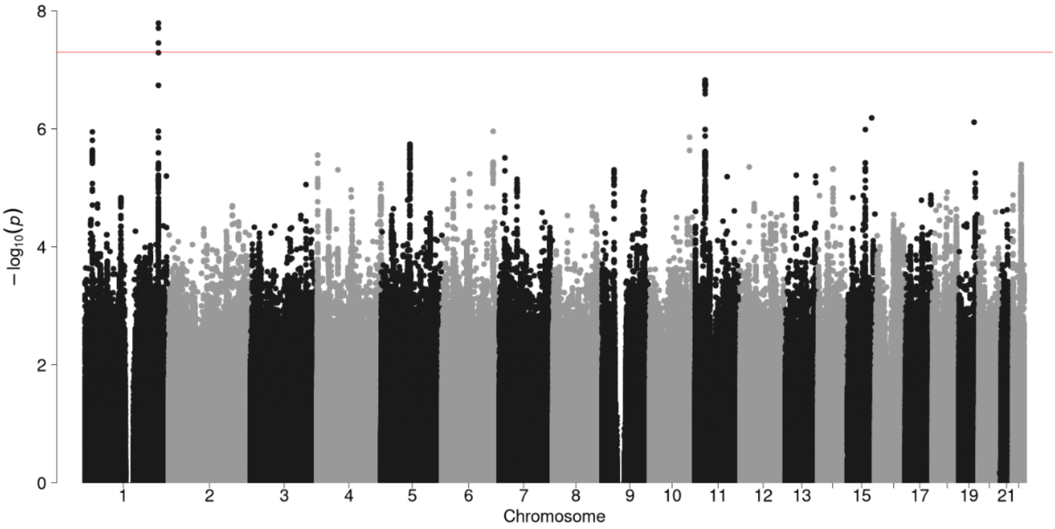

**l) Skeletal (broadly fibrotic)**

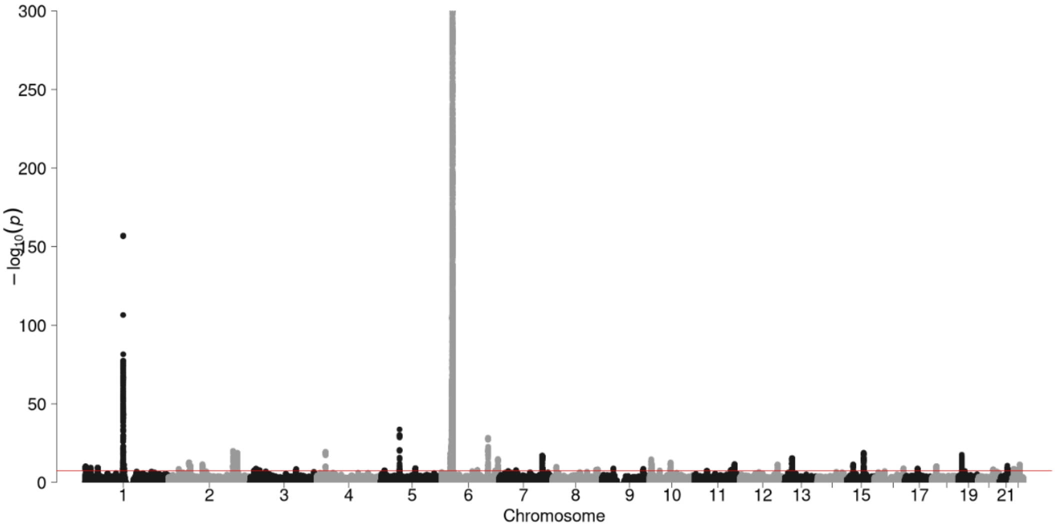

**m) Skin (always fibrotic)**

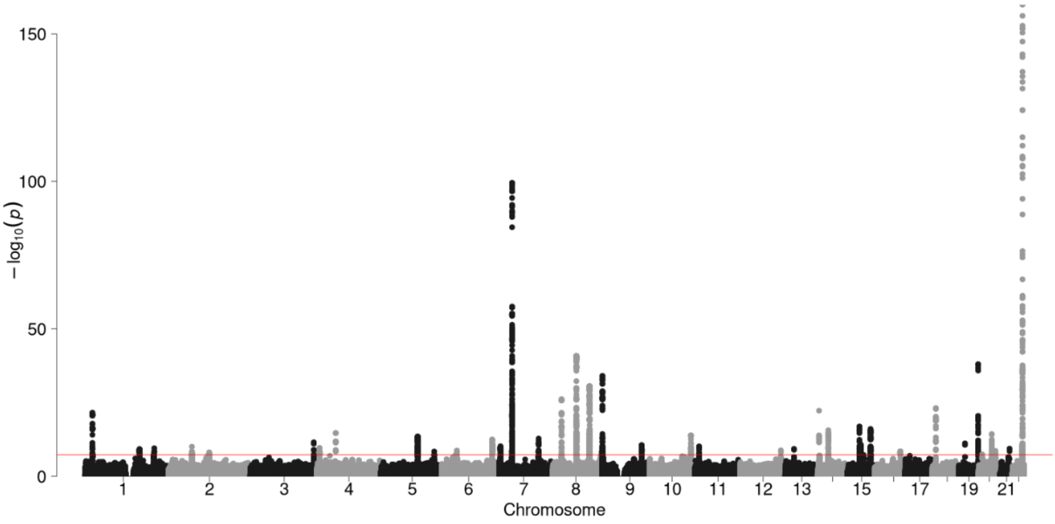

**n) Skin (broadly fibrotic)**

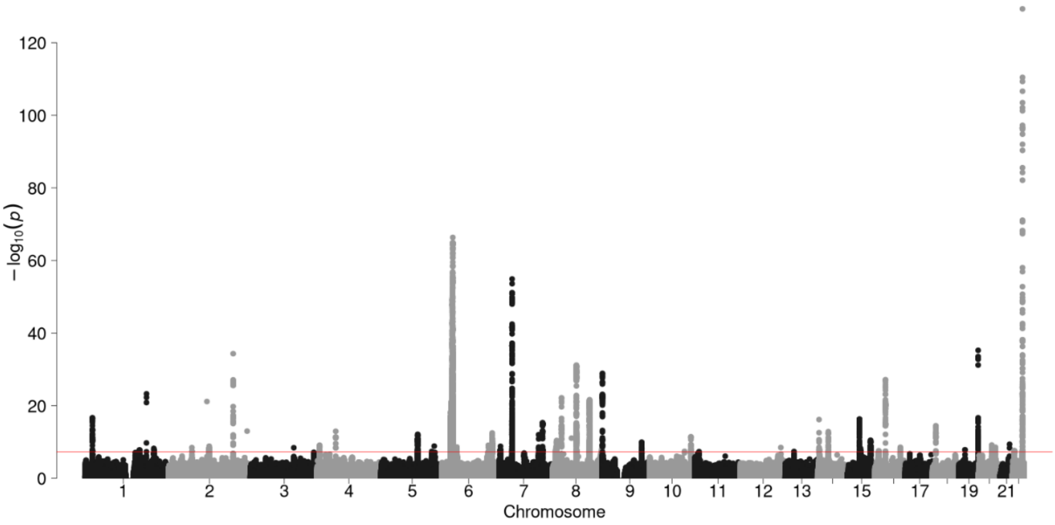

**o) Systemic (broadly fibrotic)**

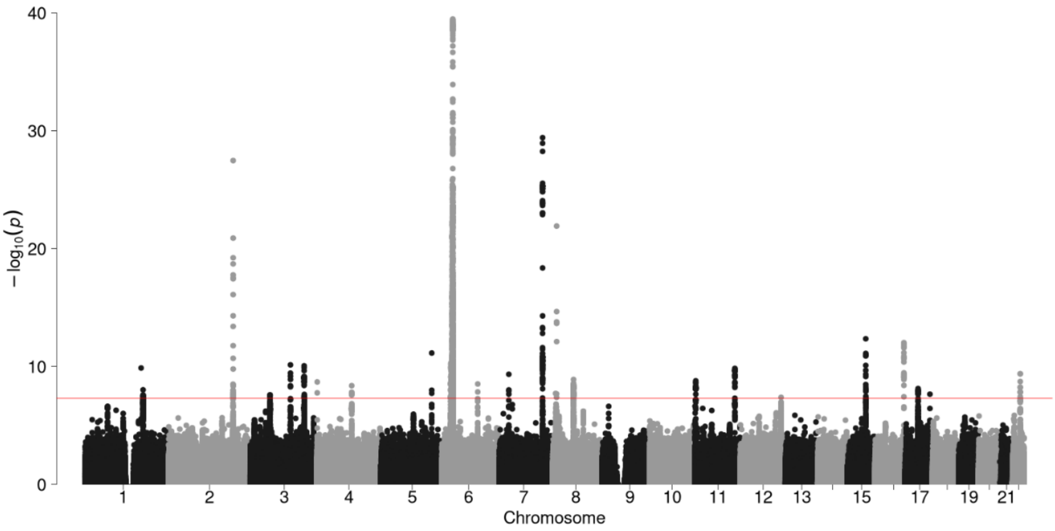

**p) Urinary (broadly fibrotic)**

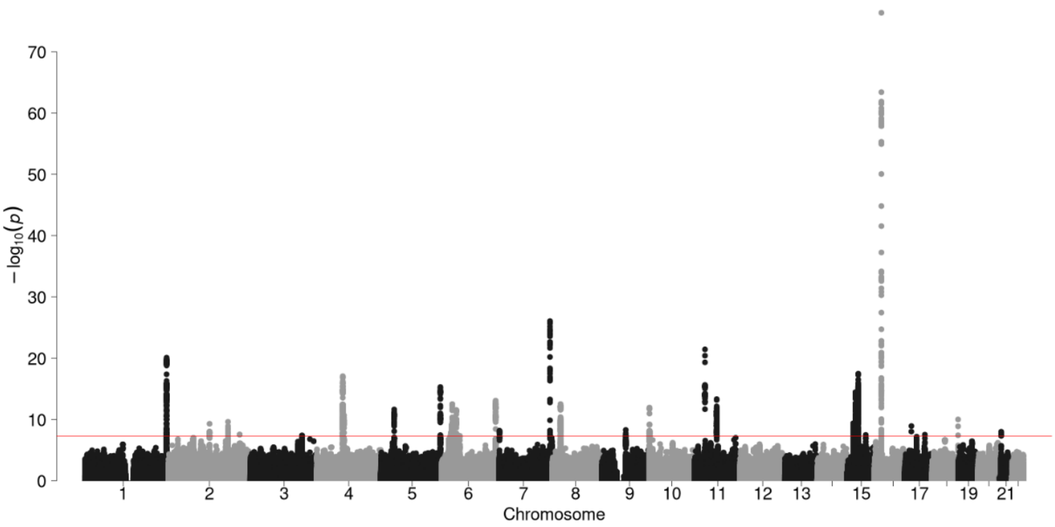

#### Supplementary Figure 3: QQ plots for European genome-wide meta-analyses

##### a) Biliary (always fibrotic)

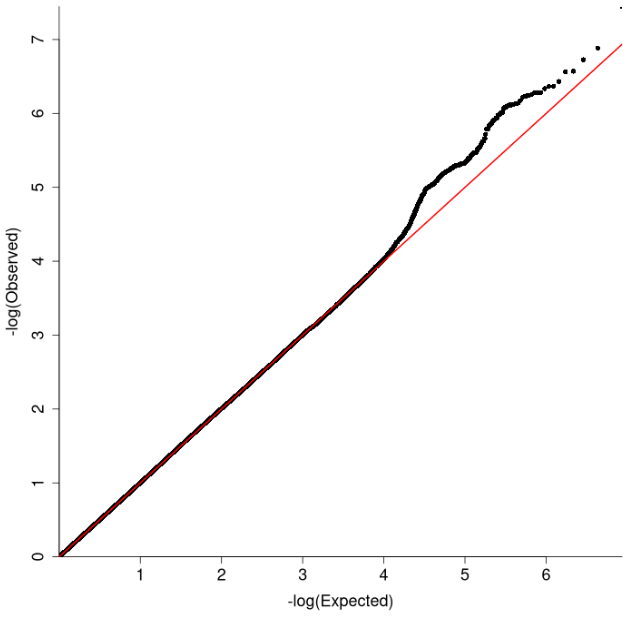

##### b) Biliary (broadly fibrotic)

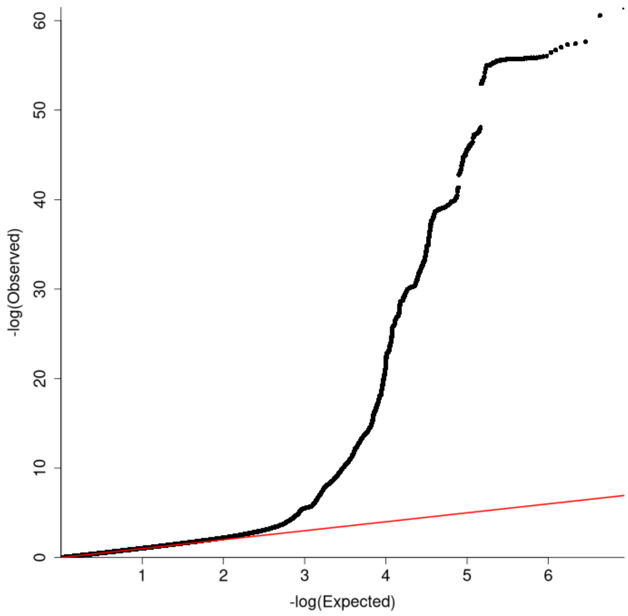

##### c) Cardiovascular (broadly fibrotic)

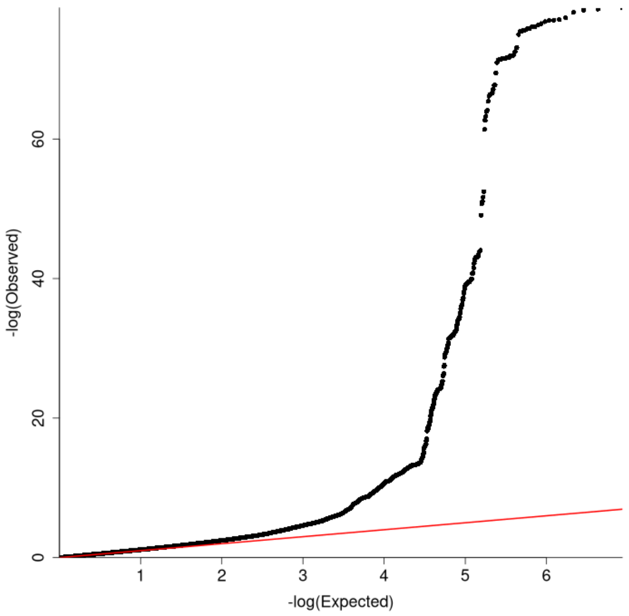

##### d) Diabetes (broadly fibrotic)

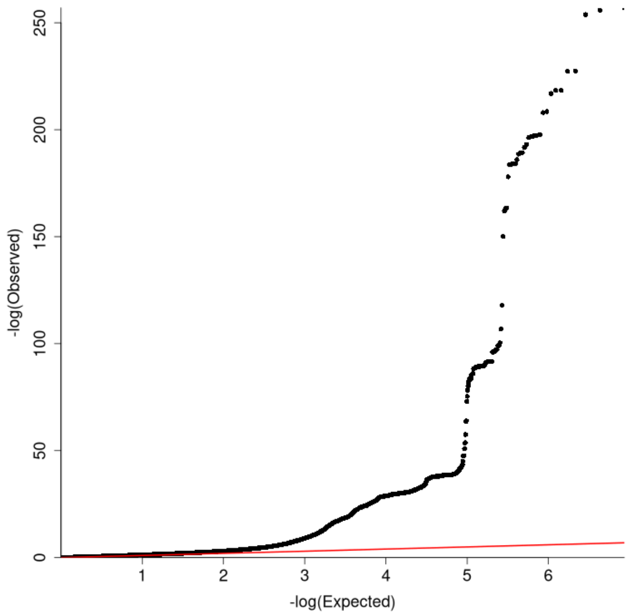

e) Intestinal-pancreatic (broadly fibrotic)

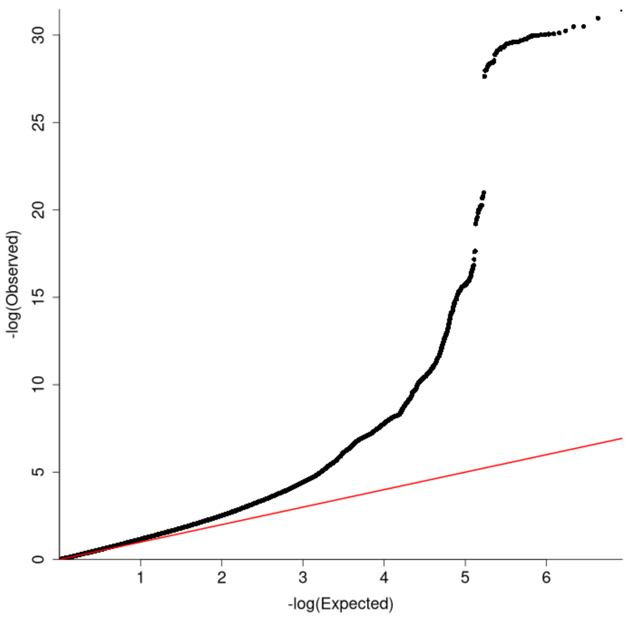

f) Liver (always fibrotic)

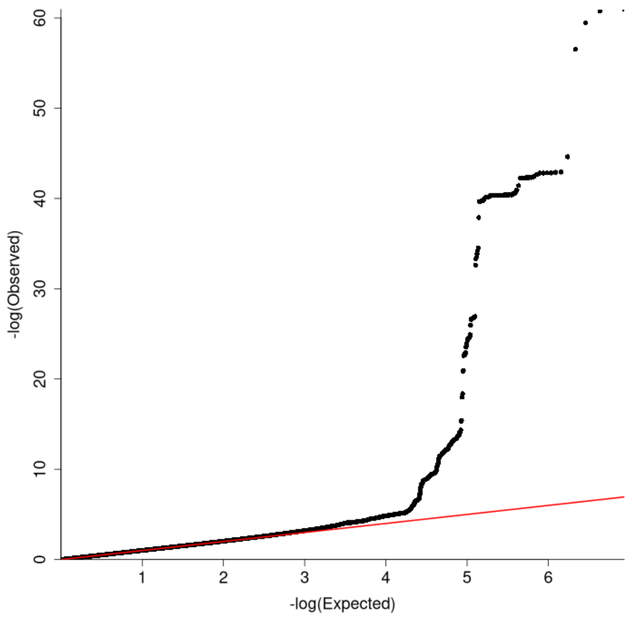

g) Liver (broadly fibrotic)

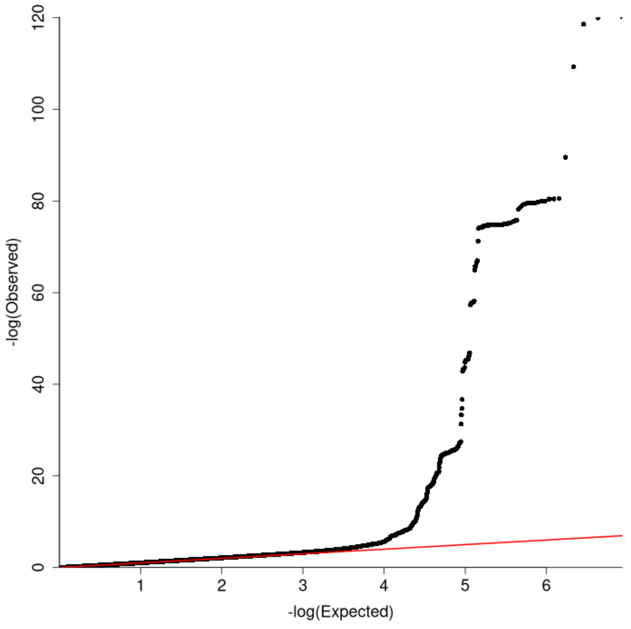

h) Lymphatic (broadly fibrotic)

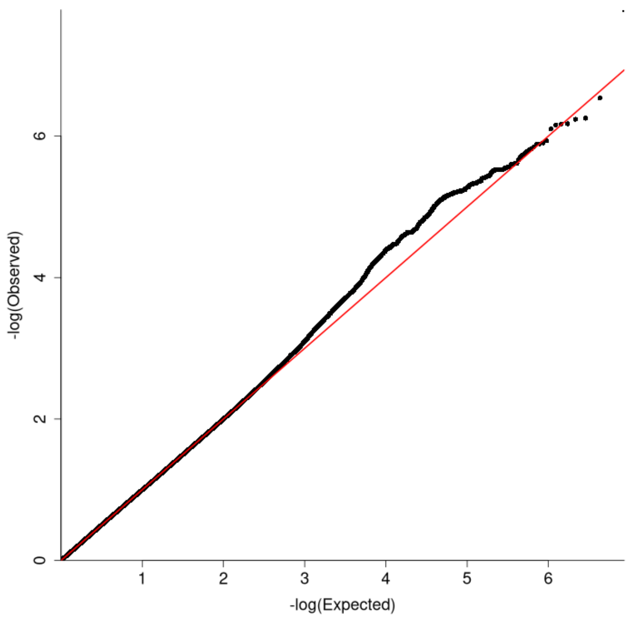

**i) Pulmonary (always fibrotic)**

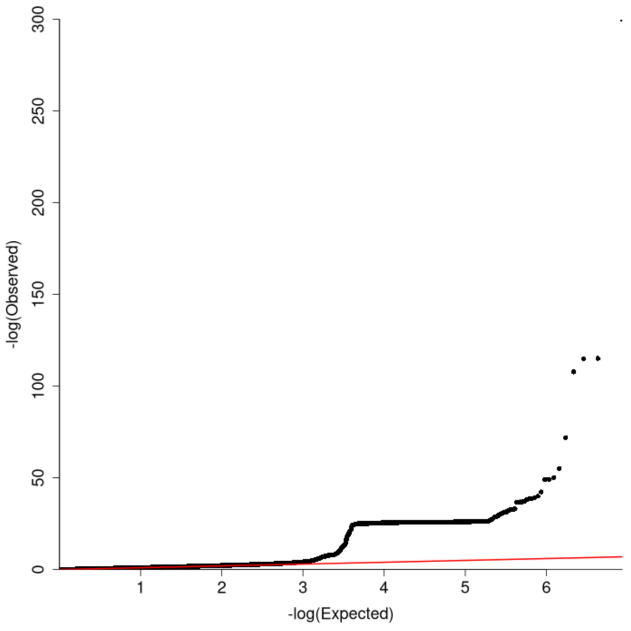

**j) Pulmonary (broadly fibrotic)**

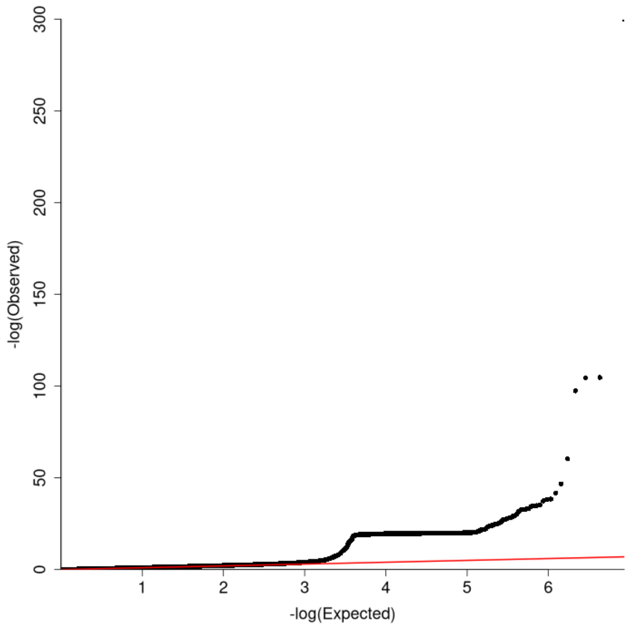

**k) Reproductive (broadly fibrotic)**

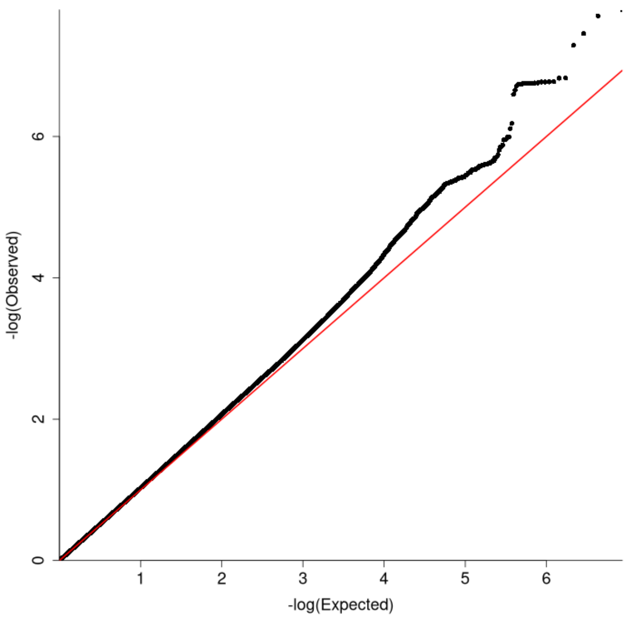

**l) Skeletal (broadly fibrotic)**

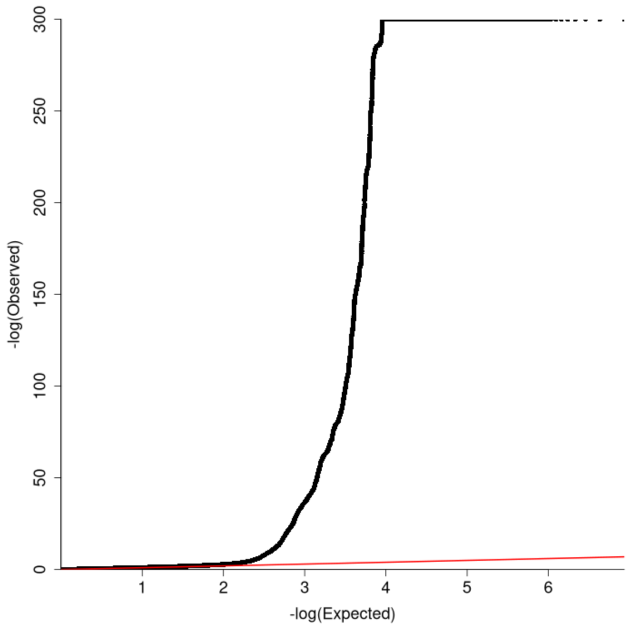

**m) Skin (always fibrotic)**

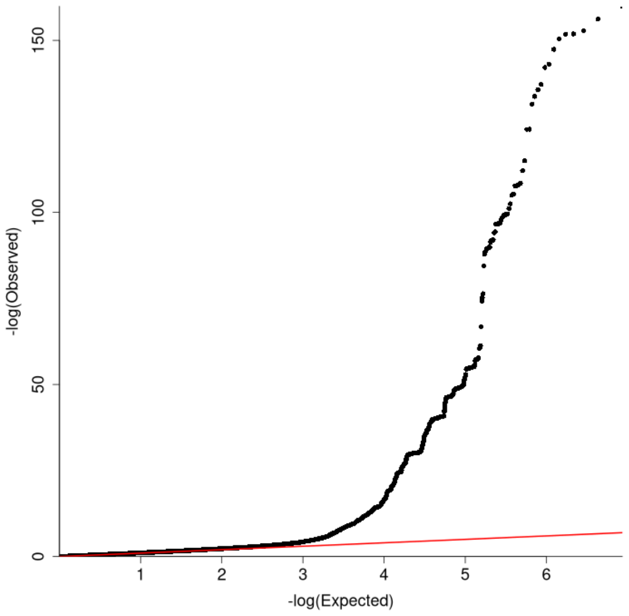

**n) Skin (broadly fibrotic)**

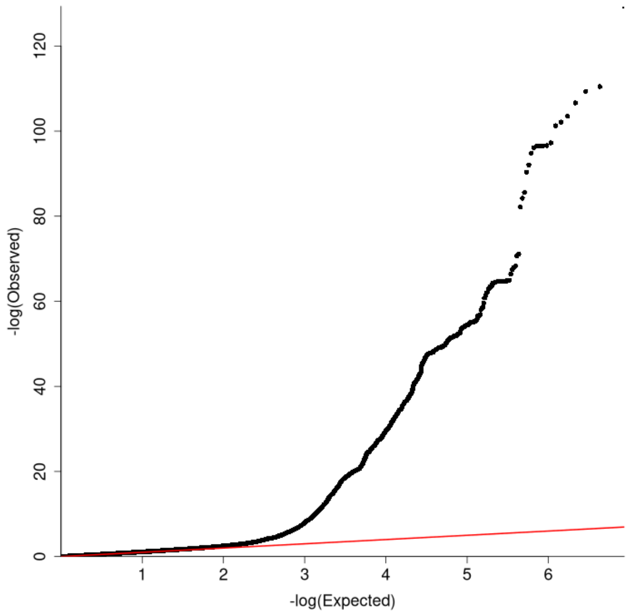

**o) Systemic (broadly fibrotic)**

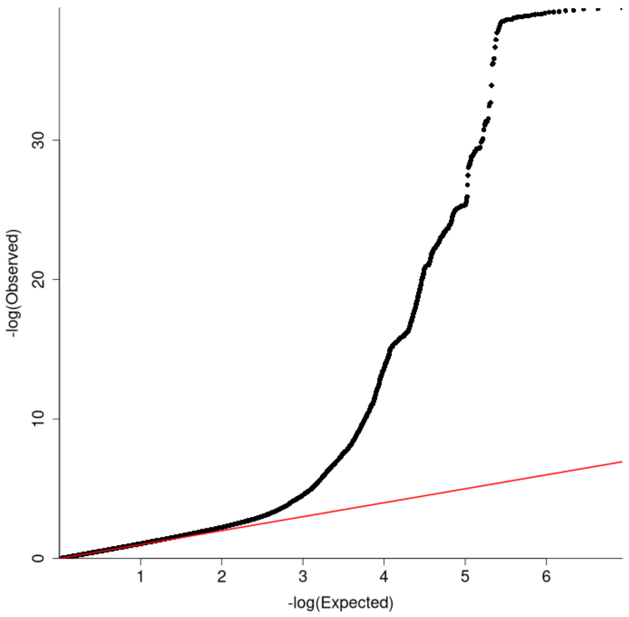

**p) Urinary (broadly fibrotic)**

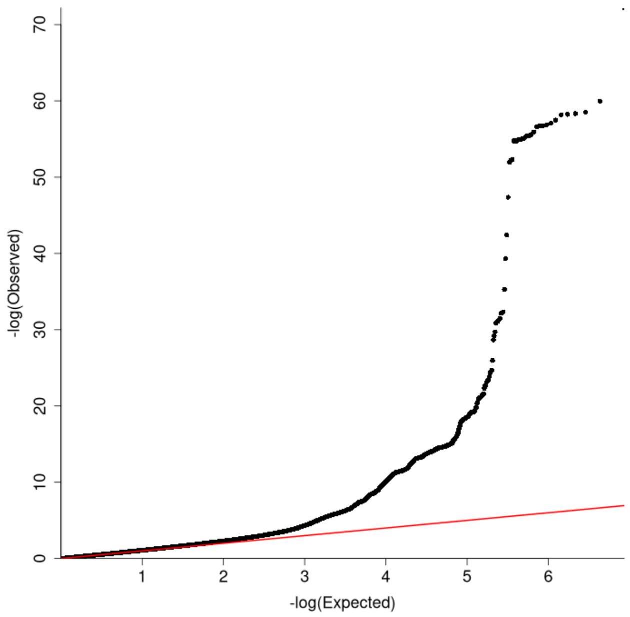

**Supplementary Figure 4: Region plots for genetic loci that colocalised across three or more organs**

Each point represents a variant with chromosomal position (GRCh37) on x axis and $-\log_{10}$(p value) on y axis. The sentinel variant is coloured purple with other variants coloured by strength of LD with the sentinel variant based on the 1000 genomes EUR population. Recombination rates shown by the blue lines and positions of genes shown at bottom. $H_4$ is the probability of a shared causal variant between that organ and the organ with the strongest association at that locus. Organs with $H_4 \geq 80\%$ were deemed to have colocalised.

**a) rs7310615 (*SH2B3*)**

a) Cardiovascular

b) Skeletal ($H_4$=99.4%)

c) Diabetes ($H_4$=97.9%)

d) Urinary ($H_4$=97.7%)

e) Pulmonary ($H_4$=84.0%)

f) Biliary ($H_4$=79.1%)

g) Systemic ($H_4$=71.3%)

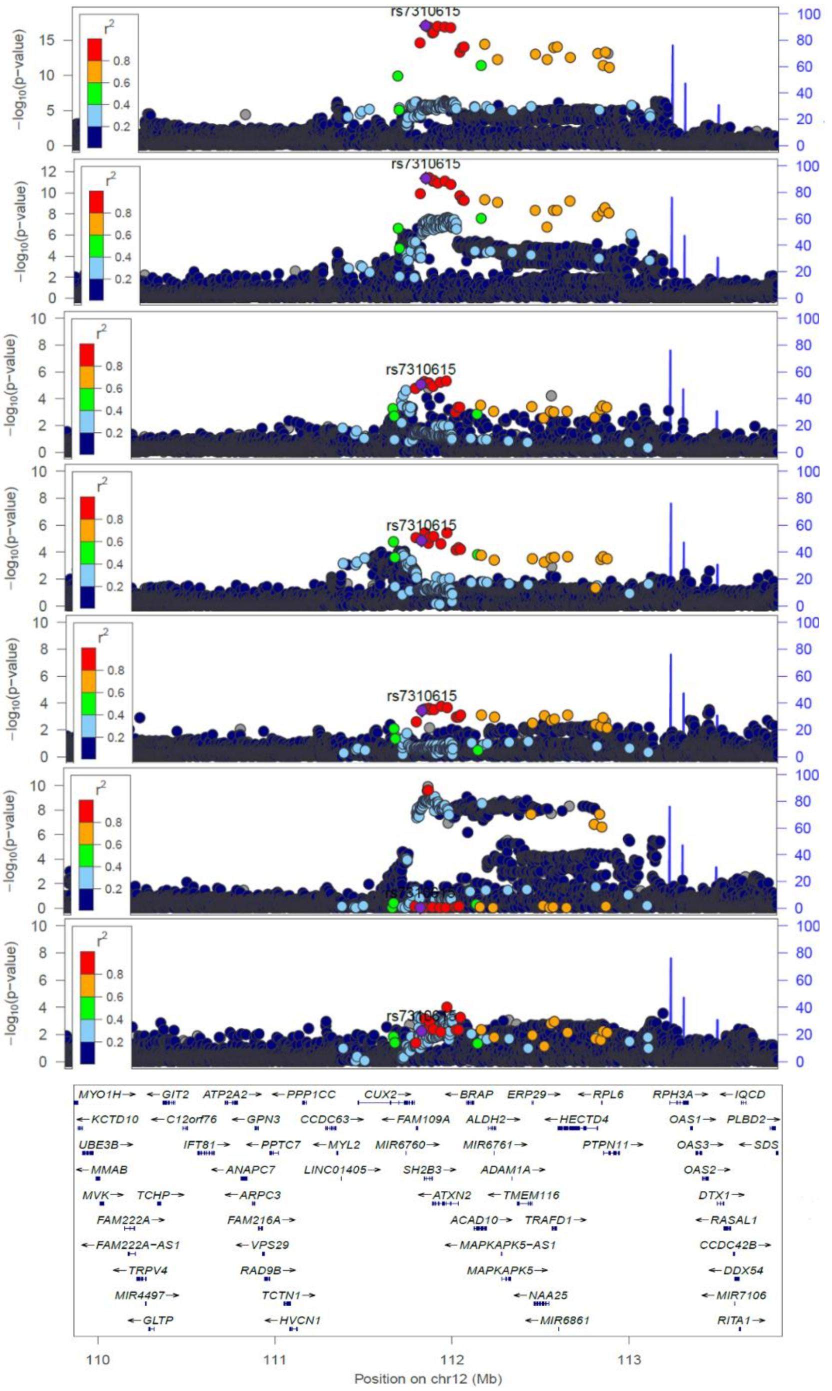

**b) rs6679356 (*IL12RB2*)**

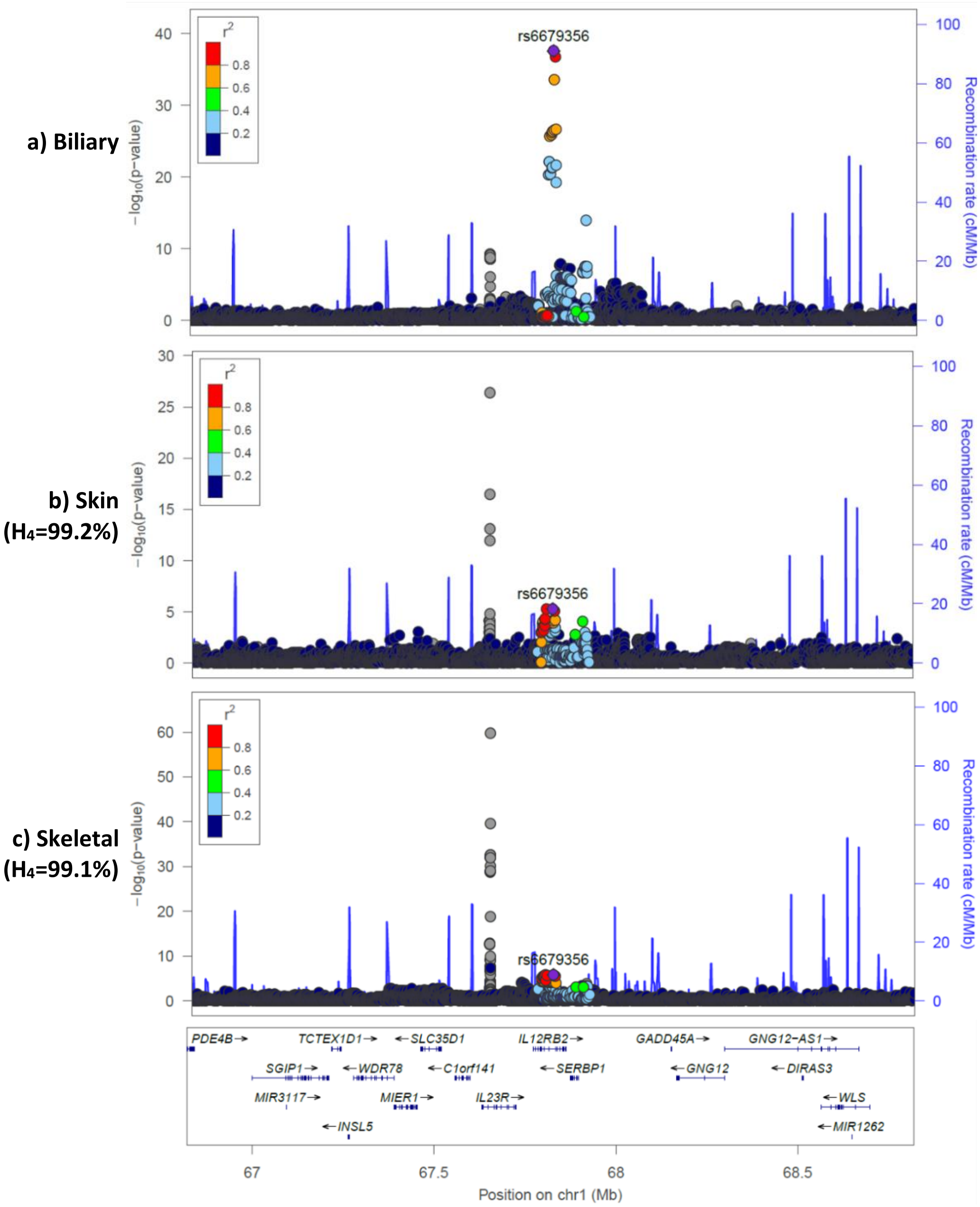

c) rs2476601 (*PTPN22*)

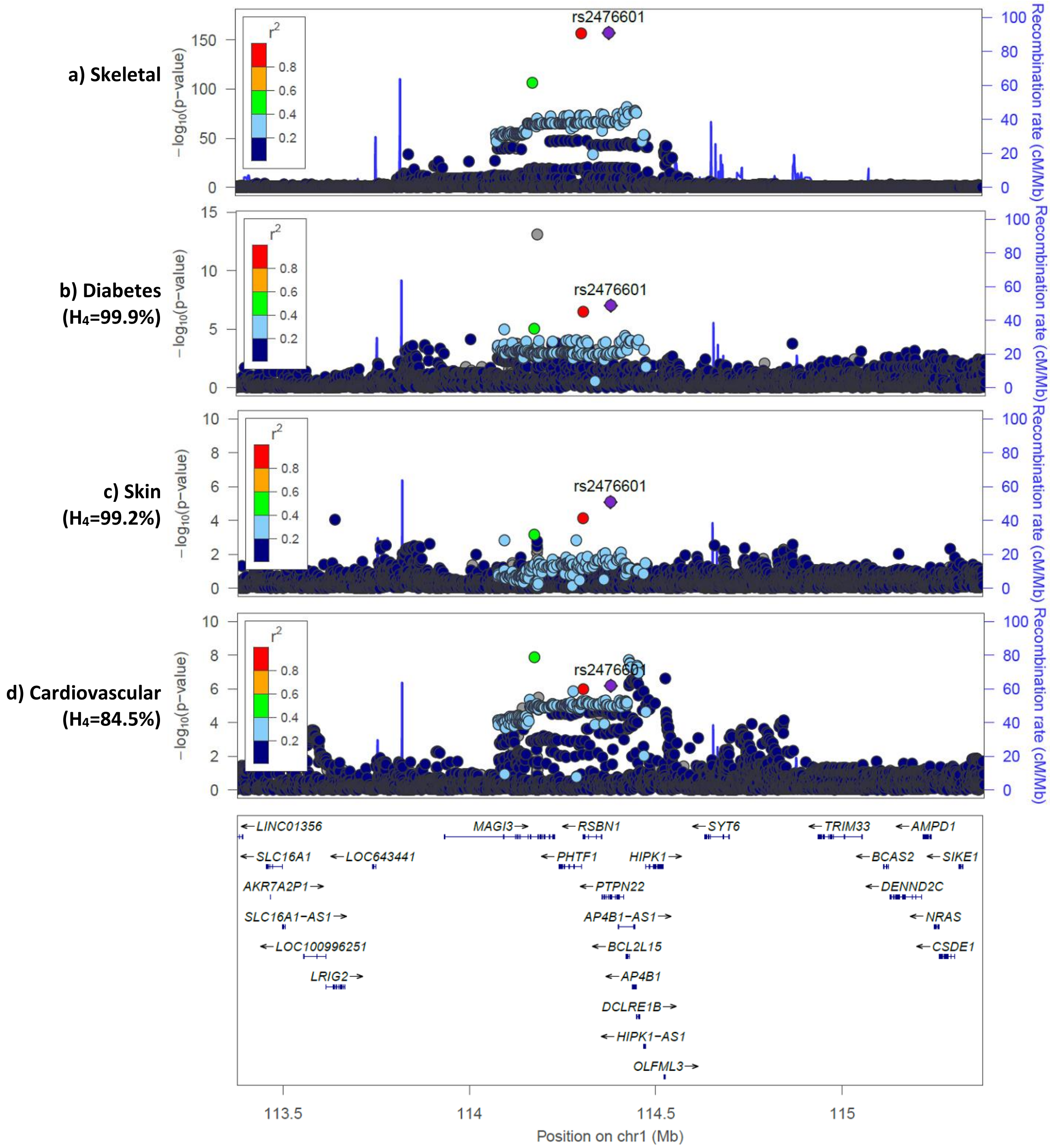

**d) rs780093 (*GCKR*)**

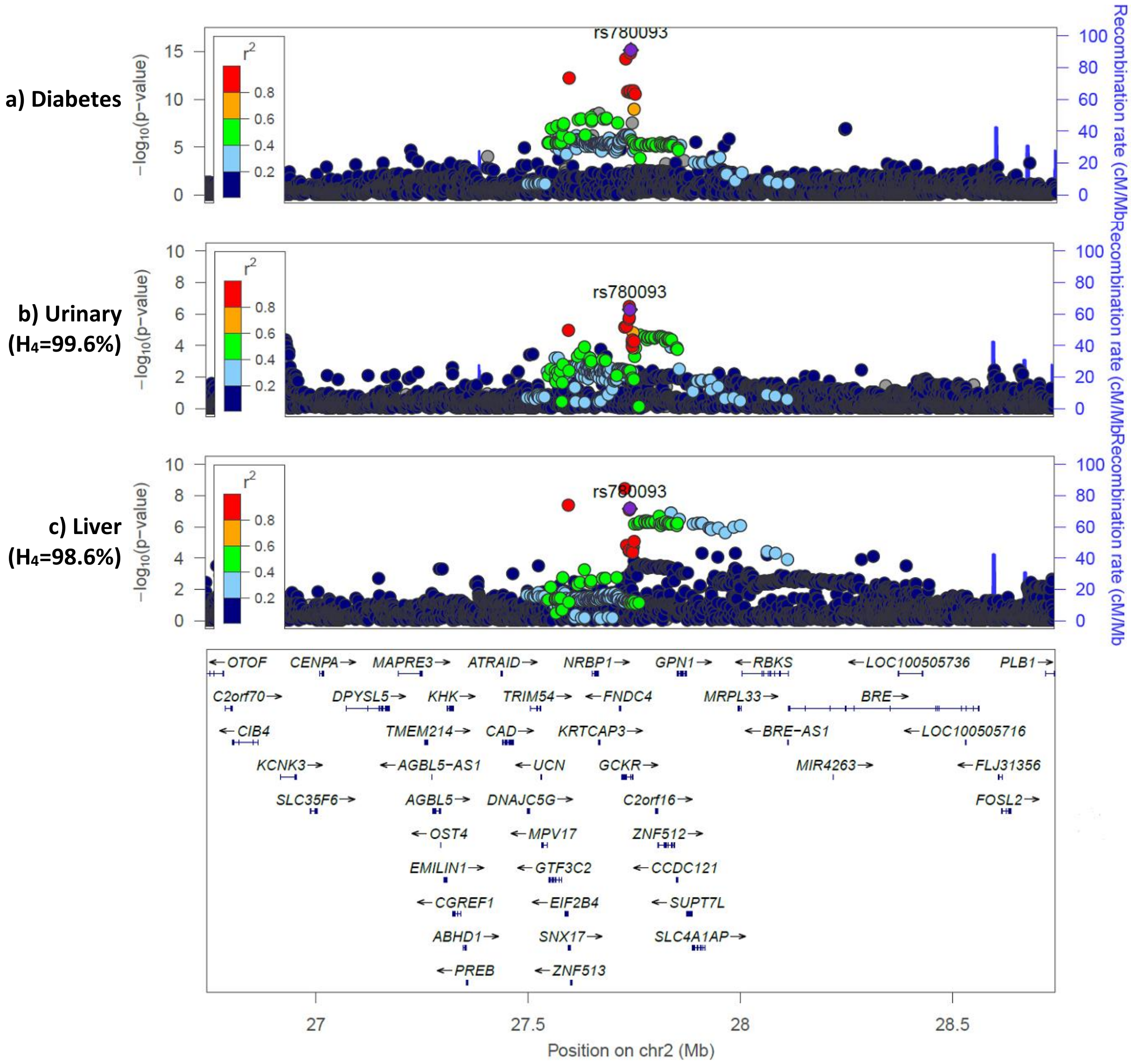

**e) rs13246321 (*TPI1P2*)**

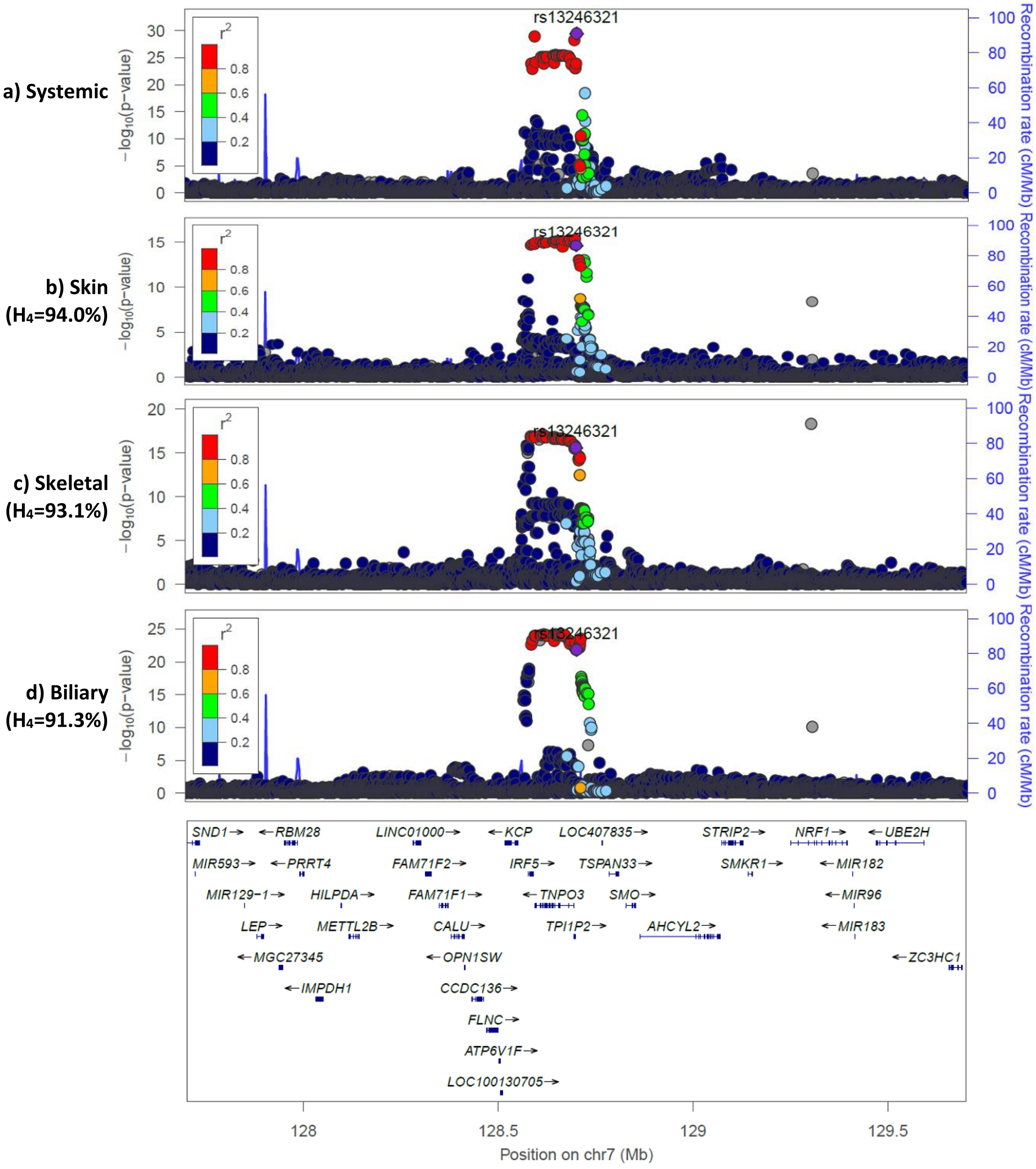

**f) rs2736340 (*BLK*)**

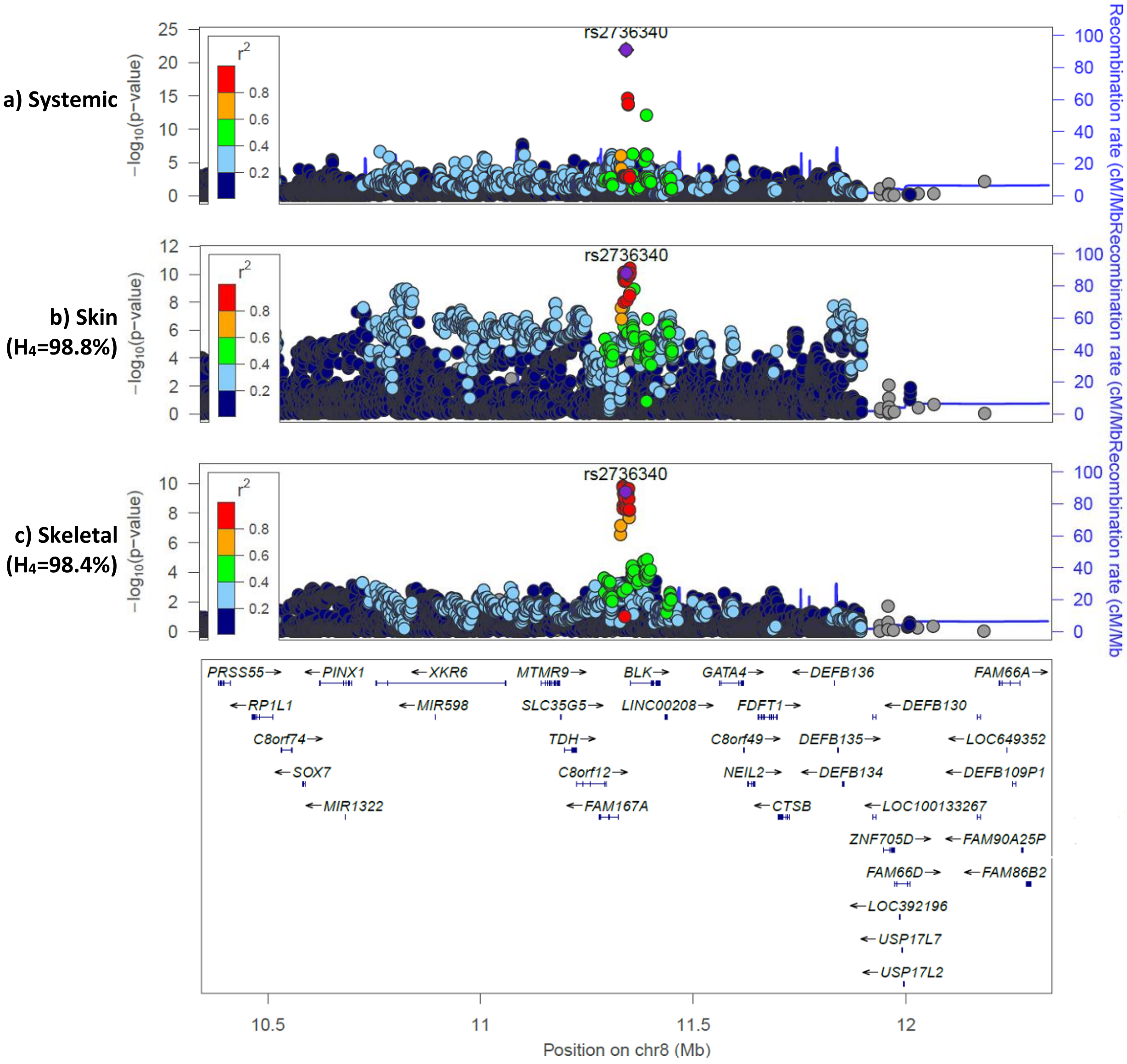

**g) rs8032939 (*RASGRP1*)**

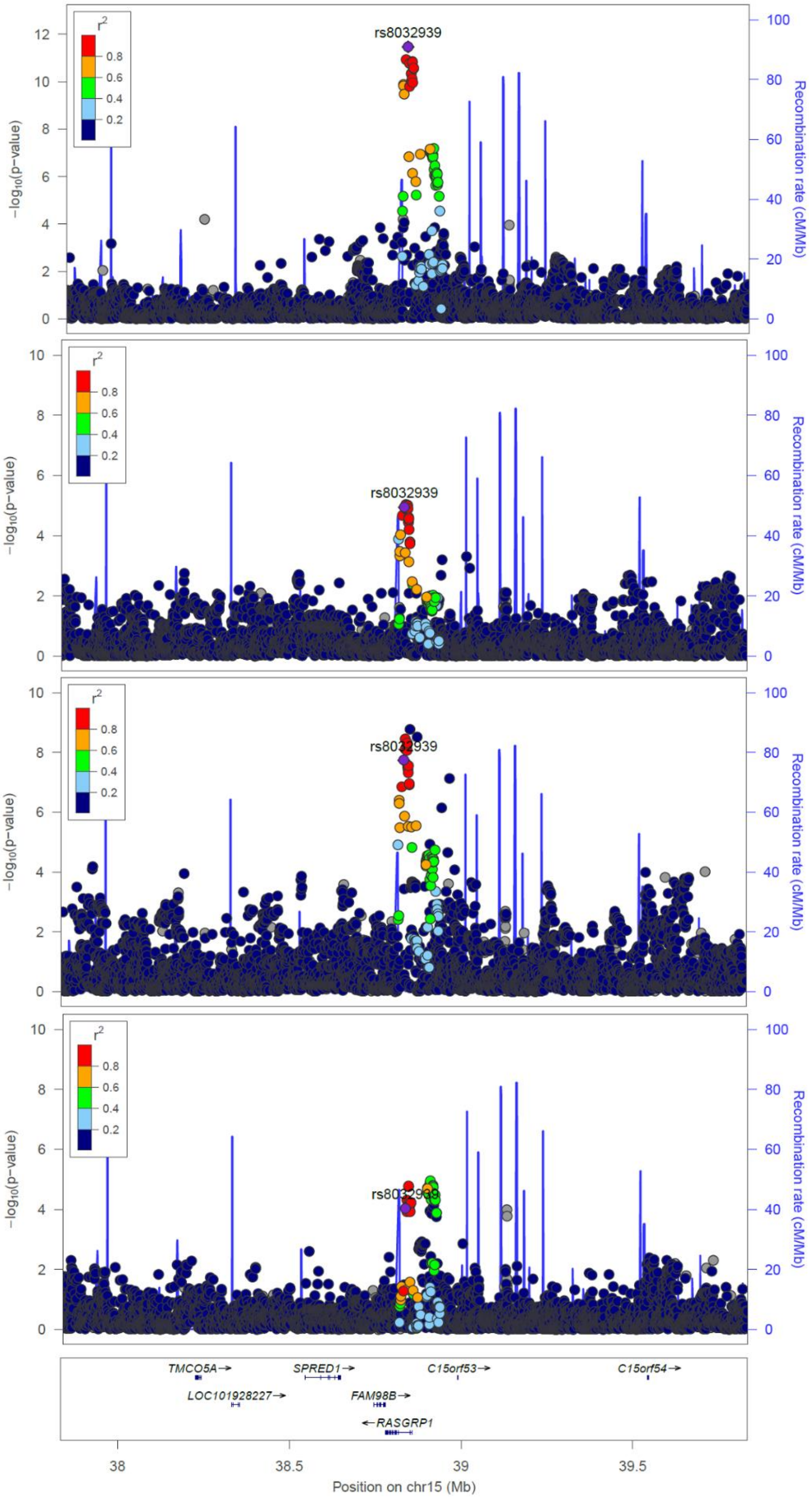

**a) Skeletal**

**b) Cardiovascular ($H_4$=96.1%)**

**c) Diabetes ($H_4$=95.3%)**

**d) Biliary ($H_4$=84.1%)**

**h) rs11089637 (*UBE2L3*)**

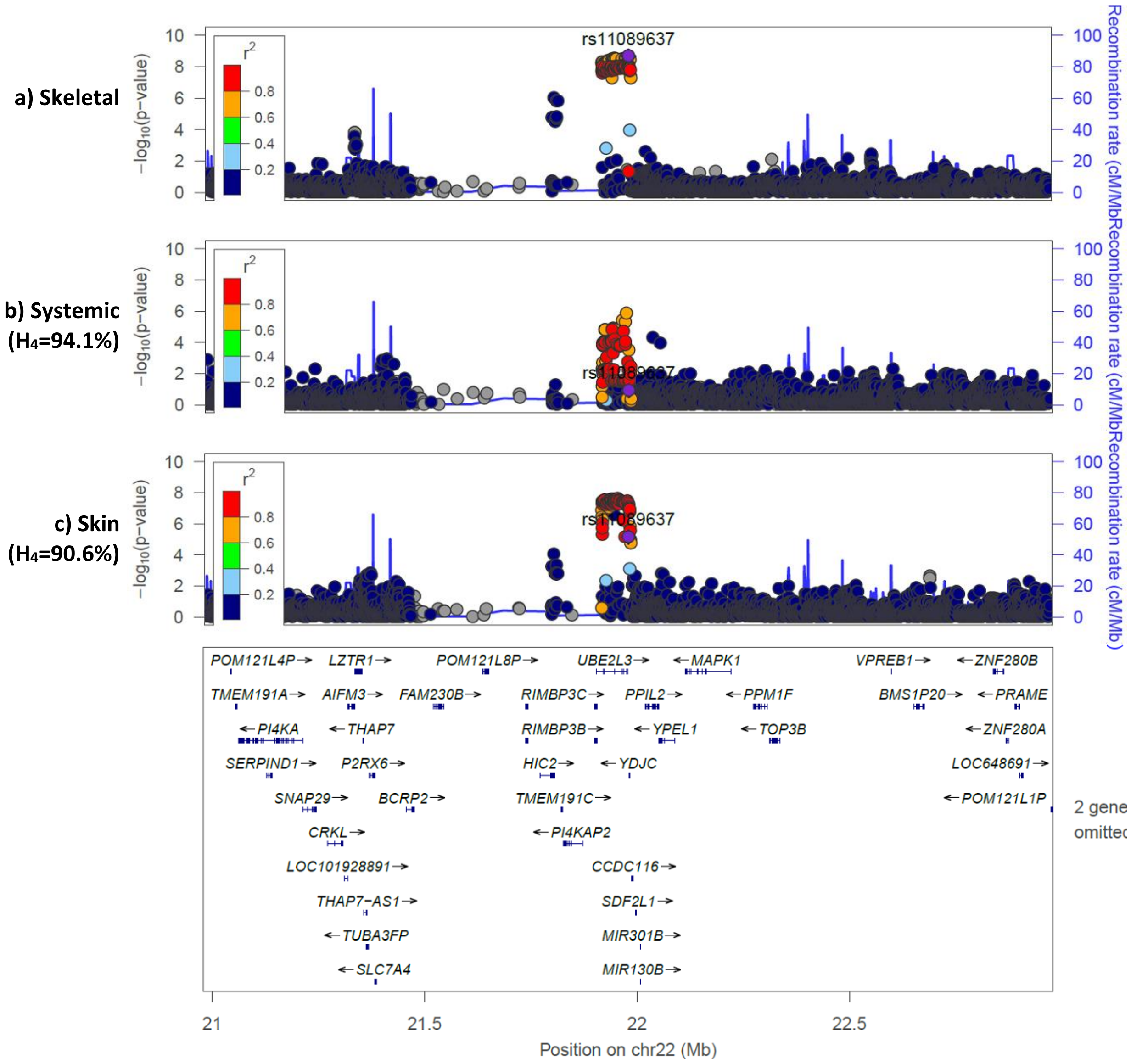

**Supplementary Figure 5: Association of rs7310615 with individual fibrotic diseases**

Forest plots for the association of the rs7310615 variant (sentinel variant for signal that colocalised across five organs and showed some evidence of colocalisation for two other organs) for individual fibrotic diseases. Odds ratios and confidence intervals are given for effect allele C. For analyses performed in UK Biobank (UKB), analyses were performed by ICD10 code.

##### a) Biliary

| Disease | Case | Control | OR [95% CI] | P |
|---|---|---|---|---|
| K74.4 - Secondary biliary cirrhosis (UKB) | 11 | 49,160 | 1.33 [0.57, 3.09] | 0.515 |
| K74.5 - Biliary cirrhosis, unspecified (UKB) | 90 | 49,160 | 1.44 [1.07, 1.94] | 0.017 |
| K80.3 - Calculus of bile duct with cholangitis (UKB) | 315 | 49,160 | 1.12 [1.96, 1.31] | 0.162 |
| K80.4 - Calculus of bile duct with cholecystitis (UKB) | 386 | 49,160 | 0.81 [0.71, 0.94] | 0.005 |
| K80.5 - Calculus of bile duct without cholangitis or cholecystitis (UKB) | 3,055 | 49,160 | 0.98 [0.93, 1.04] | 0.521 |
| K83.0 - Cholangitis (UKB) | 795 | 49,160 | 1.11 [1.00, 1.22] | 0.046 |
| K83.1 - Obstruction of bile duct (UKB) | 1,113 | 49,160 | 1.02 [0.94, 1.11] | 0.590 |
| Biliary fibrosis (UKB) | 4,916 | 49,160 | 1.00 [0.96, 1.05] | 0.921 |
| **Biliary fibrosis (Meta-analysis)** | **4,916** | **49,160** | **1.00 [0.96, 1.05]** | **0.921** |

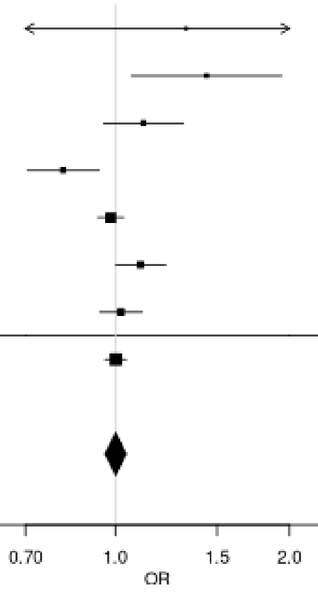

#### b) Cardiovascular

| Disease | Case | Control | OR [95% CI] | P |
|---|---|---|---|---|
| I05.0 - Mitral stenosis (UKB) | 139 | 336,710 | 1.35 [1.06, 1.71] | 0.015 |
| I05.1 - Rheumatic mitral insufficiency (UKB) | 68 | 336,710 | 1.02 [0.73, 1.43] | 0.915 |
| I05.2 - Mitral stenosis with insufficiency (UKB) | 77 | 336,710 | 1.23 [0.89, 1.69] | 0.216 |
| I06.0 - Rheumatic aortic stenosis (UKB) | 27 | 336,710 | 1.09 [0.64, 1.86] | 0.755 |
| I06.2 - Rheumatic aortic stenosis with insufficiency (UKB) | 7 | 336,710 | 2.68 [0.84, 8.54] | 0.096 |
| I08.0 - Disorders of both mitral and aortic valves (UKB) | 761 | 336,710 | 1.09 [0.99, 1.21] | 0.084 |
| I25.1 - Atherosclerotic heart disease (UKB) | 24,023 | 336,710 | 1.08 [1.06, 1.10] | 6.597e-17 |
| I31.8 - Other specified diseases of pericardium (UKB) | 12 | 336,710 | 1.08 [0.48, 2.40] | 0.856 |
| I34.0 - Mitral (valve) insufficiency (UKB) | 3,027 | 336,710 | 0.99 [0.94, 1.04] | 0.619 |
| I34.1 - Mitral (valve) prolapse (UKB) | 726 | 336,710 | 0.98 [0.88, 1.09] | 0.673 |
| I34.2 - Nonrheumatic mitral (valve) stenosis (UKB) | 62 | 336,710 | 1.09 [0.76, 1.55] | 0.644 |
| I35.0 - Aortic (valve) stenosis (UKB) | 2,618 | 336,710 | 1.10 [1.04, 1.16] | 0.0005 |
| I35.2 - Aortic (valve) stenosis with insufficiency (UKB) | 477 | 336,710 | 1.21 [1.06, 1.38] | 0.004 |
| I37.0 - Pulmonary valve stenosis (UKB) | 17 | 336,710 | 0.54 [0.25, 1.15] | 0.111 |
| I47.0 - Re-entry ventricular arrhythmia (UKB) | 78 | 336,710 | 1.16 [0.85, 1.59] | 0.346 |
| I65.2 - Occlusion and stenosis of carotid artery (UKB) | 1,282 | 336,710 | 1.10 [1.01, 1.19] | 0.021 |
| I67.2 - Cerebral atherosclerosis (UKB) | 377 | 336,710 | 0.99 [0.86, 1.15] | 0.922 |
| I70.0 - Atherosclerosis of aorta (UKB) | 239 | 336,710 | 1.08 [0.90, 1.29] | 0.424 |
| I70.00 - Atherosclerosis of aorta (without gangrene) (UKB) | 214 | 336,710 | 0.92 [0.76, 1.11] | 0.384 |
| I70.2 - Atherosclerosis of arteries of the extremities (UKB) | 587 | 336,710 | 1.11 [0.99, 1.25] | 0.079 |
| I70.20 - Atherosclerosis of arteries of the extremities (without gangrene) (UKB) | 526 | 336,710 | 1.08 [0.96, 1.22] | 0.200 |
| I70.8 - Atherosclerosis of other arteries (UKB) | 75 | 336,710 | 1.04 [0.75, 1.44] | 0.813 |
| I70.9 - Generalized and unspecified atherosclerosis (UKB) | 191 | 336,710 | 1.20 [0.98, 1.47] | 0.081 |
| I80.1 - Phlebitis and thrombophlebitis of femoral vein (UKB) | 468 | 336,710 | 1.18 [1.04, 1.35] | 0.011 |
| I80.2 - Phlebitis and thrombophlebitis of other deep vessels of lower extremities (UKB) | 3,937 | 336,710 | 1.07 [1.02, 1.12] | 0.004 |
| M35.2 - Behcet's disease (UKB) | 36 | 336,710 | 0.81 [0.50, 1.30] | 0.377 |
| Q22.1 - Congenital pulmonary valve stenosis (UKB) | 15 | 336,710 | 1.43 [0.70, 2.95] | 0.330 |
| Q23.0 - Congenital stenosis of aortic valve (UKB) | 42 | 336,710 | 1.07 [0.70, 1.64] | 0.767 |
| Q23.3 - Congenital mitral insufficiency (UKB) | 13 | 336,710 | 0.69 [0.31, 1.51] | 0.350 |
| Q24.4 - Congenital subaortic stenosis (UKB) | 7 | 336,710 | 0.82 [0.29, 2.38] | 0.717 |
| Q25.1 - Coarctation of aorta (UKB) | 33 | 336,710 | 1.00 [0.60, 1.68] | 0.987 |
| Q25.3 - Stenosis of aorta (UKB) | 13 | 336,710 | 1.27 [0.59, 2.75] | 0.539 |
| **Cardiovascular fibrosis (UKB)** | **33,671** | **336,710** | **1.07 [1.06, 1.09]** | **9.198e-18** |

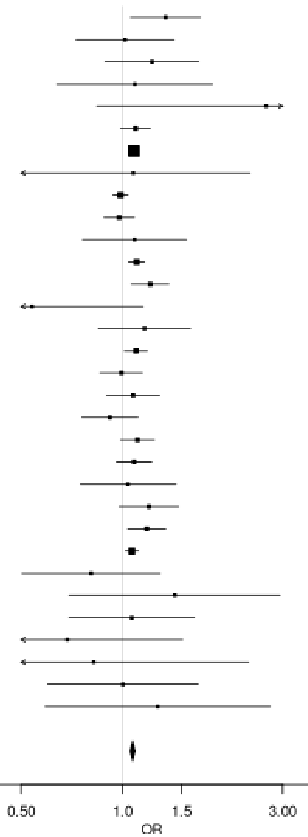

#### c) Diabetes

| Disease | Case | Control | OR [95% CI] | P |
|---|---|---|---|---|
| E10.0 - Type 1 diabetes mellitus with coma (UKB) | 39 | 249,070 | 1.71 [1.09, 2.70] | 0.021 |
| E10.1 - Type 1 diabetes mellitus with ketoacidosis (UKB) | 314 | 249,070 | 1.24 [1.06, 1.47] | 0.008 |
| E10.2 - Type 1 diabetes mellitus with renal complications (UKB) | 101 | 249,070 | 1.20 [0.90, 1.58] | 0.214 |
| E10.3 - Type 1 diabetes mellitus with ophthalmic complications (UKB) | 598 | 249,070 | 1.18 [1.05, 1.32] | 0.005 |
| E10.4 - Type 1 diabetes mellitus with neurological complications (UKB) | 211 | 249,070 | 1.33 [1.10, 1.62] | 0.004 |
| E10.5 - Type 1 diabetes mellitus with circulatory complications (UKB) | 120 | 249,070 | 1.42 [1.09, 1.84] | 0.009 |
| E10.6 - Type 1 diabetes mellitus with other specified complications (UKB) | 45 | 249,070 | 0.95 [0.63, 1.43] | 0.800 |
| E10.7 - Type 1 diabetes mellitus with multiple complications (UKB) | 13 | 249,070 | 0.40 [0.17, 0.95] | 0.038 |
| E10.8 - Type 1 diabetes mellitus with unspecified complications (UKB) | 42 | 249,070 | 1.08 [0.70, 1.66] | 0.735 |
| E10.9 - Type 1 diabetes mellitus without (mention of) complication (UKB) | 3,164 | 249,070 | 1.12 [1.06, 1.18] | 1.380e-05 |
| E11.0 - Type 2 diabetes mellitus with hyperosmolarity (UKB) | 85 | 249,070 | 0.86 [0.63, 1.17] | 0.331 |
| E11.1 - Type 2 diabetes mellitus with ketoacidosis without coma (UKB) | 160 | 249,070 | 1.03 [0.83, 1.29] | 0.792 |
| E11.2 - Type 2 diabetes mellitus with renal complications (UKB) | 298 | 249,070 | 1.08 [0.92, 1.27] | 0.354 |
| E11.3 - Type 2 diabetes mellitus with ophthalmic complications (UKB) | 1,397 | 249,070 | 0.99 [0.92, 1.06] | 0.734 |
| E11.4 - Type 2 diabetes mellitus with neurological complications (UKB) | 661 | 249,070 | 1.13 [1.01, 1.26] | 0.028 |
| E11.5 - Type 2 diabetes mellitus with circulatory complications (UKB) | 446 | 249,070 | 1.09 [0.95, 1.24] | 0.228 |
| E11.6 - Type 2 diabetes mellitus with other specified complications (UKB) | 121 | 249,070 | 1.02 [0.79, 1.31] | 0.907 |
| E11.7 - Type 2 diabetes mellitus with multiple complications (UKB) | 30 | 249,070 | 1.23 [0.74, 2.06] | 0.429 |
| E11.8 - Type 2 diabetes mellitus with unspecified complications (UKB) | 61 | 249,070 | 1.33 [0.93, 1.90] | 0.123 |
| E11.9 - Type 2 diabetes mellitus without (mention of) complication (UKB) | 23,654 | 249,070 | 1.03 [1.01, 1.05] | 0.002 |
| Diabetes (UKB) | 24,907 | 249,070 | 1.04 [1.02, 1.06] | 0.0001 |
| Type 2 diabetes (Mansour Aly et al. 2021) | 9,486 | 2,744 | 1.08 [1.00, 1.15] | 0.029 |
| Type 2 diabetes (Bonàs-Guarch et al. 2018) | 12,931 | 57,196 | 1.03 [0.99, 1.06] | 0.123 |
| **Diabetes (Meta-analysis)** | **47,324** | **309,010** | **1.04 [1.02, 1.05]** | **8.543e-06** |

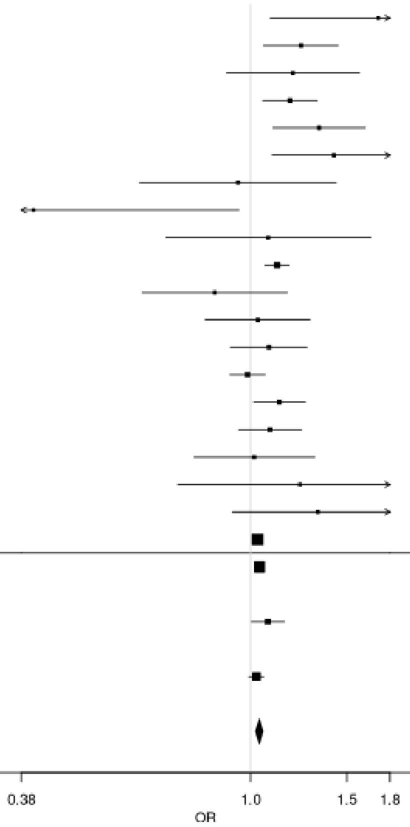

**d) Pulmonary**

| Disease | Case | Control | OR [95% CI] | P |
|---|---|---|---|---|
| J60 - Coalworker pneumoconiosis (UKB) | 31 | 25,980 | 2.30 [1.33, 3.96] | 0.003 |
| J61 - Pneumoconiosis due to asbestos and other mineral fibres (UKB) | 299 | 25,980 | 1.17 [1.00, 1.38] | 0.055 |
| J62.8 - Pneumoconiosis due to other dust containing silica (UKB) | 16 | 25,980 | 1.76 [0.86, 3.61] | 0.123 |
| J63.4 - Siderosis (UKB) | 8 | 25,980 | 1.46 [0.53, 4.00] | 0.467 |
| J67.9 - Hypersensitivity pneumonitis due to unspecified organic dust (UKB) | 86 | 25,980 | 1.04 [0.77, 1.41] | 0.796 |
| J70.3 - Chronic drug-induced interstitial lung disorders (UKB) | 3 | 25,980 | 0.34 [0.03, 3.35] | 0.354 |
| J70.4 - Drug-induced interstitial lung disorders, unspecified (UKB) | 21 | 25,980 | 0.55 [0.28, 1.08] | 0.082 |
| J84.1 - Other interstitial pulmonary diseases with fibrosis (UKB) | 1,485 | 25,980 | 1.04 [0.96, 1.12] | 0.321 |
| J84.8 - Other specified interstitial pulmonary diseases (UKB) | 68 | 25,980 | 1.14 [0.81, 1.59] | 0.464 |
| J84.9 - Interstitial pulmonary disease, unspecified (UKB) | 521 | 25,980 | 1.22 [1.07, 1.38] | 0.002 |
| J92.0 - Pleural plaque with presence of asbestos (UKB) | 602 | 25,980 | 1.10 [0.98, 1.23] | 0.108 |
| J99.0 - Rheumatoid lung disease (UKB) | 47 | 25,980 | 0.93 [0.61, 1.41] | 0.730 |
| Pulmonary fibrosis (UKB) | 2,598 | 25,980 | 1.08 [1.02, 1.14] | 0.009 |
| Idiopathic pulmonary fibrosis (Allen et al. 2022) | 4,125 | 20,464 | 1.07 [1.01, 1.13] | 0.015 |
| **Pulmonary fibrosis (Meta-analysis)** | **6,723** | **46,444** | **1.07 [1.03, 1.12]** | **0.0004** |

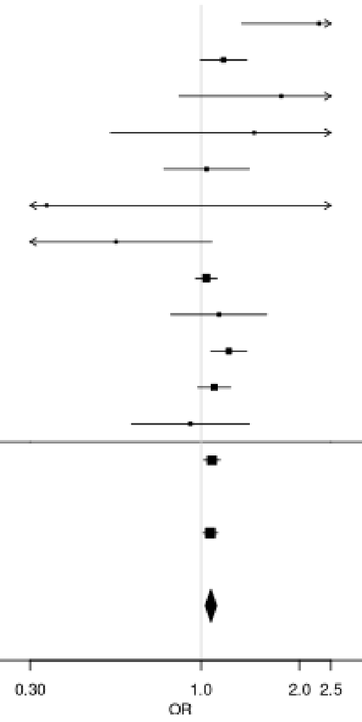

#### e) Skeletal

| Disease | Case | Control | OR [95% CI] | P |
|---|---|---|---|---|
| M05.3 - Rheumatoid arthritis with involvement of other organs and systems (UKB) | 4 | 65,160 | 1.78 [0.42, 7.55] | 0.434 |
| M05.9 - Seropositive rheumatoid arthritis, unspecified (UKB) | 296 | 65,160 | 1.25 [1.06, 1.47] | 0.007 |
| M06.0 - Seronegative rheumatoid arthritis (UKB) | 277 | 65,160 | 1.04 [0.88, 1.23] | 0.620 |
| M06.00 - Seronegative rheumatoid arthritis, unspecified site (UKB) | 238 | 65,160 | 1.02 [0.85, 1.23] | 0.803 |
| M06.9 - Rheumatoid arthritis, unspecified (UKB) | 3,348 | 65,160 | 1.07 [1.02, 1.13] | 0.006 |
| M06.90 - Rheumatoid arthritis, unspecified multiple sites (UKB) | 1,356 | 65,160 | 1.15 [1.07, 1.25] | 0.0003 |
| M06.91 - Rheumatoid arthritis, unspecified shoulder region (UKB) | 128 | 65,160 | 0.96 [0.75, 1.24] | 0.760 |
| M06.92 - Rheumatoid arthritis, unspecified upper arm (UKB) | 60 | 65,160 | 1.19 [0.82, 1.72] | 0.352 |
| M06.93 - Rheumatoid arthritis, unspecified of the forearm (UKB) | 87 | 65,160 | 0.95 [0.71, 1.29] | 0.753 |
| M06.94 - Rheumatoid arthritis, unspecified hand (UKB) | 256 | 65,160 | 1.20 [1.00, 1.43] | 0.045 |
| M06.95 - Rheumatoid arthritis, unspecified pelvic region and thigh (UKB) | 98 | 65,160 | 0.95 [0.72, 1.26] | 0.718 |
| M06.96 - Rheumatoid arthritis, unspecified lower leg (UKB) | 255 | 65,160 | 1.07 [0.90, 1.27] | 0.465 |
| M06.97 - Rheumatoid arthritis, unspecified ankle and foot (UKB) | 211 | 65,160 | 1.09 [0.90, 1.32] | 0.377 |
| M75.0 - Adhesive capsulitis of shoulder (UKB) | 2,023 | 65,160 | 1.02 [0.95, 1.08] | 0.620 |
| Skeletal fibrosis (UKB) | 6,516 | 65,160 | 1.06 [1.02, 1.10] | 0.001 |
| Rheumatoid arthritis (Ishigaki et al. 2022) | 22,350 | 74,823 | 1.09 [1.06, 1.12] | 1.147e-10 |
| **Skeletal fibrosis (Meta-analysis)** | **28,866** | **139,983** | **1.08 [1.06, 1.11]** | **4.191e-12** |

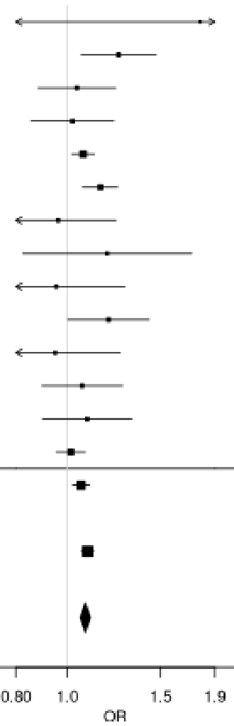

#### f) Systemic

| Disease | Case | Control | OR [95% CI] | P |
|---|---|---|---|---|
| E85.8 - Other amyloidosis (UKB) | 20 | 16,200 | 1.48 [0.79, 2.76] | 0.219 |
| H19.3 - Keratitis and keratoconjunctivitis in other diseases classified elsewhere (sicca) (UKB) | 31 | 16,200 | 1.02 [0.61, 1.70] | 0.936 |
| J98.5 - Diseases of mediastinum, not elsewhere classified (UKB) | 84 | 16,200 | 0.71 [0.52, 0.98] | 0.035 |
| M34.0 - Progressive systemic sclerosis (UKB) | 13 | 16,200 | 0.79 [0.36, 1.71] | 0.546 |
| M34.1 - CR(E)ST syndrome (UKB) | 77 | 16,200 | 0.87 [0.63, 1.21] | 0.408 |
| M34.2 - Systemic sclerosis induced by drugs and chemicals (UKB) | 3 | 16,200 | 0.23 [0.03, 1.97] | 0.179 |
| M34.8 - Other forms of systemic sclerosis (UKB) | 63 | 16,200 | 1.60 [1.11, 2.30] | 0.011 |
| M34.9 - Systemic sclerosis, unspecified (UKB) | 194 | 16,200 | 1.03 [0.84, 1.27] | 0.751 |
| M35.0 - Sicca syndrome (Sjögren's syndrome) (UKB) | 688 | 16,200 | 1.20 [1.08, 1.34] | 0.001 |
| T85.8 - Other complications of internal prosthetic devices, implants and grafts, not elsewhere classified (UKB) | 615 | 16,200 | 1.07 [0.96, 1.20] | 0.234 |
| Systemic fibrosis (UKB) | 1,620 | 16,200 | 1.10 [1.02, 1.18] | 0.010 |
| Systemic sclerosis (López-Isac et al. 2019) | 9,095 | 17,584 | 1.05 [0.98, 1.14] | 0.187 |
| **Systemic fibrosis (Meta-analysis)** | **10,715** | **33,784** | **1.08 [1.02, 1.14]** | **0.006** |

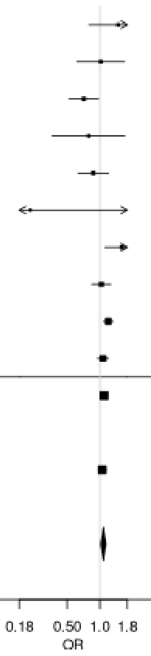

#### g) Urinary

| Disease | Case | Control | OR [95% CI] | P |
|---|---|---|---|---|
| N00.1 - Acute nephritic syndrome, focal and segmental glomerular lesions (UKB) | 4 | 60,110 | 0.68 [1.16, 2.89] | 0.605 |
| N00.2 - Acute nephritic syndrome, diffuse membranous glomerulonephritis (UKB) | 6 | 60,110 | 0.79 [0.25, 2.46] | 0.678 |
| N00.7 - Acute nephritic syndrome, diffuse crescentic glomerulonephritis (UKB) | 3 | 60,110 | 1.07 [0.22, 5.27] | 0.935 |
| N00.8 - Acute nephritic syndrome, other (UKB) | 5 | 60,110 | 1.05 [1.30, 3.63] | 0.939 |
| N00.9 - Acute nephritic syndrome, unspecified (UKB) | 23 | 60,110 | 1.18 [0.65, 2.12] | 0.588 |
| N01.9 - Rapidly progressive nephritic syndrome with unspecified morphological changes (UKB) | 8 | 60,110 | 0.64 [0.23, 1.78] | 0.387 |
| N03.1 - chronic nephritic syndrome, focal and segmental glomerular lesions (UKB) | 18 | 60,110 | 1.50 [0.78, 2.91] | 0.229 |
| N03.2 - chronid nephritic syndrome, diffuse membranous glomerulonephritis (UKB) | 9 | 60,110 | 1.79 [0.65, 4.91] | 0.258 |
| N03.3 - Chronic nephritic syndrome, diffuse mesangial proliferative glomerulonephritis (UKB) | 5 | 60,110 | 1.59 [0.45, 5.60] | 0.469 |
| N03.5 - Chronic nephritic syndrome, diffuse, diffuse mesangiocapillary glomerulonephritis (UKB) | 12 | 60,110 | 1.78 [0.78, 4.04] | 0.170 |
| N03.8 - Chronic nephritic syndrome, other (UKB) | 10 | 60,110 | 1.27 [0.53, 3.06] | 0.595 |
| N03.9 - Chronic nephritic syndrome, unspecified (UKB) | 950 | 60,110 | 1.07 [0.97, 1.17] | 0.165 |
| N04.0 - Nephritic syndrome, minor glomerular abnormality (UKB) | 17 | 60,110 | 1.54 [0.78, 3.04] | 0.213 |
| N04.1 - Nephrotic sundrome, focal and segmental glomerular lesions (UKB) | 21 | 60,110 | 1.58 [0.84, 2.97] | 0.152 |
| N04.2 - Nephrotic syndrome, diffuse membranous glomerulonephritis (UKB) | 19 | 60,110 | 1.21 [0.64, 2.27] | 0.566 |
| N04.3 - Nephrotic syndrome, diffuse mesangial proliferative glomerulonephritis (UKB) | 4 | 60,110 | 0.63 [0.15, 2.61] | 0.521 |
| N04.5 - Nephrotic syndrome, diffuse mesangiocapillary glomerulonephritis | 6 | 60,110 | 0.34 [0.09, 1.26] | 0.106 |
| N04.8 - Nephrotic syndrome, other (UKB) | 11 | 60,110 | 1.08 [0.45, 2.58] | 0.866 |
| N04.9 - Nephrotic syndrome, unspecified (UKB) | 186 | 60,110 | 1.19 [0.97, 1.46] | 0.102 |
| N05.0 - Unspecified nephritic syndrome, minor glomerular abnormality (UKB) | 7 | 60,110 | 1.95 [0.66, 5.75] | 0.229 |
| N05.1 - Unspecified nephritic syndrome, focal and segmental glomerular lesions (UKB) | 45 | 60,110 | 0.98 [0.65, 1.49] | 0.941 |
| N05.2 - Unspecified nephritic syndrome, diffuse membranous glomerulonephritis (UKB) | 52 | 60,110 | 1.17 [0.79, 1.72] | 0.432 |
| N05.3 - Unspecified nephritic syndrome, diffuse mesangial proliferative glomerulonephritis (UKB) | 18 | 60,110 | 0.96 [0.50, 1.84] | 0.895 |
| N05.4 - Unspecified nephritic syndrome, diffuse endocapillary proliferative glomerulonephritis (UKB) | 5 | 60,110 | 0.46 [0.12, 1.75] | 0.253 |
| N05.5 - Unspecified nephritic syndrome, didduse mesangiocapillary glomerulonephritis (UKB) | 23 | 60,110 | 1.67 [0.92, 3.00] | 0.090 |
| N05.7 - Unspecified nephritic syndrome, diffuse crescentic glomerulonephritis (UKB) | 28 | 60,110 | 1.41 [0.83, 2.39] | 0.200 |
| N05.8 - Unspecified nephritic syndrome, other (UKB) | 44 | 60,110 | 0.81 [0.53, 1.24] | 0.334 |
| N05.9 - Unspecified nephritic syndrome, unspecified (UKB) | 179 | 60,110 | 1.16 [0.94, 1.43] | 0.160 |
| N18.3 - Chronic kidney disease, stage 3 (UKB) | 4140 | 60,110 | 1.01 [0.97, 1.06] | 0.617 |
| N18.4 - Chronic kidney disease, stage 4 (UKB) | 596 | 60,110 | 1.13 [1.01, 1.27] | 0.034 |
| N18.5 - Chronic kidney disease, stage 5 (UKB) | 636 | 60,110 | 1.04 [0.93, 1.16] | 0.471 |
| N35.0 - Post-traumatic urethral stricture (UKB) | 350 | 60,110 | 1.06 [0.42, 2.65] | 0.905 |
| N35.1 - Postinfective urethral stricture, not elsewhere classified (UKB) | 351 | 60,110 | 0.76 [0.24, 2.38] | 0.641 |
| N99.1 - Postprocedural urethral stricture (UKB) | 79 | 60,110 | 0.96 [0.69, 1.35] | 0.831 |
| Q61.2 - Polycystic kidney, autosomal dominant (UKB) | 185 | 60,110 | 1.13 [0.92, 1.38] | 0.247 |
| Urinary fibrosis (UKB) | 6,011 | 60,110 | 1.04 [1.00, 1.08] | 0.029 |
| Chronic kidney disease (Wuttke et al. 2019) | 41,395 | 439,303 | 1.04 [1.02, 1.06] | 8.787e-05 |
| **Urinary fibrosis (Meta-analysis)** | **47,406** | **499,413** | **1.04 [1.02, 1.06]** | **1.384e-05** |

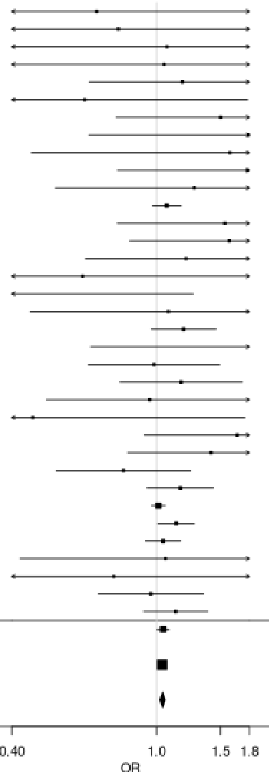

**Supplementary Figure 6: Association of rs6679356 (*IL12RB2*) with individual fibrotic diseases**

**a) Biliary**

| Disease | Case | Control | OR [95% CI] | P |
|---|---|---|---|---|
| K74.4 - Secondary biliary cirrhosis (UKB) | 11 | 49,160 | 0.76 [0.23, 2.55] | 0.653 |
| K74.5 - Biliary cirrhosis, unspecified (UKB) | 90 | 49,160 | 1.54 [0.09, 2.17] | 0.014 |
| K80.3 - Calculus of bile duct with cholangitis (UKB) | 315 | 49,160 | 0.91 [0.73, 1.13] | 0.376 |
| K80.4 - Calculus of bile duct with cholecystitis (UKB) | 386 | 49,160 | 0.97 [0.80, 1.17] | 0.745 |
| K80.5 - Calculus of bile duct without cholangitis or cholecystitis (UKB) | 3,055 | 49,160 | 1.01 [0.94, 1.08] | 0.776 |
| K83.0 - Cholangitis (UKB) | 795 | 49,160 | 1.02 [0.89, 1.16] | 0.777 |
| K83.1 - Obstruction of bile duct (UKB) | 1,113 | 49,160 | 1.02 [0.91, 1.14] | 0.780 |
| Biliary fibrosis (UKB) | 4,916 | 49,160 | 1.03 [0.97, 1.09] | 0.319 |
| Primary biliary cholangitis (Cordell et al. 2021) | 8,021 | 16,489 | 1.55 [1.47, 1.63] | 6.614e-63 |
| **Biliary fibrosis (Meta-analysis)** | **4,916** | **49,160** | **1.28 [1.23, 1.33]** | **2.714e-38** |

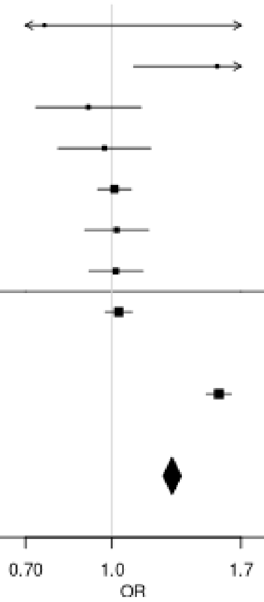

**b) Skeletal**

| Disease | Case | Control | OR [95% CI] | P |
|---|---|---|---|---|
| M05.3 - Rheumatoid arthritis with involvement of other organs and systems (UKB) | 4 | 65,160 | 3.18 [0.74, 13.56] | 0.119 |
| M05.9 - Seropositive rheumatoid arthritis, unspecified (UKB) | 296 | 65,160 | 0.86 [0.69, 1.08] | 0.193 |
| M06.0 - Seronegative rheumatoid arthritis (UKB) | 277 | 65,160 | 0.92 [0.73, 1.15] | 0.454 |
| M06.00 - Seronegative rheumatoid arthritis, unspecified site (UKB) | 238 | 65,160 | 0.91 [0.71, 1.16] | 0.441 |
| M06.9 - Rheumatoid arthritis, unspecified (UKB) | 3,348 | 65,160 | 0.99 [0.93, 1.05] | 0.701 |
| M06.90 - Rheumatoid arthritis, unspecified multiple sites (UKB) | 1,356 | 65,160 | 1.01 [0.91, 1.11] | 0.930 |
| M06.91 - Rheumatoid arthritis, unspecified shoulder region (UKB) | 128 | 65,160 | 1.05 [0.76, 1.44] | 0.783 |
| M06.92 - Rheumatoid arthritis, unspecified upper arm (UKB) | 60 | 65,160 | 1.36 [0.88, 1.10] | 1.161 |
| M06.93 - Rheumatoid arthritis, unspecified of the forearm (UKB) | 87 | 65,160 | 1.10 [0.75, 1.62] | 0.616 |
| M06.94 - Rheumatoid arthritis, unspecified hand (UKB) | 256 | 65,160 | 1.23 [0.99, 1.53] | 0.061 |
| M06.95 - Rheumatoid arthritis, unspecified pelvic region and thigh (UKB) | 98 | 65,160 | 0.07 [0.74, 1.54] | 0.731 |
| M06.96 - Rheumatoid arthritis, unspecified lower leg (UKB) | 255 | 65,160 | 0.98 [0.78, 1.24] | 0.885 |
| M06.97 - Rheumatoid arthritis, unspecified ankle and foot (UKB) | 211 | 65,160 | 1.11 [0.87, 1.42] | 0.394 |
| M75.0 - Adhesive capsulitis of shoulder (UKB) | 2,023 | 65,160 | 1.04 [0.96, 1.13] | 0.322 |
| Skeletal fibrosis (UKB) | 6,516 | 65,160 | 1.01 [0.96, 1.06] | 0.641 |
| Rheumatoid arthritis (Ishigaki et al. 2022) | 22,350 | 74,823 | 1.10 [1.07, 1.14] | 9.951e-09 |
| **Skeletal fibrosis (Meta-analysis)** | **28,866** | **139,983** | **1.07 [1.04, 1.10]** | **1.457e-06** |

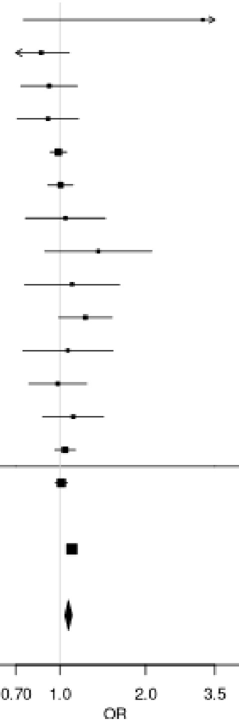

**c) Skin**

| Disease | Case | Control | OR [95% CI] | P |
|---|---|---|---|---|
| L90.5 - Scar conditions and fibrosis of skin (UKB) | 2,789 | 65,930 | 0.96 [0.90, 1.03] | 0.287 |
| L94.0 - Localized scleroderma [morphea] (UKB) | 66 | 65,930 | 1.61 [0.09, 2.38] | 0.018 |
| M32.0 - Drug-induced systemic lupus erythematosus (UKB) | 12 | 65,930 | 2.06[0.85, 4.95] | 0.108 |
| M32.1 - Systemic lupus erythematosus with organ or system involvement (UKB) | 59 | 65,930 | 1.06 [0.66, 1.70] | 0.798 |
| M32.9 - Systemic lupus erythematosus, unspecified (UKB) | 408 | 65,930 | 1.23 [1.03, 1.46] | 0.019 |
| M33.1 - Other dermatomyositis (UKB) | 48 | 65,930 | 1.27 [0.78, 2.07] | 0.345 |
| M35.4 - Diffuse (eosinophilic) fasciitis (UKB) | 7 | 65,930 | 1.31 [0.37, 4.69] | 0.678 |
| M36.0 - Dermato(poly)myositis in neoplastic disease (UKB) | 5 | 65,930 | 1.16 [0.25, 5.49] | 0.849 |
| M72.0 - Palmar fascial fibromatosis [Dupuytren] (UKB) | 3,316 | 65,930 | 1.04 [0.98, 1.11] | 0.210 |
| Skin fibrosis (UKB) | 6,593 | 65,930 | 1.03 [0.98, 1.08] | 0.290 |
| Dupuytren's disease (Riesmeijer et al. 2024) | 7,584 | 28,343 | 1.09 [1.02, 1.16] | 0.009 |
| Systemic lupus erythematosus (Bentham et al. 2015) | 5,201 | 9,066 | 1.17 [1.09, 1.26] | 1.12e-05 |
| Systemic lupus erythematosus (Julia et al. 2018) | 907 | 1,524 | 1.24 [1.03, 1.50] | 0.022 |
| **Skin fibrosis (Meta-analysis)** | **20,285** | **104,863** | **1.08 [1.04, 1.12]** | **5.273e-06** |

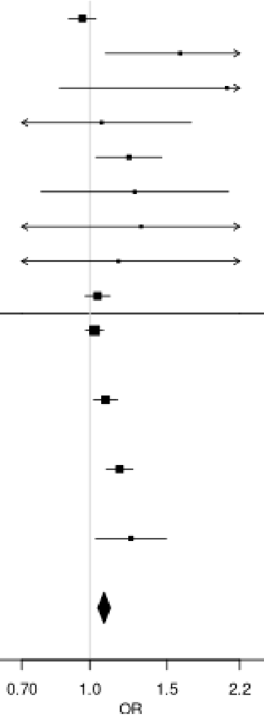

#### Supplementary Figure 7: Association of rs2476601 (*PTPN22*) with individual fibrotic diseases

##### a) Cardiovascular

| Disease | Case | Control | OR [95% CI] | P |
|---|---|---|---|---|
| I05.0 - Mitral stenosis (UKB) | 139 | 336,710 | 1.26 [0.88, 1.80] | 0.213 |
| I05.1 - Rheumatic mitral insufficiency (UKB) | 68 | 336,710 | 1.10 [0.64, 1.88] | 0.724 |
| I05.2 - Mitral stenosis with insufficiency (UKB) | 77 | 336,710 | 1.33 [0.83, 2.13] | 0.235 |
| I06.0 - Rheumatic aortic stenosis (UKB) | 27 | 336,710 | 1.31 [0.59, 1.86] | 0.505 |
| I06.2 - Rheumatic aortic stenosis with insufficiency (UKB) | 7 | 336,710 | 1.15 [0.35, 6.46] | 0.630 |
| I08.0 - Disorders of both mitral and aortic valves (UKB) | 761 | 336,710 | 0.96 [0.81, 1.14] | 0.629 |
| I25.1 - Atherosclerotic heart disease (UKB) | 24,023 | 336,710 | 1.07 [1.04, 1.10] | 6.192e-06 |
| I31.8 - Other specified diseases of pericardium (UKB) | 12 | 336,710 | 1.25 [0.37, 4.17] | 0.721 |
| I34.0 - Mitral (valve) insufficiency (UKB) | 3,027 | 336,710 | 1.11 [1.03, 1.21] | 0.010 |
| I34.1 - Mitral (valve) prolapse (UKB) | 726 | 336,710 | 1.24 [1.06, 1.45] | 0.008 |
| I34.2 - Nonrheumatic mitral (valve) stenosis (UKB) | 62 | 336,710 | 1.23 [0.72, 2.11] | 0.451 |
| I35.0 - Aortic (valve) stenosis (UKB) | 2,618 | 336,710 | 1.08 [0.99, 1.18] | 0.081 |
| I35.2 - Aortic (valve) stenosis with insufficiency (UKB) | 477 | 336,710 | 1.23 [1.02, 1.50] | 0.034 |
| I37.0 - Pulmonary valve stenosis (UKB) | 17 | 336,710 | 0.84 [0.26, 2.75] | 0.77 |
| I47.0 - Re-entry ventricular arrhythmia (UKB) | 78 | 336,710 | 1.17 [0.71, 1.91] | 0.530 |
| I65.2 - Occlusion and stenosis of carotid artery (UKB) | 1,282 | 336,710 | 1.09 [0.96, 1.23] | 0.199 |
| I67.2 - Cerebral atherosclerosis (UKB) | 377 | 336,710 | 1.00 [0.79, 1.27] | 0.985 |
| I70.0 - Atherosclerosis of aorta (UKB) | 239 | 336,710 | 1.22 [0.92, 1.60] | 0.168 |
| I70.00 - Atherosclerosis of aorta (without gangrene) (UKB) | 214 | 336,710 | 1.31 [0.99, 1.74] | 0.060 |
| I70.2 - Atherosclerosis of arteries of the extremities (UKB) | 587 | 336,710 | 1.11 [0.92, 1.33] | 0.275 |
| I70.20 - Atherosclerosis of arteries of the extremities (without gangrene) (UKB) | 526 | 336,710 | 0.90 [0.73, 1.11] | 0.328 |
| I70.8 - Atherosclerosis of other arteries (UKB) | 75 | 336,710 | 1.29 [0.80, 2.09] | 0.296 |
| I70.9 - Generalized and unspecified atherosclerosis (UKB) | 191 | 336,710 | 1.08 [0.78, 1.49] | 0.654 |
| I80.1 - Phlebitis and thrombophlebitis of femoral vein (UKB) | 468 | 336,710 | 1.12 [0.91, 1.37] | 0.289 |
| I80.2 - Phlebitis and thrombophlebitis of other deep vessels of lower extremities (UKB) | 3,937 | 336,710 | 1.08 [1.00, 1.16] | 0.043 |
| M35.2 - Behcet's disease (UKB) | 36 | 336,710 | 1.45 [0.75, 2.83] | 0.273 |
| Q22.1 - Congenital pulmonary valve stenosis (UKB) | 15 | 336,710 | 0.63 [0.91, 2.96] | 0.521 |
| Q23.0 - Congenital stenosis of aortic valve (UKB) | 42 | 336,710 | 1.64 [0.91, 2.96] | 0.101 |
| Q23.3 - Congenital mitral insufficiency (UKB) | 13 | 336,710 | 2.63 [1.06, 6.56] | 0.037 |
| Q24.4 - Congenital subaortic stenosis (UKB) | 7 | 336,710 | 0.66 [0.09, 5.05] | 0.689 |
| Q25.1 - Coarctation of aorta (UKB) | 33 | 336,710 | 1.39 [0.69, 2.81] | 0.356 |
| Q25.3 - Stenosis of aorta (UKB) | 13 | 336,710 | 1.27 [0.59, 3.83] | 0.820 |
| **Cardiovascular fibrosis (UKB)** | **33,671** | **336,710** | **1.07 [1.04, 1.10]** | **6.502e-07** |

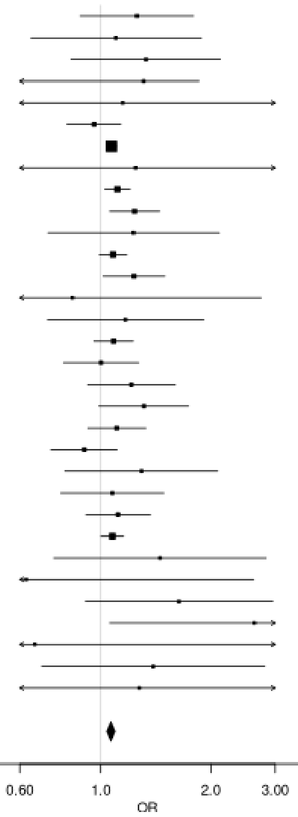

#### b) Diabetes

| Disease | Case | Control | OR [95% CI] | P |
|---|---|---|---|---|
| E10.0 - Type 1 diabetes mellitus with coma (UKB) | 39 | 249,070 | 1.65 [0.89, 3.05] | 0.111 |
| E10.1 - Type 1 diabetes mellitus with ketoacidosis (UKB) | 314 | 249,070 | 1.99 [1.62, 2.44] | 3.346e-11 |
| E10.2 - Type 1 diabetes mellitus with renal complications (UKB) | 101 | 249,070 | 1.93 [1.35, 2.77] | 0.0003 |
| E10.3 - Type 1 diabetes mellitus with ophthalmic complications (UKB) | 598 | 249,070 | 1.54 [1.31, 1.80] | 1.385e-07 |
| E10.4 - Type 1 diabetes mellitus with neurological complications (UKB) | 211 | 249,070 | 1.59 [1.22, 2.07] | 0.0007 |
| E10.5 - Type 1 diabetes mellitus with circulatory complications (UKB) | 120 | 249,070 | 1.88 [1.35, 3.89] | 0.0002 |
| E10.6 - Type 1 diabetes mellitus with other specified complications (UKB) | 45 | 249,070 | 2.06 [1.22, 3.50] | 0.007 |
| E10.7 - Type 1 diabetes mellitus with multiple complications (UKB) | 13 | 249,070 | 1.17 [0.35, 3.89] | 0.798 |
| E10.8 - Type 1 diabetes mellitus with unspecified complications (UKB) | 42 | 249,070 | 2.63 [1.58, 4.39] | 0.0002 |
| E10.9 - Type 1 diabetes mellitus without (mention of) complication (UKB) | 3,164 | 249,070 | 1.40 [1.30, 1.50] | 1.375e-19 |
| E11.0 - Type 2 diabetes mellitus with hyperosmolarity (UKB) | 85 | 249,070 | 1.46 [0.95, 2.25] | 0.084 |
| E11.1 - Type 2 diabetes mellitus with ketoacidosis without coma (UKB) | 160 | 249,070 | 1.58 [1.16, 2.15] | 0.003 |
| E11.2 - Type 2 diabetes mellitus with renal complications (UKB) | 298 | 249,070 | 1.11 [0.86, 1.43] | 0.435 |
| E11.3 - Type 2 diabetes mellitus with ophthalmic complications (UKB) | 1,397 | 249,070 | 1.13 [1.00, 1.27] | 0.045 |
| E11.4 - Type 2 diabetes mellitus with neurological complications (UKB) | 661 | 249,070 | 1.03 [0.86, 1.23] | 0.741 |
| E11.5 - Type 2 diabetes mellitus with circulatory complications (UKB) | 446 | 249,070 | 1.24 [1.01, 1.51] | 0.036 |
| E11.6 - Type 2 diabetes mellitus with other specified complications (UKB) | 121 | 249,070 | 1.06 [0.71, 1.60] | 0.764 |
| E11.7 - Type 2 diabetes mellitus with multiple complications (UKB) | 30 | 249,070 | 1.57 [0.77, 3.19] | 0.213 |
| E11.8 - Type 2 diabetes mellitus with unspecified complications (UKB) | 61 | 249,070 | 1.24 [0.72, 2.13] | 0.432 |
| E11.9 - Type 2 diabetes mellitus without (mention of) complication (UKB) | 23,654 | 249,070 | 1.06 [1.02, 1.09] | 0.0006 |
| Diabetes (UKB) | 24,907 | 249,070 | 1.08 [1.05, 1.11] | 1.574e-06 |
| Type 2 diabetes (Mansour Aly et al. 2021) | 9,486 | 2,744 | 1.04 [0.94, 1.15] | 0.500 |
| Type 2 diabetes (Bonàs-Guarch et al. 2018) | 12,931 | 57,196 | 1.06 [1.01, 1.12] | 0.018 |
| **Diabetes (Meta-analysis)** | **47,324** | **309,010** | **1.07 [1.05, 1.10]** | **9.594e-08** |

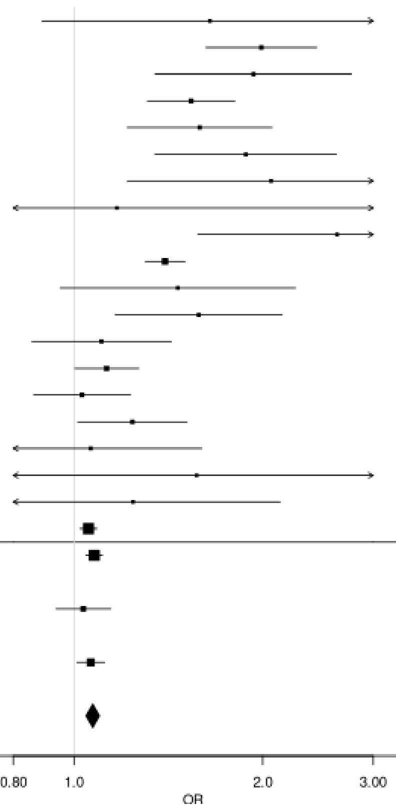

**c) Skeletal**

| Disease | Case | Control | OR [95% CI] | P |
|---|---|---|---|---|
| M05.3 - Rheumatoid arthritis with involvement of other organs and systems (UKB) | 4 | 65,160 | 1.31 [0.16, 10.757] | 0.799 |
| M05.9 - Seropositive rheumatoid arthritis, unspecified (UKB) | 296 | 65,160 | 1.49 [1.18, 1.88] | 0.0007 |
| M06.0 - Seronegative rheumatoid arthritis (UKB) | 277 | 65,160 | 1.31 [1.00, 1.72] | 0.049 |
| M06.00 - Seronegative rheumatoid arthritis, unspecified site (UKB) | 238 | 65,160 | 1.13 [0.87, 1.47] | 0.365 |
| M06.9 - Rheumatoid arthritis, unspecified (UKB) | 3,348 | 65,160 | 1.50 [1.35, 1.67] | 3.138e-13 |
| M06.90 - Rheumatoid arthritis, unspecified multiple sites (UKB) | 1,356 | 65,160 | 1.76 [1.27, 2.46] | 0.0008 |
| M06.91 - Rheumatoid arthritis, unspecified shoulder region (UKB) | 128 | 65,160 | 1.91 [1.19, 3.07] | 0.007 |
| M06.92 - Rheumatoid arthritis, unspecified upper arm (UKB) | 60 | 65,160 | 1.72 [1.14, 2.58] | 0.009 |
| M06.93 - Rheumatoid arthritis, unspecified of the forearm (UKB) | 87 | 65,160 | 1.61 [1.27, 2.06] | 0.0001 |
| M06.94 - Rheumatoid arthritis, unspecified hand (UKB) | 256 | 65,160 | 1.57 [1.06, 2.33] | 0.026 |
| M06.95 - Rheumatoid arthritis, unspecified pelvic region and thigh (UKB) | 98 | 65,160 | 1.49 [1.16, 1.91] | 0.002 |
| M06.96 - Rheumatoid arthritis, unspecified lower leg (UKB) | 255 | 65,160 | 1.65 [1.27, 2.15] | 0.0002 |
| M06.97 - Rheumatoid arthritis, unspecified ankle and foot (UKB) | 211 | 65,160 | 1.38 [1.28, 1.48] | 7.068e-18 |
| M75.0 - Adhesive capsulitis of shoulder (UKB) | 2,023 | 65,160 | 1.19 [1.08, 1.31] | 0.0006 |
| Skeletal fibrosis (UKB) | 6,516 | 65,160 | 1.28 [1.21, 1.35] | 5.812e-18 |
| Rheumatoid arthritis (Ishigaki et al. 2022) | 22,350 | 74,823 | 1.72 [1.66, 1.79] | 3.75e-168 |
| **Skeletal fibrosis (Meta-analysis)** | **28,866** | **139,983** | **1.56 [1.51, 1.61]** | **8.719e-158** |

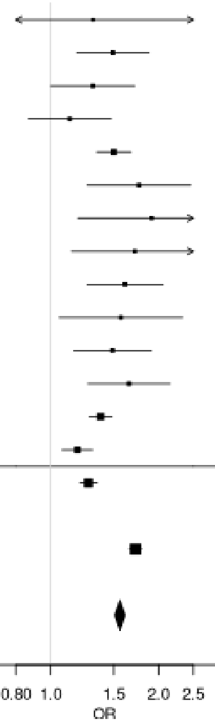

**d) Skin**

| Disease | Case | Control | OR [95% CI] | P |
|---|---|---|---|---|
| L90.5 - Scar conditions and fibrosis of skin (UKB) | 2,789 | 65,930 | 0.92 [0.84, 1.01] | 0.063 |
| L94.0 - Localized scleroderma [morphea] (UKB) | 66 | 65,930 | 1.22 [0.72, 2.06] | 0.457 |
| M32.0 - Drug-induced systemic lupus erythematosus (UKB) | 12 | 65,930 | 1.79 [0.61, 5.25] | 0.290 |
| M32.1 - Systemic lupus erythematosus with organ or system involvement (UKB) | 59 | 65,930 | 1.40 [0.83, 2.38] | 0.209 |
| M32.9 - Systemic lupus erythematosus, unspecified (UKB) | 408 | 65,930 | 1.31 [1.07, 1.61] | 0.010 |
| M33.1 - Other dermatomyositis (UKB) | 48 | 65,930 | 1.16 [0.62, 2.17] | 0.650 |
| M35.4 - Diffuse (eosinophilic) fasciitis (UKB) | 7 | 65,930 | 1.38 [0.31, 6.15] | 0.670 |
| M72.0 - Palmar fascial fibromatosis [Dupuytren] (UKB) | 3,316 | 65,930 | 1.05 [0.97, 1.13] | 0.272 |
| Skin fibrosis (UKB) | 6,593 | 65,930 | 1.01 [0.96, 1.08] | 0.639 |
| Dupuytren's disease (Riesmeijer et al. 2024) | 7,584 | 28,343 | 1.01 [0.94, 1.10] | 0.738 |
| Systemic lupus erythematosus (Bentham et al. 2015) | 5,201 | 9,066 | 1.39 [1.27, 1.52] | 8.382e-13 |
| Systemic lupus erythematosus (Julia et al. 2018) | 907 | 1,524 | 1.39 [1.15, 1.68] | 7e-04 |
| **Skin fibrosis (Meta-analysis)** | **20,285** | **104,863** | **1.10 [1.05, 1.14]** | **8.227e-06** |

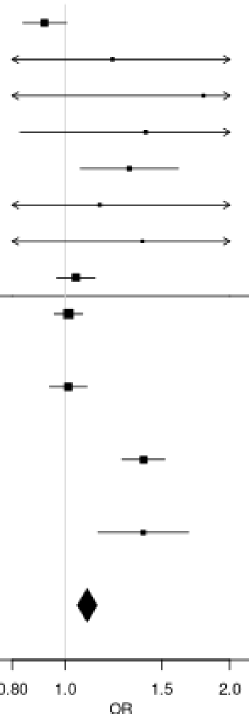

#### Supplementary Figure 8: Association of rs780093 (*GCKR*) with individual fibrotic diseases

##### a) Diabetes

| Disease | Case | Control | OR [95% CI] | P |
|---|---|---|---|---|
| E10.0 - Type 1 diabetes mellitus with coma (UKB) | 39 | 249,070 | 0.84 [0.53, 1.34] | 0.473 |
| E10.1 - Type 1 diabetes mellitus with ketoacidosis (UKB) | 314 | 249,070 | 0.96 [0.81, 1.12] | 0.590 |
| E10.2 - Type 1 diabetes mellitus with renal complications (UKB) | 101 | 249,070 | 0.86 [0.65, 1.15] | 0.318 |
| E10.3 - Type 1 diabetes mellitus with ophthalmic complications (UKB) | 598 | 249,070 | 0.88 [0.79, 1.00] | 0.041 |
| E10.4 - Type 1 diabetes mellitus with neurological complications (UKB) | 211 | 249,070 | 1.06 [0.87, 1.29] | 0.549 |
| E10.5 - Type 1 diabetes mellitus with circulatory complications (UKB) | 120 | 249,070 | 0.93 [0.72, 1.21] | 0.595 |
| E10.6 - Type 1 diabetes mellitus with other specified complications (UKB) | 45 | 249,070 | 0.88 [0.57, 1.36] | 0.569 |
| E10.7 - Type 1 diabetes mellitus with multiple complications (UKB) | 13 | 249,070 | 0.99 [0.45, 2.18] | 0.981 |
| E10.8 - Type 1 diabetes mellitus with unspecified complications (UKB) | 42 | 249,070 | 0.84 [0.54, 1.32] | 0.461 |
| E10.9 - Type 1 diabetes mellitus without (mention of) complication (UKB) | 3,164 | 249,070 | 0.95 [0.90, 0.99] | 0.030 |
| E11.0 - Type 2 diabetes mellitus with hyperosmolarity (UKB) | 85 | 249,070 | 1.00 [0.73, 1.36] | 0.991 |
| E11.1 - Type 2 diabetes mellitus with ketoacidosis without coma (UKB) | 160 | 249,070 | 1.10 [0.88, 1.37] | 0.422 |
| E11.2 - Type 2 diabetes mellitus with renal complications (UKB) | 298 | 249,070 | 1.01 [0.85, 1.19] | 0.948 |
| E11.3 - Type 2 diabetes mellitus with ophthalmic complications (UKB) | 1,397 | 249,070 | 0.91 [0.85, 0.99] | 0.021 |
| E11.4 - Type 2 diabetes mellitus with neurological complications (UKB) | 661 | 249,070 | 0.92 [0.82, 1.03] | 0.154 |
| E11.5 - Type 2 diabetes mellitus with circulatory complications (UKB) | 446 | 249,070 | 0.94 [0.82, 1.08] | 0.404 |
| E11.6 - Type 2 diabetes mellitus with other specified complications (UKB) | 121 | 249,070 | 1.11 [0.86, 1.44] | 0.423 |
| E11.7 - Type 2 diabetes mellitus with multiple complications (UKB) | 30 | 249,070 | 1.00 [0.59, 1.69] | 0.997 |
| E11.8 - Type 2 diabetes mellitus with unspecified complications (UKB) | 61 | 249,070 | 0.67 [0.45, 0.99] | 0.043 |
| E11.9 - Type 2 diabetes mellitus without (mention of) complication (UKB) | 23,654 | 249,070 | 0.93 [0.91, 0.95] | 4.233e-13 |
| Diabetes (UKB) | 24,907 | 249,070 | 0.93 [0.91, 0.95] | 5.857e-13 |
| Type 2 diabetes (Mansour Aly et al. 2021) | 9,486 | 2,744 | 0.97 [0.91, 1.04] | 0.418 |
| Type 2 diabetes (Bonàs-Guarch et al. 2018) | 12,931 | 57,196 | 0.94 [0.91, 0.97] | 0.0002 |
| **Diabetes (Meta-analysis)** | **47,324** | **309,010** | **0.94 [0.92, 0.95]** | **7.148e-16** |

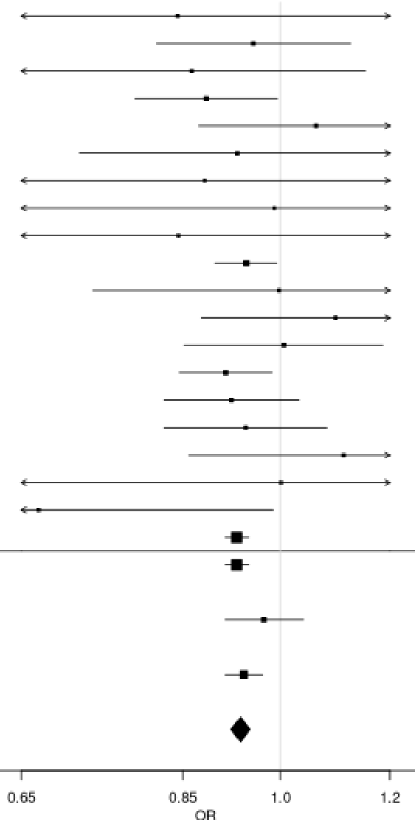

**b) Liver**

| Disease | Case | Control | OR [95% CI] | P |
|---|---|---|---|---|
| B18.2 - Chronic viral hepatitis C (UKB) | 336 | 44,370 | 0.96 [0.82, 1.12] | 0.571 |
| K70.2 - Alcoholic fibrosis and sclerosis of liver (UKB) | 26 | 44,370 | 0.96 [0.54, 1.68] | 0.875 |
| K70.3 - Alcoholic cirrhosis of liver (UKB) | 530 | 44,370 | 0.90 [0.79, 1.02] | 0.096 |
| K70.9 - Alcoholic liver disease, unspecified (UKB) | 769 | 44,370 | 0.93 [0.84, 1.04] | 0.204 |
| K71.2 - Toxic liver disease with acute hepatitis (UKB) | 5 | 44,370 | 0.69 [0.18, 2.72] | 0.598 |
| K71.6 - Toxic liver disease with hepatitis, not elsewhere classified (UKB) | 20 | 44,370 | 0.97 [0.51, 1.84] | 0.924 |
| K71.7 - Toxic liver disease with fibrosis and cirrhosis of liver (UKB) | 6 | 44,370 | 1.51 [0.48, 4.78] | 0.482 |
| K71.9 - Toxic liver disease, unspecified (UKB) | 17 | 44,370 | 1.83 [0.93, 3.59] | 0.079 |
| K74.0 - Hepatic fibrosis (UKB) | 156 | 44,370 | 1.03 [0.82, 1.29] | 0.806 |
| K741 - Hepatic sclerosis (UKB) | 15 | 44,370 | 1.20 [0.59, 2.48] | 0.614 |
| K742 - Hepatic fibrosis with hepatic sclerosis (UKB) | 3 | 44,370 | 3.31 [0.48, 22.83] | 0.225 |
| K74.6 - Other and unspecified cirrhosis of liver (UKB) | 958 | 44,370 | 1.00 [0.91, 1.10] | 0.978 |
| K76.0 - Fatty (change of) liver, not elsewhere classified (UKB) | 2,556 | 44,370 | 1.14 [1.08, 1.21] | 5.799e-06 |
| K76.1 - Chronic passive congestion of liver (UKB) | 29 | 44,370 | 1.00 [0.59, 1.69] | 0.992 |
| Q44.6 - Cystic disease of liver (UKB) | 87 | 44,370 | 1.11 [0.82, 1.51] | 0.490 |
| Liver fibrosis (UKB) | 4,437 | 44,370 | 1.06 [1.01, 1.11] | 0.010 |
| Alcohol-related liver cirrhosis (Buch et al. 2015) | 712 | 1,426 | 0.97 [0.84, 1.11] | 0.649 |
| Non-alcoholic fatty liver disease (Anstee et al. 2020) | 1,483 | 17,781 | 1.26 [1.17, 1.36] | 5.828e-10 |
| Non-alcoholic fatty liver disease (Namjou et al. 2019) | 1,106 | 8,571 | 1.11 [0.99, 1.24] | 0.076 |
| **Liver fibrosis (Meta-analysis)** | **7,738** | **72,148** | **1.10 [1.06, 1.14]** | **7.175e-08** |

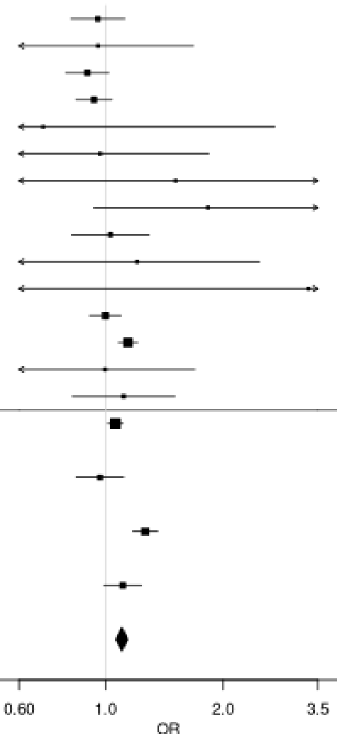

#### c) Urinary

| Disease | Case | Control | OR [95% CI] | P |
|---|---|---|---|---|
| N00.1 - Acute nephritic syndrome, focal and segmental glomerular lesions (UKB) | 4 | 60,110 | 2.66 [0.63, 11.16] | 0.183 |
| N00.2 - Acute nephritic syndrome, diffuse membranous glomerulonephritis (UKB) | 6 | 60,110 | 1.18 [0.37, 3.79] | 0.777 |
| N00.7 - Acute nephritic syndrome, diffuse crescentic glomerulonephritis (UKB) | 3 | 60,110 | 0.33 [0.04, 2.82] | 0.312 |
| N00.8 - Acute nephritic syndrome, other (UKB) | 5 | 60,110 | 1.62 [0.47, 5.65] | 0.447 |
| N00.9 - Acute nephritic syndrome, unspecified (UKB) | 23 | 60,110 | 1.77 [0.99, 3.15] | 0.053 |
| N01.9 - Rapidly progressive nephritic syndrome with unspecified morphological changes (UKB) | 8 | 60,110 | 0.57 [0.18, 1.78] | 0.335 |
| N03.1 - chronic nephritic syndrome, focal and segmental glomerular lesions (UKB) | 18 | 60,110 | 1.29 [0.67, 2.49] | 0.447 |
| N03.2 - chronid nephritic syndrome, diffuse membranous glomerulonephritis (UKB) | 9 | 60,110 | 0.61 [0.22, 1.72] | 0.352 |
| N03.3 - Chronic nephritic syndrome, diffuse mesangial proliferative glomerulonephritis (UKB) | 5 | 60,110 | 0.70 [0.18, 2.71] | 0.609 |
| N03.5 - Chronic nephritic syndrome, diffuse, diffuse mesangiocapillary glomerulonephritis (UKB) | 12 | 60,110 | 0.68 [0.28, 1.63] | 0.383 |
| N03.8 - Chronic nephritic syndrome, other (UKB) | 10 | 60,110 | 0.90 [0.36, 2.25] | 0.814 |
| N03.9 - Chronic nephritic syndrome, unspecified (UKB) | 950 | 60,110 | 0.97 [0.89, 1.07] | 0.577 |
| N04.0 - Nephritic syndrome, minor glomerular abnormality (UKB) | 17 | 60,110 | 1.12 [0.57, 2.21] | 0.749 |
| N04.1 - Nephrotic sundrome, focal and segmental glomerular lesions (UKB) | 21 | 60,110 | 1.11 [0.60, 2.04] | 0.751 |
| N04.2 - Nephrotic syndrome, diffuse membranous glomerulonephritis (UKB) | 19 | 60,110 | 0.94 [0.49, 1.82] | 0.860 |
| N04.3 - Nephrotic syndrome, diffuse mesangial proliferative glomerulonephritis (UKB) | 4 | 60,110 | 0.98 [0.23, 4.11] | 0.979 |
| N04.5 - Nephrotic syndrome, diffuse mesangiocapillary glomerulonephritis | 6 | 60,110 | 0.34 [0.08, 1.52] | 0.157 |
| N04.8 - Nephrotic syndrome, other (UKB) | 11 | 60,110 | 0.94 [0.40, 2.21] | 0.879 |
| N04.9 - Nephrotic syndrome, unspecified (UKB) | 186 | 60,110 | 1.03 [0.84, 1.27] | 0.752 |
| N05.0 - Unspecified nephritic syndrome, minor glomerular abnormality (UKB) | 7 | 60,110 | 0.91 [0.30, 2.73] | 0.861 |
| N05.1 - Unspecified nephritic syndrome, focal and segmental glomerular lesions (UKB) | 45 | 60,110 | 1.24 [0.82, 1.88] | 0.308 |
| N05.2 - Unspecified nephritic syndrome, diffuse membranous glomerulonephritis (UKB) | 52 | 60,110 | 1.46 [0.99, 2.15] | 0.054 |
| N05.3 - Unspecified nephritic syndrome, diffuse mesangial proliferative glomerulonephritis (UKB) | 18 | 60,110 | 0.63 [0.30, 1.30] | 0.210 |
| N05.4 - Unspecified nephritic syndrome, diffuse endocapillary proliferative glomerulonephritis (UKB) | 5 | 60,110 | 2.48 [0.70, 8.78] | 0.158 |
| N05.5 - Unspecified nephritic syndrome, didduse mesangiocapillary glomerulonephritis (UKB) | 23 | 60,110 | 0.95 [0.52, 1.73] | 0.872 |
| N05.7 - Unspecified nephritic syndrome, diffuse crescentic glomerulonephritis (UKB) | 28 | 60,110 | 0.89 [0.52, 1.54] | 0.677 |
| N05.8 - Unspecified nephritic syndrome, other (UKB) | 44 | 60,110 | 1.08 [0.71, 1.66] | 0.712 |
| N05.9 - Unspecified nephritic syndrome, unspecified (UKB) | 179 | 60,110 | 1.13 [0.91, 1.39] | 0.262 |
| N18.3 - Chronic kidney disease, stage 3 (UKB) | 4140 | 60,110 | 0.95 [0.91, 1.00] | 0.039 |
| N18.4 - Chronic kidney disease, stage 4 (UKB) | 596 | 60,110 | 1.02 [0.91, 1.15] | 0.693 |
| N18.5 - Chronic kidney disease, stage 5 (UKB) | 636 | 60,110 | 1.00 [0.89, 1.12] | 0.933 |
| N35.0 - Post-traumatic urethral stricture (UKB) | 350 | 60,110 | 0.62 [0.22, 1.74] | 0.366 |
| N35.1 - Postinfective urethral stricture, not elsewhere classified (UKB) | 351 | 60,110 | 0.57 [0.15, 2.07] | 0.388 |
| N99.1 - Postprocedural urethral stricture (UKB) | 79 | 60,110 | 0.67 [0.47, 0.97] | 0.032 |
| Q61.2 - Polycystic kidney, autosomal dominant (UKB) | 185 | 60,110 | 1.16 [0.95, 1.43] | 0.150 |
| Urinary fibrosis (UKB) | 6,011 | 60,110 | 0.96 [0.93, 1.00] | 0.051 |
| Chronic kidney disease (Wuttke et al. 2019) | 41,395 | 439,303 | 0.96 [0.94, 0.97] | 1.672e-06 |
| **Urinary fibrosis (Meta-analysis)** | **47,406** | **499,413** | **0.96 [0.94, 0.97]** | **5.206e-07** |

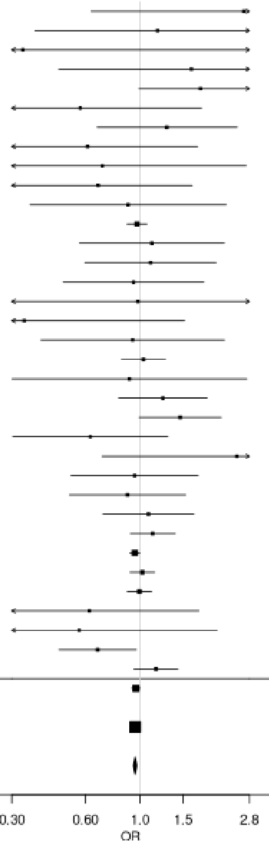

**Supplementary Figure 9: Association of rs13246321 (*TPI1P2*/*TNPO3*) with individual fibrotic diseases**

**a) Biliary**

| Disease | Case | Control | OR [95% CI] | P |
|---|---|---|---|---|
| K74.5 - Biliary cirrhosis, unspecified (UKB) | 90 | 49,160 | 1.75 [1.18, 2.59] | 0.005 |
| K80.3 - Calculus of bile duct with cholangitis (UKB) | 315 | 49,160 | 1.31 [1.04, 1.65] | 0.020 |
| K80.4 - Calculus of bile duct with cholecystitis (UKB) | 386 | 49,160 | 1.16 [0.94, 1.44] | 0.178 |
| K80.5 - Calculus of bile duct without cholangitis or cholecystitis (UKB) | 3,055 | 49,160 | 1.05 [0.97, 1.14] | 0.201 |
| K83.0 - Cholangitis (UKB) | 795 | 49,160 | 0.96 [0.82, 1.13] | 0.650 |
| K83.1 - Obstruction of bile duct (UKB) | 1,113 | 49,160 | 0.98 [0.86, 1.13] | 0.807 |
| Biliary fibrosis (UKB) | 4,916 | 49,160 | 1.03 [0.97, 1.10] | 0.353 |
| Primary biliary cholangitis (Cordell et al. 2021) | 8,021 | 16,489 | 1.51 [1.42, 1.61] | 4.317e-37 |
| **Biliary fibrosis (Meta-analysis)** | **12,937** | **65,649** | **1.26 [1.20, 1.32]** | **1.361e-22** |

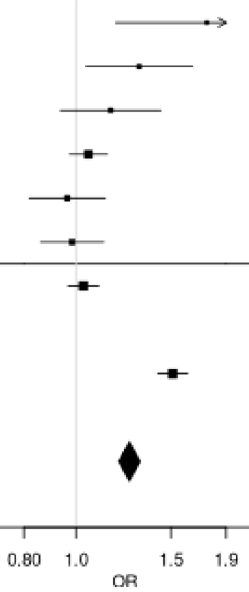

**b) Skeletal**

| Disease | Case | Control | OR [95% CI] | P |
|---|---|---|---|---|
| M05.3 - Rheumatoid arthritis with involvement of other organs and systems (UKB) | 4 | 65,160 | 1.08 [0.14, 8.66] | 0.942 |
| M05.9 - Seropositive rheumatoid arthritis, unspecified (UKB) | 296 | 65,160 | 1.24 [0.98, 1.57] | 0.080 |
| M06.0 - Seronegative rheumatoid arthritis (UKB) | 277 | 65,160 | 1.19 [0.93, 1.53] | 0.176 |
| M06.00 - Seronegative rheumatoid arthritis, unspecified site (UKB) | 238 | 65,160 | 1.18 [0.90, 1.55] | 0.238 |
| M06.9 - Rheumatoid arthritis, unspecified (UKB) | 3,348 | 65,160 | 1.08 [1.00, 1.17] | 0.039 |
| M06.90 - Rheumatoid arthritis, unspecified multiple sites (UKB) | 1,356 | 65,160 | 1.17 [1.04, 1.31] | 0.007 |
| M06.91 - Rheumatoid arthritis, unspecified shoulder region (UKB) | 128 | 65,160 | 0.89 [0.60, 1.35] | 0.577 |
| M06.92 - Rheumatoid arthritis, unspecified upper arm (UKB) | 60 | 65,160 | 1.08 [0.62, 1.90] | 0.776 |
| M06.93 - Rheumatoid arthritis, unspecified of the forearm (UKB) | 87 | 65,160 | 1.27 [0.82, 1.97] | 0.289 |
| M06.94 - Rheumatoid arthritis, unspecified hand (UKB) | 256 | 65,160 | 1.27 [0.99, 1.64] | 0.063 |
| M06.95 - Rheumatoid arthritis, unspecified pelvic region and thigh (UKB) | 98 | 65,160 | 1.71 [1.18, 2.48] | 0.005 |
| M06.96 - Rheumatoid arthritis, unspecified lower leg (UKB) | 255 | 65,160 | 0.99 [0.74, 1.30] | 0.913 |
| M06.97 - Rheumatoid arthritis, unspecified ankle and foot (UKB) | 211 | 65,160 | 1.35 [1.02, 1.78] | 0.036 |
| M75.0 - Adhesive capsulitis of shoulder (UKB) | 2,023 | 65,160 | 1.09 [0.99, 1.20] | 0.620 |
| Skeletal fibrosis (UKB) | 6,516 | 65,160 | 1.10 [1.04, 1.16] | 0.001 |
| Rheumatoid arthritis (Ishigaki et al. 2022) | 22,350 | 74,823 | 1.17 [1.13, 1.22] | 1.181e-15 |
| **Skeletal fibrosis (Meta-analysis)** | **28,866** | **139,983** | **1.15 [1.11, 1.18]** | **2.735e-16** |

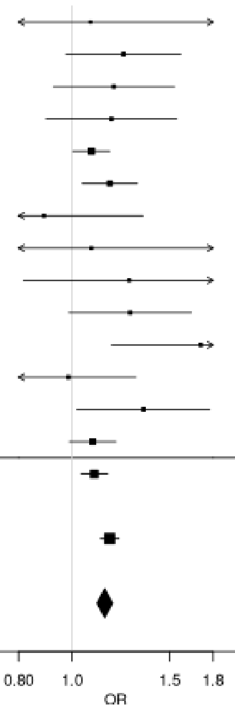

**c) Skin**

| Disease | Case | Control | OR [95% CI] | P |
|---|---|---|---|---|
| L90.5 - Scar conditions and fibrosis of skin (UKB) | 2,789 | 65,930 | 1.02 [0.93, 1.11] | 0.697 |
| L94.0 - Localized scleroderma [morphea] (UKB) | 66 | 65,930 | 1.03 [0.60, 1.76] | 0.920 |
| M32.0 - Drug-induced systemic lupus erythematosus (UKB) | 12 | 65,930 | 1.99 [0.74, 5.35] | 0.170 |
| M32.1 - Systemic lupus erythematosus with organ or system involvement (UKB) | 59 | 65,930 | 1.53 [0.94, 2.50] | 0.089 |
| M32.9 - Systemic lupus erythematosus, unspecified (UKB) | 408 | 65,930 | 1.48 [1.23, 1.79] | 4.852e-05 |
| M33.1 - Other dermatomyositis (UKB) | 48 | 65,930 | 1.28 [0.72, 2.31] | 0.403 |
| M35.4 - Diffuse (eosinophilic) fasciitis (UKB) | 7 | 65,930 | 2.15 [0.59, 7.75] | 0.244 |
| M72.0 - Palmar fascial fibromatosis [Dupuytren] (UKB) | 3,316 | 65,930 | 1.00 [0.93, 1.09] | 0.912 |
| Skin fibrosis (UKB) | 6,593 | 65,930 | 1.04 [0.98, 1.10] | 0.160 |
| Dupuytren's disease (Riesmeijer et al. 2024) | 7,584 | 28,343 | 0.94 [0.87, 1.01] | 0.087 |
| Systemic lupus erythematosus (Bentham et al. 2015) | 5,201 | 9,066 | 1.79 [1.65, 1.94] | 3.658e-44 |
| Systemic lupus erythematosus (Julia et al. 2018) | 907 | 1,524 | 1.03 [0.60, 1.76] | 7.597e-09 |
| **Skin fibrosis (Meta-analysis)** | **20,285** | **104,863** | **1.17 [1.13, 1.22]** | **2.54e-15** |

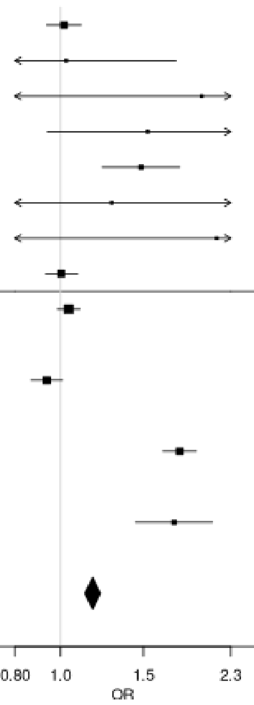

#### d) Systemic

| Disease | Case | Control | OR [95% CI] | P |
|---|---|---|---|---|
| E85.8 - Other amyloidosis (UKB) | 20 | 16,200 | 0.68 [0.21, 2.21] | 0.524 |
| H19.3 - Keratitis and keratoconjunctivitis in other diseases classified elsewhere (sicca) (UKB) | 31 | 16,200 | 2.28 [1.23, 4.22] | 0.009 |
| J98.5 - Diseases of mediastinum, not elsewhere classified (UKB) | 84 | 16,200 | 1.50 [0.98, 2.28] | 0.061 |
| M34.0 - Progressive systemic sclerosis (UKB) | 13 | 16,200 | 0.33 [0.05, 2.49] | 0.284 |
| M34.1 - CR(E)ST syndrome (UKB) | 77 | 16,200 | 1.25 [0.78, 2.00] | 0.360 |
| M34.8 - Other forms of systemic sclerosis (UKB) | 63 | 16,200 | 1.23 [0.72, 2.08] | 0.447 |
| M34.9 - Systemic sclerosis, unspecified (UKB) | 194 | 16,200 | 1.38 [1.04, 1.85] | 0.027 |
| M35.0 - Sicca syndrome (Sjögren's syndrome) (UKB) | 688 | 16,200 | 1.50 [1.29, 1.74] | 1.941e-07 |
| T85.8 - Other complications of internal prosthetic devices, implants and grafts, not elsewhere classified (UKB) | 615 | 16,200 | 1.07 [0.90, 1.28] | 0.449 |
| Systemic fibrosis (UKB) | 1,620 | 16,200 | 1.30 [1.17, 1.45] | 1.298e-06 |
| Systemic sclerosis (López-Isac et al. 2019) | 9,095 | 17,584 | 1.50 [1.38, 1.64] | 1.07e-20 |
| Sjögren's syndrome (Taylor et al. 2017) | 585 | 1,546 | 1.60 [1.33, 1.93] | 5.071e-07 |
| **Systemic fibrosis (Meta-analysis)** | **11,300** | **35,330** | **1.44 [1.35, 1.54]** | **3.825e-30** |

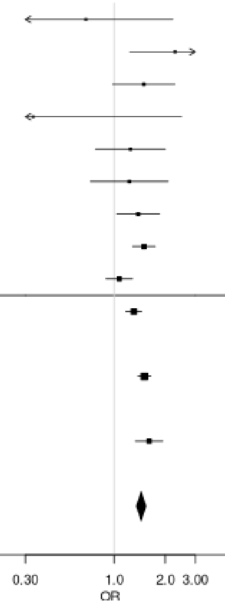

#### Supplementary Figure 10: Association of rs2736340 (*BLK*) with individual fibrotic diseases

##### a) Skeletal

| Disease | Case | Control | OR [95% CI] | P |
|---|---|---|---|---|
| M05.3 - Rheumatoid arthritis with involvement of other organs and systems (UKB) | 4 | 65,160 | 3.01 [0.76, 11.99] | 0.117 |
| M05.9 - Seropositive rheumatoid arthritis, unspecified (UKB) | 296 | 65,160 | 1.20 [1.00, 1.44] | 0.042 |
| M06.0 - Seronegative rheumatoid arthritis (UKB) | 277 | 65,160 | 1.02 [0.84, 1.23] | 0.850 |
| M06.00 - Seronegative rheumatoid arthritis, unspecified site (UKB) | 238 | 65,160 | 0.96 [0.78, 1.18] | 0.669 |
| M06.9 - Rheumatoid arthritis, unspecified (UKB) | 3,348 | 65,160 | 1.08 [1.02, 1.14] | 0.009 |
| M06.90 - Rheumatoid arthritis, unspecified multiple sites (UKB) | 1,356 | 65,160 | 1.09 [1.01, 1.19] | 0.038 |
| M06.91 - Rheumatoid arthritis, unspecified shoulder region (UKB) | 128 | 65,160 | 1.14 [1.87, 1.49] | 0.356 |
| M06.92 - Rheumatoid arthritis, unspecified upper arm (UKB) | 60 | 65,160 | 1.29 [0.87, 1.90] | 0.201 |
| M06.93 - Rheumatoid arthritis, unspecified of the forearm (UKB) | 87 | 65,160 | 1.10 [0.79, 1.54] | 0.571 |
| M06.94 - Rheumatoid arthritis, unspecified hand (UKB) | 256 | 65,160 | 0.96 [0.78, 1.17] | 0.663 |
| M06.95 - Rheumatoid arthritis, unspecified pelvic region and thigh (UKB) | 98 | 65,160 | 0.92 [0.66, 1.27] | 0.597 |
| M06.96 - Rheumatoid arthritis, unspecified lower leg (UKB) | 255 | 65,160 | 0.88 [0.72, 1.08] | 0.229 |
| M06.97 - Rheumatoid arthritis, unspecified ankle and foot (UKB) | 211 | 65,160 | 1.12 [0.89, 1.37] | 0.356 |
| M75.0 - Adhesive capsulitis of shoulder (UKB) | 2,023 | 65,160 | 0.96 [0.89, 1.03] | 0.283 |
| Skeletal fibrosis (UKB) | 6,516 | 65,160 | 1.04 [1.00, 1.09] | 0.052 |
| Rheumatoid arthritis (Ishigaki et al. 2022) | 22,350 | 74,823 | 1.11 [1.08, 1.15] | 3.403e-11 |
| **Skeletal fibrosis (Meta-analysis)** | **28,866** | **139,983** | **1.09 [1.06, 1.11]** | **3.973e-10** |

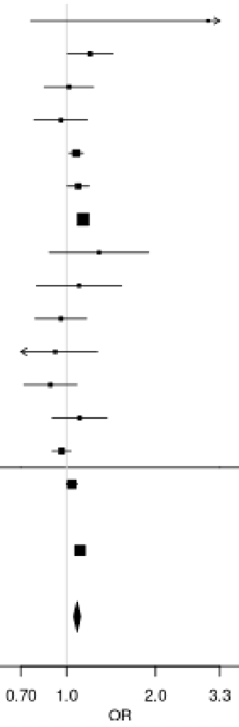

**b) Skin**

| Disease | Case | Control | OR [95% CI] | P |
|---|---|---|---|---|
| L90.5 - Scar conditions and fibrosis of skin (UKB) | 2,789 | 65,930 | 1.05 [0.98, 1.11] | 0.159 |
| L94.0 - Localized scleroderma [morphea] (UKB) | 66 | 65,930 | 1.28 [0.88, 1.85] | 0.197 |
| M32.0 - Drug-induced systemic lupus erythematosus (UKB) | 12 | 65,930 | 2.10[0.94, 4.72] | 0.071 |
| M32.1 - Systemic lupus erythematosus with organ or system involvement (UKB) | 59 | 65,930 | 1.10 [0.73, 1.65] | 0.650 |
| M32.9 - Systemic lupus erythematosus, unspecified (UKB) | 408 | 65,930 | 1.07 [0.92, 1.25] | 0.401 |
| M33.1 - Other dermatomyositis (UKB) | 48 | 65,930 | 1.41 [0.92, 2.16] | 0.118 |
| M35.4 - Diffuse (eosinophilic) fasciitis (UKB) | 7 | 65,930 | 0.75 [0.21, 2.68] | 0.662 |
| M36.0 - Dermato(poly)myositis in neoplastic disease (UKB) | 5 | 65,930 | 0.31 [0.04, 2.48] | 0.272 |
| M72.0 - Palmar fascial fibromatosis [Dupuytren] (UKB) | 3,316 | 65,930 | 1.06 [1.00, 1.12] | 0.058 |
| Skin fibrosis (UKB) | 6,593 | 65,930 | 1.05 [1.01, 1.10] | 0.013 |
| Dupuytren's disease (Riesmeijer et al. 2024) | 7,584 | 28,343 | 1.00 [0.94, 1.07] | 0.933 |
| Systemic lupus erythematosus (Bentham et al. 2015) | 5,201 | 9,066 | 1.30 [1.22, 1.38] | 2.139e-16 |
| Systemic lupus erythematosus (Julia et al. 2018) | 907 | 1,524 | 1.40 [1.20, 1.63] | 2.042e-05 |
| **Skin fibrosis (Meta-analysis)** | **20,285** | **104,863** | **1.10 [1.07, 1.14]** | **8.083e-11** |

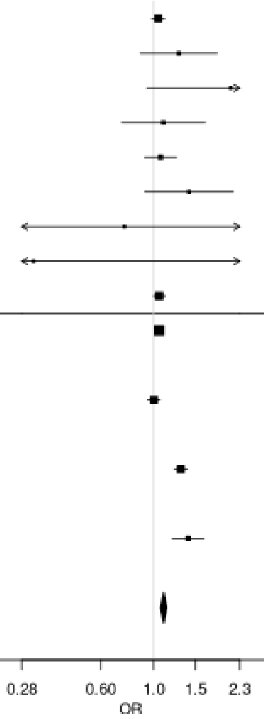

#### c) Systemic

| Disease | Case | Control | OR [95% CI] | P |
|---|---|---|---|---|
| E85.8 - Other amyloidosis (UKB) | 20 | 16,200 | 1.23 [0.63, 2.40] | 0.554 |
| H19.3 - Keratitis and keratoconjunctivitis in other diseases classified elsewhere (sicca) (UKB) | 31 | 16,200 | 1.34 [0.79, 2.30] | 0.282 |
| J98.5 - Diseases of mediastinum, not elsewhere classified (UKB) | 84 | 16,200 | 1.22 [0.88, 1.70] | 0.239 |
| M34.0 - Progressive systemic sclerosis (UKB) | 13 | 16,200 | 0.84 [0.34, 2.08] | 0.706 |
| M34.1 - CR(E)ST syndrome (UKB) | 77 | 16,200 | 1.40 [1.00, 1.96] | 0.051 |
| M34.2 - Systemic sclerosis induced by drugs and chemicals (UKB) | 3 | 16,200 | 1.52 [0.28, 8.37] | 0.633 |
| M34.8 - Other forms of systemic sclerosis (UKB) | 63 | 16,200 | 1.58 [1.09, 2.27] | 0.015 |
| M34.9 - Systemic sclerosis, unspecified (UKB) | 194 | 16,200 | 1.52 [1.23, 1.87] | 0.0001 |
| M35.0 - Sicca syndrome (Sjögren's syndrome) (UKB) | 688 | 16,200 | 1.17 [1.04, 1.32] | 0.011 |
| T85.8 - Other complications of internal prosthetic devices, implants and grafts, not elsewhere classified (UKB) | 615 | 16,200 | 1.01 [0.88, 1.15] | 0.917 |
| Systemic fibrosis (UKB) | 1,620 | 16,200 | 1.14 [1.05, 1.23] | 0.002 |
| Systemic sclerosis (López-Isac et al. 2019) | 9,095 | 17,584 | 1.24 [1.19, 1.30] | 3.331e-21 |
| Sjögren's syndrome (Taylor et al. 2017) | 585 | 1,546 | 1.10 [1.96, 1.27] | 0.172 |
| **Systemic fibrosis (Meta-analysis)** | **11,300** | **35,330** | **1.21 [1.16, 1.26]** | **1.211e-22** |

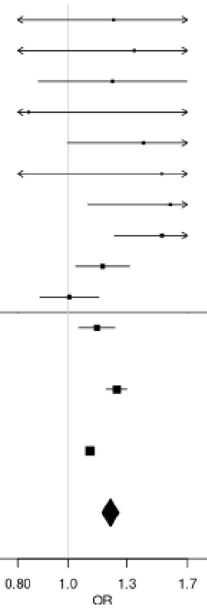

**Supplementary Figure 11: Association of rs8032939 (*RASGRP1*) with individual fibrotic diseases**

**a) Biliary**

| Disease | Case | Control | OR [95% CI] | P |
|---|---|---|---|---|
| K74.4 - Secondary biliary cirrhosis (UKB) | 11 | 49,160 | 1.11 [0.43, 2.87] | 0.823 |
| K74.5 - Biliary cirrhosis, unspecified (UKB) | 90 | 49,160 | 1.16 [0.84, 1.61] | 0.378 |
| K80.3 - Calculus of bile duct with cholangitis (UKB) | 315 | 49,160 | 0.97 [0.81, 1.16] | 0.730 |
| K80.4 - Calculus of bile duct with cholecystitis (UKB) | 386 | 49,160 | 1.05 [0.89, 1.23] | 0.576 |
| K80.5 - Calculus of bile duct without cholangitis or cholecystitis (UKB) | 3,055 | 49,160 | 1.04 [0.98, 1.11] | 0.156 |
| K83.0 - Cholangitis (UKB) | 795 | 49,160 | 1.11 [0.99, 1.24] | 0.073 |
| K83.1 - Obstruction of bile duct (UKB) | 1,113 | 49,160 | 1.00 [0.90, 1.10] | 0.933 |
| Biliary fibrosis (UKB) | 4,916 | 49,160 | 1.05 [1.00, 1.10] | 0.037 |
| Primary biliary cholangitis (Cordell et al. 2021) | 8,021 | 16,489 | 1.09 [1.04, 1.14] | 0.0006 |
| **Biliary fibrosis (Meta-analysis)** | **4,916** | **49,160** | **1.07 [1.03, 1.11]** | **9.299e-05** |

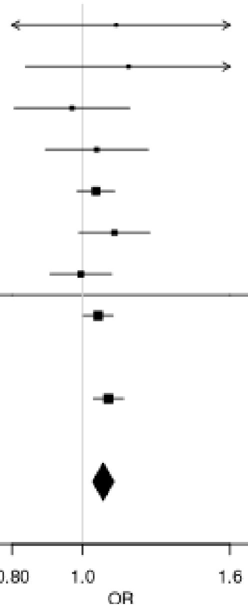

#### b) Cardiovascular

| Disease | Case | Control | OR [95% CI] | P |
|---|---|---|---|---|
| I05.0 - Mitral stenosis (UKB) | 139 | 336,710 | 0.98 [0.75, 1.29] | 0.898 |
| I05.1 - Rheumatic mitral insufficiency (UKB) | 68 | 336,710 | 1.15 [0.79, 1.68] | 0.471 |
| I05.2 - Mitral stenosis with insufficiency (UKB) | 77 | 336,710 | 1.12 [0.78, 1.60] | 0.537 |
| I06.0 - Rheumatic aortic stenosis (UKB) | 27 | 336,710 | 0.79 [0.41, 1.53] | 0.479 |
| I06.2 - Rheumatic aortic stenosis with insufficiency (UKB) | 7 | 336,710 | 0.83 [0.23, 2.99] | 0.781 |
| I08.0 - Disorders of both mitral and aortic valves (UKB) | 761 | 336,710 | 1.04 [0.93, 1.17] | 0.486 |
| I25.1 - Atherosclerotic heart disease (UKB) | 24,023 | 336,710 | 1.04 [1.01, 1.06] | 0.002 |
| I31.8 - Other specified diseases of pericardium (UKB) | 12 | 336,710 | 1.83 [0.80, 4.16] | 0.152 |
| I34.0 - Mitral (valve) insufficiency (UKB) | 3,027 | 336,710 | 1.03 [0.97, 1.09] | 0.340 |
| I34.1 - Mitral (valve) prolapse (UKB) | 726 | 336,710 | 1.11 [0.98, 1.24] | 0.090 |
| I34.2 - Nonrheumatic mitral (valve) stenosis (UKB) | 62 | 336,710 | 1.12 [0.75, 1.67] | 0.578 |
| I35.0 - Aortic (valve) stenosis (UKB) | 2,618 | 336,710 | 1.07 [1.00, 1.14] | 0.039 |
| I35.2 - Aortic (valve) stenosis with insufficiency (UKB) | 477 | 336,710 | 1.11 [0.96, 1.28] | 0.152 |
| I37.0 - Pulmonary valve stenosis (UKB) | 17 | 336,710 | 0.94 [0.43, 2.08] | 0.881 |
| I47.0 - Re-entry ventricular arrhythmia (UKB) | 78 | 336,710 | 0.82 [0.56, 1.21] | 0.319 |
| I65.2 - Occlusion and stenosis of carotid artery (UKB) | 1,282 | 336,710 | 1.06 [0.97, 1.16] | 0.190 |
| I67.2 - Cerebral atherosclerosis (UKB) | 377 | 336,710 | 1.19 [1.01, 1.39] | 0.035 |
| I70.0 - Atherosclerosis of aorta (UKB) | 239 | 336,710 | 1.10 [0.90, 1.35] | 0.339 |
| I70.00 - Atherosclerosis of aorta (without gangrene) (UKB) | 214 | 336,710 | 1.14 [0.92, 1.41] | 0.229 |
| I70.2 - Atherosclerosis of arteries of the extremities (UKB) | 587 | 336,710 | 0.94 [0.82, 1.08] | 0.399 |
| I70.20 - Atherosclerosis of arteries of the extremities (without gangrene) (UKB) | 526 | 336,710 | 0.99 [0.86, 1.14] | 0.881 |
| I70.9 - Generalized and unspecified atherosclerosis (UKB) | 191 | 336,710 | 0.80 [0.63, 1.03] | 0.083 |
| I80.1 - Phlebitis and thrombophlebitis of femoral vein (UKB) | 468 | 336,710 | 0.99 [0.85, 1.15] | 0.861 |
| I80.2 - Phlebitis and thrombophlebitis of other deep vessels of lower extremities (UKB) | 3,937 | 336,710 | 1.05 [1.00, 1.11] | 0.044 |
| M35.2 - Behcet's disease (UKB) | 36 | 336,710 | 0.88 [0.51, 1.54] | 0.658 |
| Q22.1 - Congenital pulmonary valve stenosis (UKB) | 15 | 336,710 | 1.11 [0.49, 2.48] | 0.807 |
| Q23.3 - Congenital mitral insufficiency (UKB) | 13 | 336,710 | 0.73 [0.28, 1.93] | 0.526 |
| Q24.4 - Congenital subaortic stenosis (UKB) | 7 | 336,710 | 1.69 [0.57, 5.05] | 0.346 |
| Q25.1 - Coarctation of aorta (UKB) | 33 | 336,710 | 1.54 [0.92, 2.56] | 0.100 |
| Q25.3 - Stenosis of aorta (UKB) | 13 | 336,710 | 1.64 [0.73, 3.68] | 0.231 |
| **Cardiovascular fibrosis (UKB)** | **33,671** | **336,710** | **1.04 [1.02, 1.06]** | **1.144e-05** |

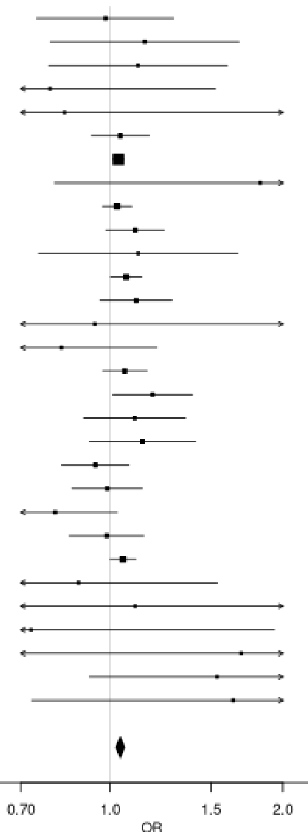

#### c) Diabetes

| Disease | Case | Control | OR [95% CI] | P |
|---|---|---|---|---|
| E10.0 - Type 1 diabetes mellitus with coma (UKB) | 39 | 249,070 | 1.95 [1.23, 3.07] | 0.004 |
| E10.1 - Type 1 diabetes mellitus with ketoacidosis (UKB) | 314 | 249,070 | 1.05 [0.88, 1.26] | 0.599 |
| E10.2 - Type 1 diabetes mellitus with renal complications (UKB) | 101 | 249,070 | 0.89 [0.64, 1.23] | 0.472 |
| E10.3 - Type 1 diabetes mellitus with ophthalmic complications (UKB) | 598 | 249,070 | 0.98 [0.86, 1.12] | 0.795 |
| E10.4 - Type 1 diabetes mellitus with neurological complications (UKB) | 211 | 249,070 | 0.95 [0.76, 1.19] | 0.635 |
| E10.5 - Type 1 diabetes mellitus with circulatory complications (UKB) | 120 | 249,070 | 1.10 [0.83, 1.47] | 0.520 |
| E10.6 - Type 1 diabetes mellitus with other specified complications (UKB) | 45 | 249,070 | 1.18 [0.75, 1.88] | 0.474 |
| E10.7 - Type 1 diabetes mellitus with multiple complications (UKB) | 13 | 249,070 | 1.14 [0.48, 2.70] | 0.775 |
| E10.8 - Type 1 diabetes mellitus with unspecified complications (UKB) | 42 | 249,070 | 1.10 [0.68, 1.79] | 0.700 |
| E10.9 - Type 1 diabetes mellitus without (mention of) complication (UKB) | 3,164 | 249,070 | 1.08 [1.02, 1.14] | 0.009 |
| E11.0 - Type 2 diabetes mellitus with hyperosmolarity (UKB) | 85 | 249,070 | 1.36 [0.98, 1.88] | 0.065 |
| E11.1 - Type 2 diabetes mellitus with ketoacidosis without coma (UKB) | 160 | 249,070 | 1.31 [1.03, 1.66] | 0.029 |
| E11.2 - Type 2 diabetes mellitus with renal complications (UKB) | 298 | 249,070 | 0.85 [0.70, 1.04] | 0.110 |
| E11.3 - Type 2 diabetes mellitus with ophthalmic complications (UKB) | 1,397 | 249,070 | 1.06 [0.97, 1.16] | 0.178 |
| E11.4 - Type 2 diabetes mellitus with neurological complications (UKB) | 661 | 249,070 | 1.05 [0.93, 1.19] | 0.466 |
| E11.5 - Type 2 diabetes mellitus with circulatory complications (UKB) | 446 | 249,070 | 0.98 [0.84, 1.14] | 0.745 |
| E11.6 - Type 2 diabetes mellitus with other specified complications (UKB) | 121 | 249,070 | 0.93 [0.69, 1.25] | 0.611 |
| E11.7 - Type 2 diabetes mellitus with multiple complications (UKB) | 30 | 249,070 | 1.67 [0.98, 2.83] | 0.059 |
| E11.8 - Type 2 diabetes mellitus with unspecified complications (UKB) | 61 | 249,070 | 1.01 [0.67, 1.52] | 0.975 |
| E11.9 - Type 2 diabetes mellitus without (mention of) complication (UKB) | 23,654 | 249,070 | 1.06 [1.03, 1.08] | 7.393e-07 |
| Diabetes (UKB) | 24,907 | 249,070 | 1.06 [1.04, 1.08] | 4.424e-07 |
| Type 2 diabetes (Mansour Aly et al. 2021) | 9,486 | 2,744 | 1.12 [1.04, 1.20] | 0.002 |
| Type 2 diabetes (Bonàs-Guarch et al. 2018) | 12,931 | 57,196 | 1.03 [0.99, 1.06] | 0.172 |
| **Diabetes (Meta-analysis)** | **47,324** | **309,010** | **1.05 [1.03, 1.07]** | **1.838e-08** |

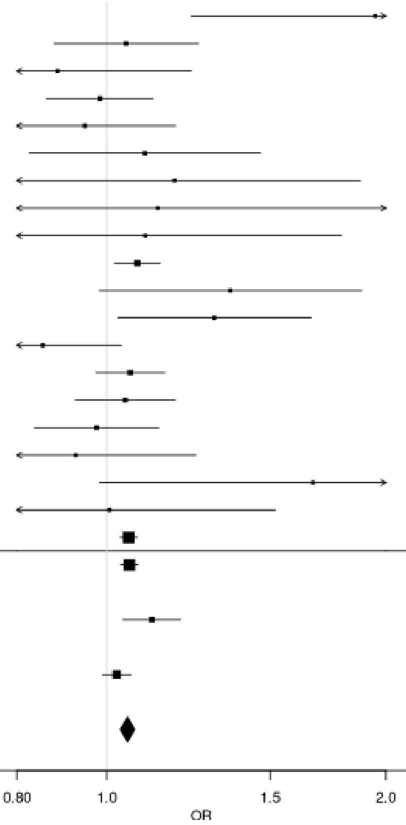

#### d) Skeletal

| Disease | Case | Control | OR [95% CI] | P |
|---|---|---|---|---|
| M05.3 - Rheumatoid arthritis with involvement of other organs and systems (UKB) | 4 | 65,160 | 0.46 [0.06, 3.81] | 0.474 |
| M05.9 - Seropositive rheumatoid arthritis, unspecified (UKB) | 296 | 65,160 | 1.01 [0.84, 1.22] | 0.924 |
| M06.0 - Seronegative rheumatoid arthritis (UKB) | 277 | 65,160 | 1.21 [1.00, 1.46] | 0.564 |
| M06.00 - Seronegative rheumatoid arthritis, unspecified site (UKB) | 238 | 65,160 | 1.06 [0.86, 1.31] | 0.047 |
| M06.9 - Rheumatoid arthritis, unspecified (UKB) | 3,348 | 65,160 | 1.08 [1.02, 1.14] | 0.057 |
| M06.90 - Rheumatoid arthritis, unspecified multiple sites (UKB) | 1,356 | 65,160 | 1.09 [1.00, 1.19] | 0.180 |
| M06.91 - Rheumatoid arthritis, unspecified shoulder region (UKB) | 128 | 65,160 | 1.21 [0.92, 1.59] | 0.232 |
| M06.92 - Rheumatoid arthritis, unspecified upper arm (UKB) | 60 | 65,160 | 1.27 [0.86, 1.89] | 0.907 |
| M06.93 - Rheumatoid arthritis, unspecified of the forearm (UKB) | 87 | 65,160 | 0.98 [0.69, 1.39] | 0.545 |
| M06.94 - Rheumatoid arthritis, unspecified hand (UKB) | 256 | 65,160 | 1.06 [0.87, 1.30] | 0.521 |
| M06.95 - Rheumatoid arthritis, unspecified pelvic region and thigh (UKB) | 98 | 65,160 | 1.11 [0.81, 1.53] | 0.973 |
| M06.96 - Rheumatoid arthritis, unspecified lower leg (UKB) | 255 | 65,160 | 1.00 [1.82, 1.23] | 0.181 |
| M06.97 - Rheumatoid arthritis, unspecified ankle and foot (UKB) | 211 | 65,160 | 1.16 [0.93, 1.44] | 0.012 |
| M75.0 - Adhesive capsulitis of shoulder (UKB) | 2,023 | 65,160 | 1.00 [0.93, 1.08] | 0.992 |
| Skeletal fibrosis (UKB) | 6,516 | 65,160 | 1.05 [1.01, 1.10] | 0.020 |
| Rheumatoid arthritis (Ishigaki et al. 2022) | 22,350 | 74,823 | 1.11 [1.08, 1.14] | 1.029e-12 |
| **Skeletal fibrosis (Meta-analysis)** | **28,866** | **139,983** | **1.09 [1.06, 1.12]** | **3.420e-12** |

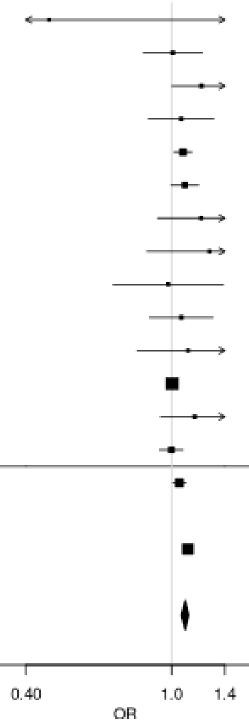

**Supplementary Figure 12: Association of rs11089637 (*UBE2L3*) with individual fibrotic diseases**

**a) Skeletal**

| Disease | Case | Control | OR [95% CI] | P |
|---|---|---|---|---|
| M05.3 - Rheumatoid arthritis with involvement of other organs and systems (UKB) | 4 | 65,160 | 3.39 [0.84, 13.69] | 0.086 |
| M05.9 - Seropositive rheumatoid arthritis, unspecified (UKB) | 296 | 65,160 | 1.19 [0.95, 1.48] | 0.122 |
| M06.0 - Seronegative rheumatoid arthritis (UKB) | 277 | 65,160 | 1.09 [0.87, 1.38] | 0.458 |
| M06.00 - Seronegative rheumatoid arthritis, unspecified site (UKB) | 238 | 65,160 | 1.04 [0.81, 1.35] | 0.714 |
| M06.9 - Rheumatoid arthritis, unspecified (UKB) | 3,348 | 65,160 | 1.04 [0.97, 1.12] | 0.260 |
| M06.90 - Rheumatoid arthritis, unspecified multiple sites (UKB) | 1,356 | 65,160 | 1.14 [1.02, 1.26] | 0.017 |
| M06.91 - Rheumatoid arthritis, unspecified shoulder region (UKB) | 128 | 65,160 | 1.22 [0.88, 1.70] | 0.239 |
| M06.92 - Rheumatoid arthritis, unspecified upper arm (UKB) | 60 | 65,160 | 1.64 [1.06, 2.53] | 0.027 |
| M06.93 - Rheumatoid arthritis, unspecified of the forearm (UKB) | 87 | 65,160 | 0.81 [0.51, 1.27] | 0.354 |
| M06.94 - Rheumatoid arthritis, unspecified hand (UKB) | 256 | 65,160 | 1.20 [0.95, 1.51] | 0.137 |
| M06.95 - Rheumatoid arthritis, unspecified pelvic region and thigh (UKB) | 98 | 65,160 | 1.08 [0.73, 1.60] | 0.692 |
| M06.96 - Rheumatoid arthritis, unspecified lower leg (UKB) | 255 | 65,160 | 1.14 [0.89, 1.45] | 0.291 |
| M06.97 - Rheumatoid arthritis, unspecified ankle and foot (UKB) | 211 | 65,160 | 1.08 [0.82, 1.41] | 0.588 |
| M75.0 - Adhesive capsulitis of shoulder (UKB) | 2,023 | 65,160 | 0.99 [0.91, 1.09] | 0.849 |
| Skeletal fibrosis (UKB) | 6,516 | 65,160 | 1.04 [0.99, 1.10] | 0.108 |
| Rheumatoid arthritis (Ishigaki et al. 2022) | 22,350 | 74,823 | 1.12 [1.08, 1.16] | 2.123e-10 |
| **Skeletal fibrosis (Meta-analysis)** | **28,866** | **139,983** | **1.09 [1.06, 1.13]** | **2.216e-09** |

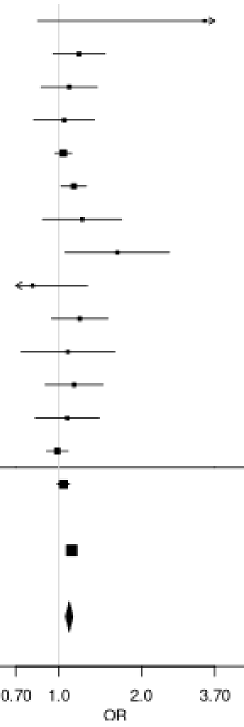

**b) Skin**

| Disease | Case | Control | OR [95% CI] | P |
|---|---|---|---|---|
| L90.5 - Scar conditions and fibrosis of skin (UKB) | 2,789 | 65,930 | 0.92 [0.85, 0.99] | 0.035 |
| L94.0 - Localized scleroderma [morphea] (UKB) | 66 | 65,930 | 0.77 [0.45, 1.32] | 0.345 |
| M32.0 - Drug-induced systemic lupus erythematosus (UKB) | 12 | 65,930 | 1.14[0.39, 3.34] | 0.806 |
| M32.1 - Systemic lupus erythematosus with organ or system involvement (UKB) | 59 | 65,930 | 1.33 [0.83, 2.14] | 0.239 |
| M32.9 - Systemic lupus erythematosus, unspecified (UKB) | 408 | 65,930 | 1.10 [0.90, 1.33] | 0.357 |
| M33.1 - Other dermatomyositis (UKB) | 48 | 65,930 | 1.29 [0.76, 2.18] | 0.339 |
| M35.4 - Diffuse (eosinophilic) fasciitis (UKB) | 7 | 65,930 | 1.58 [0.44, 5.66] | 0.484 |
| M36.0 - Dermato(poly)myositis in neoplastic disease (UKB) | 5 | 65,930 | 0.64 [0.08, 5.06] | 0.673 |
| M72.0 - Palmar fascial fibromatosis [Dupuytren] (UKB) | 3,316 | 65,930 | 1.02 [0.95, 1.10] | 0.530 |
| Skin fibrosis (UKB) | 6,593 | 65,930 | 0.98 [0.93, 1.04] | 0.518 |
| Dupuytren's disease (Riesmeijer et al. 2024) | 7,584 | 28,343 | 1.07 [1.01, 1.14] | 0.036 |
| Systemic lupus erythematosus (Bentham et al. 2015) | 5,201 | 9,066 | 1.28 [1.19, 1.37] | 2.729e-12 |
| Systemic lupus erythematosus (Julia et al. 2018) | 907 | 1,524 | 1.20 [0.99, 1.45] | 0.061 |
| **Skin fibrosis (Meta-analysis)** | **20,285** | **104,863** | **1.08 [1.05, 1.12]** | **7.195e-06** |

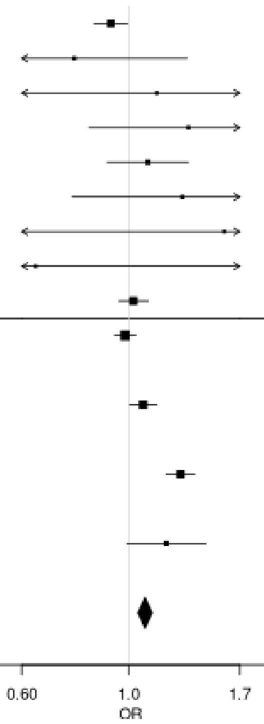

**c) Systemic**

| Disease | Case | Control | OR [95% CI] | P |
|---|---|---|---|---|
| E85.8 - Other amyloidosis (UKB) | 20 | 16,200 | 1.02 [0.42, 2.44] | 0.971 |
| H19.3 - Keratitis and keratoconjunctivitis in other diseases classified elsewhere (sicca) (UKB) | 31 | 16,200 | 1.23 [0.64, 2.39] | 0.537 |
| J98.5 - Diseases of mediastinum, not elsewhere classified (UKB) | 84 | 16,200 | 1.21 [0.80, 1.83] | 0.358 |
| M34.0 - Progressive systemic sclerosis (UKB) | 13 | 16,200 | 0.48 [0.11, 2.03] | 0.316 |
| M34.1 - CR(E)ST syndrome (UKB) | 77 | 16,200 | 0.94 [0.59, 1.49] | 0.778 |
| M34.2 - Systemic sclerosis induced by drugs and chemicals (UKB) | 3 | 16,200 | 1.11 [0.13, 9.54] | 0.922 |
| M34.8 - Other forms of systemic sclerosis (UKB) | 63 | 16,200 | 0.92 [0.55, 1.54] | 0.749 |
| M34.9 - Systemic sclerosis, unspecified (UKB) | 194 | 16,200 | 0.81[0.60, 1.11] | 0.189 |
| M35.0 - Sicca syndrome (Sjögren's syndrome) (UKB) | 688 | 16,200 | 1.03 [0.88, 1.20] | 0.698 |
| T85.8 - Other complications of internal prosthetic devices, implants and grafts, not elsewhere classified (UKB) | 615 | 16,200 | 1.01 [0.86, 1.19] | 0.908 |
| Systemic fibrosis (UKB) | 1,620 | 16,200 | 1.01 [0.91, 1.12] | 0.892 |
| Sjögren's syndrome (Taylor et al. 2017) | 585 | 1,546 | 1.26 [1.07, 1.49] | 0.007 |
| **Systemic fibrosis (Meta-analysis)** | **11,300** | **35,330** | **1.07 [0.98, 1.17]** | **0.121** |

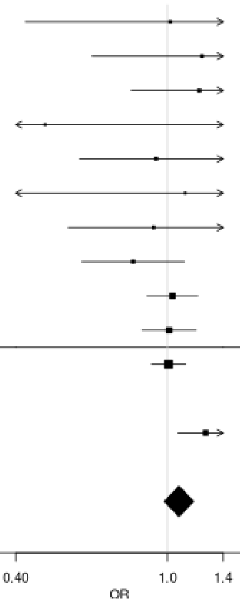

**Supplementary Figure 13: Region plot of novel replicated variants**

Variant rs113477191 was associated with intestinal-pancreatic fibrosis and variant rs2580350 was associated with urinary fibrosis. Each point on the plots represents a variant with chromosomal position (GRCh37) on x axis and $-\log_{10}$(p value) on y axis. The sentinel variant is coloured purple with other variants coloured by strength of LD with the sentinel variant based on the 1000 genomes EUR population. Recombination rates shown by the blue lines and positions of genes shown at bottom.

**a) rs113477191 (Intestinal-pancreatic)**

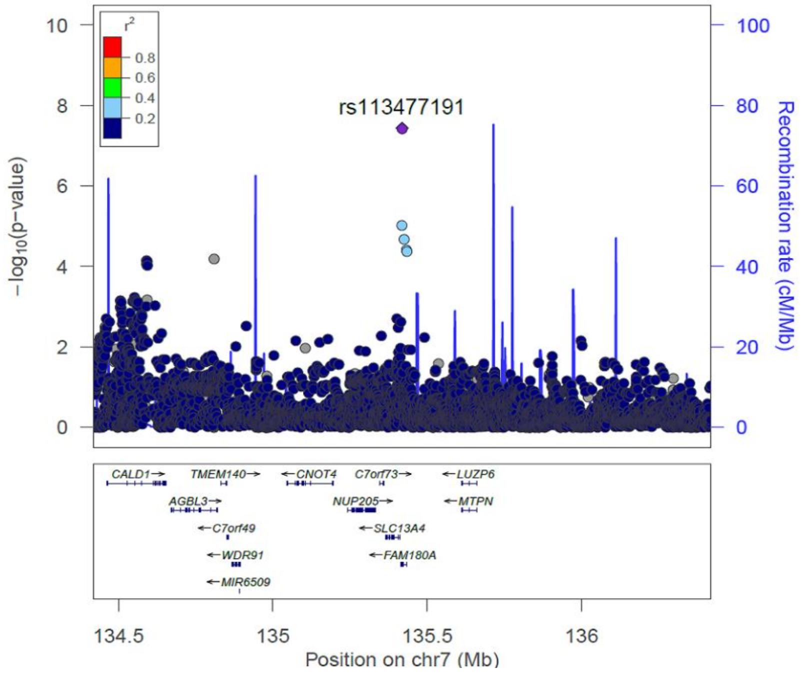

**b) rs2580350 (Urinary)**

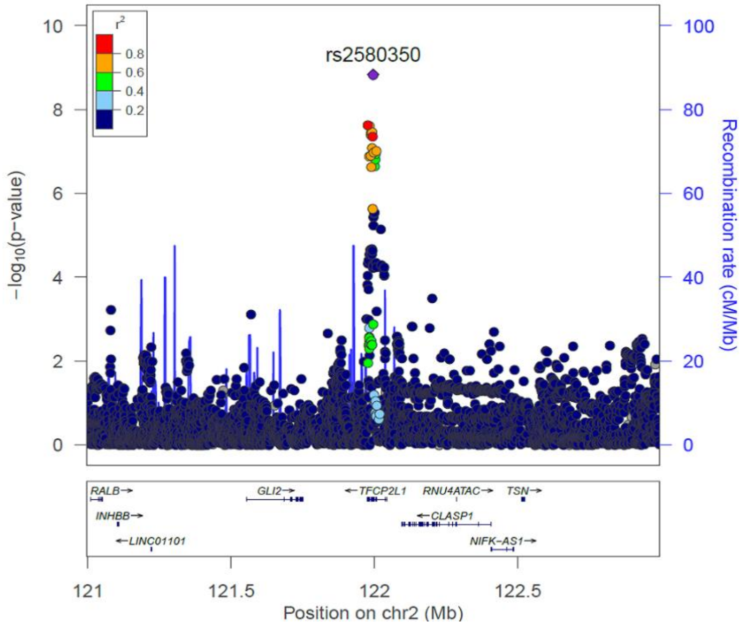

**Supplementary Figure 14: Genetic correlation heatmap for ‘always fibrotic’ case definition**
Genome-wide genetic correlations were calculated using LDSC using GWAS summary statistics from the European ‘always fibrotic’ analyses. Genetic correlations are shown and colours show the strength of the correlation. No correlations were nominally significant ($p<0.05$).

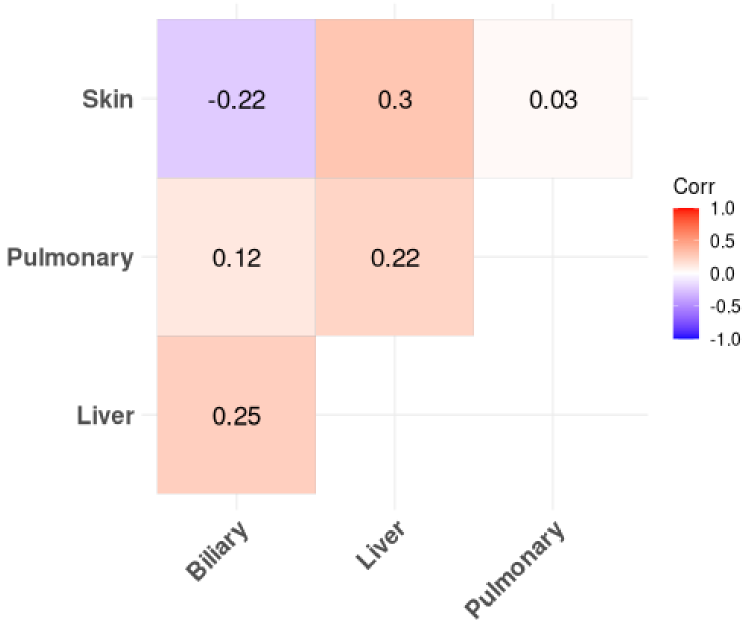

#### Supplementary Figure 15: Genetic and phenotypic correlation heatmap for UK Biobank analyses

Genome-wide genetic correlations were calculated using LDSC using GWAS summary statistics from the European ‘broadly fibrotic’ analyses only using UK Biobank. Genetic correlations are shown and colours show the strength of the correlation. ** Denotes correlations significant after Bonferroni correction ($p<9.09\times10^{-4}$). * Denotes correlations with $p<0.05$.

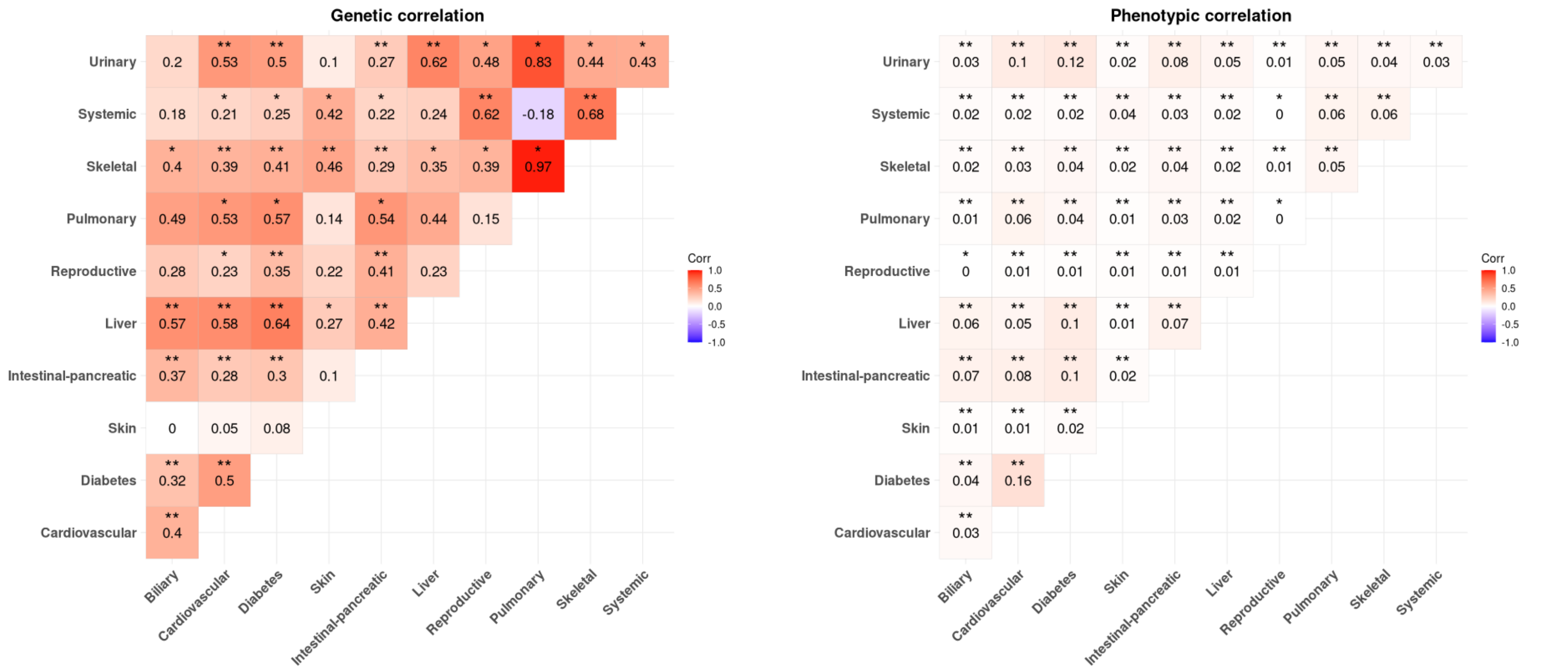

**Supplementary Figure 16: Jaccard index for sample overlap in UK Biobank between organs**

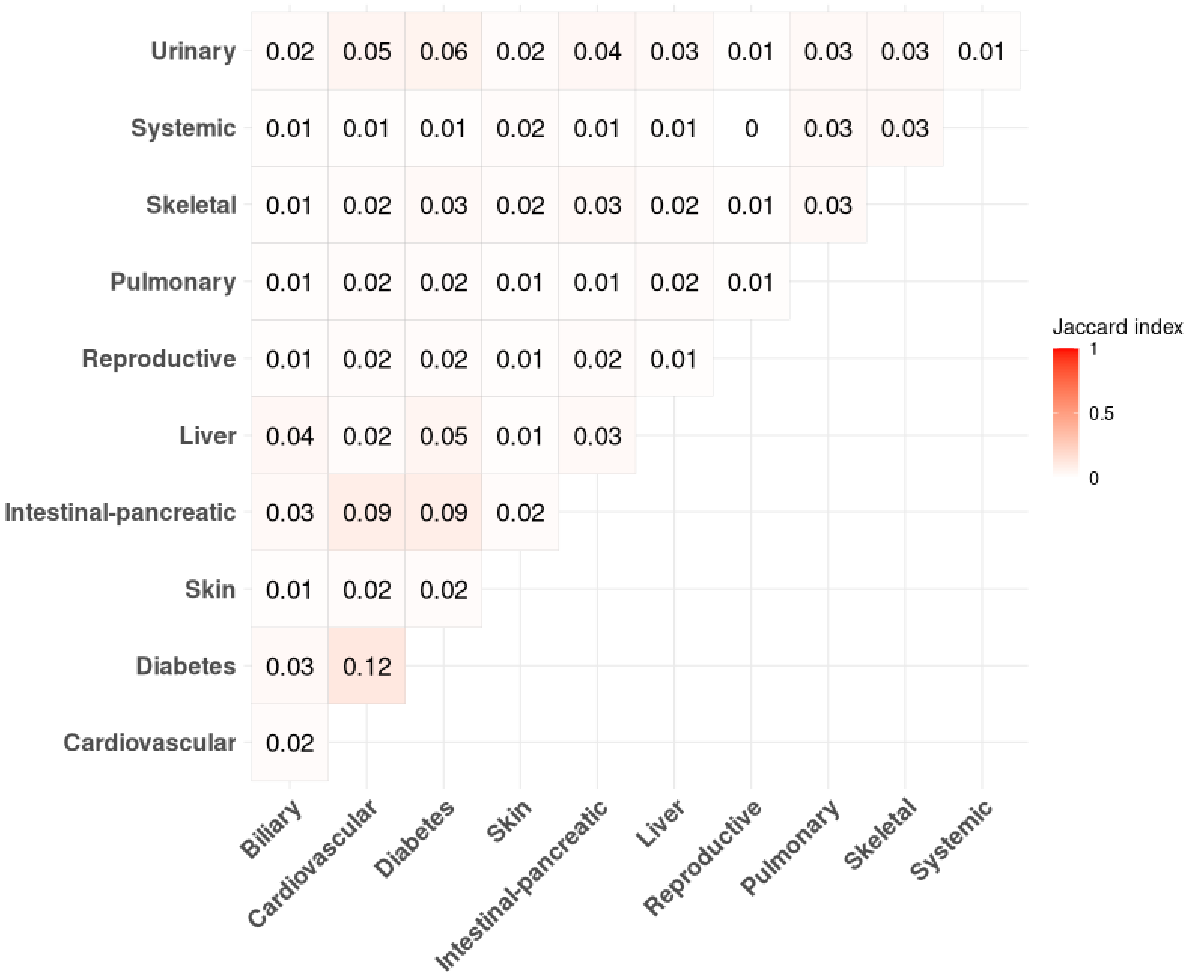

**Supplementary Figure 17: Upset plot showing overlap of cases between different 'broadly fibrotic' organs in UK Biobank**

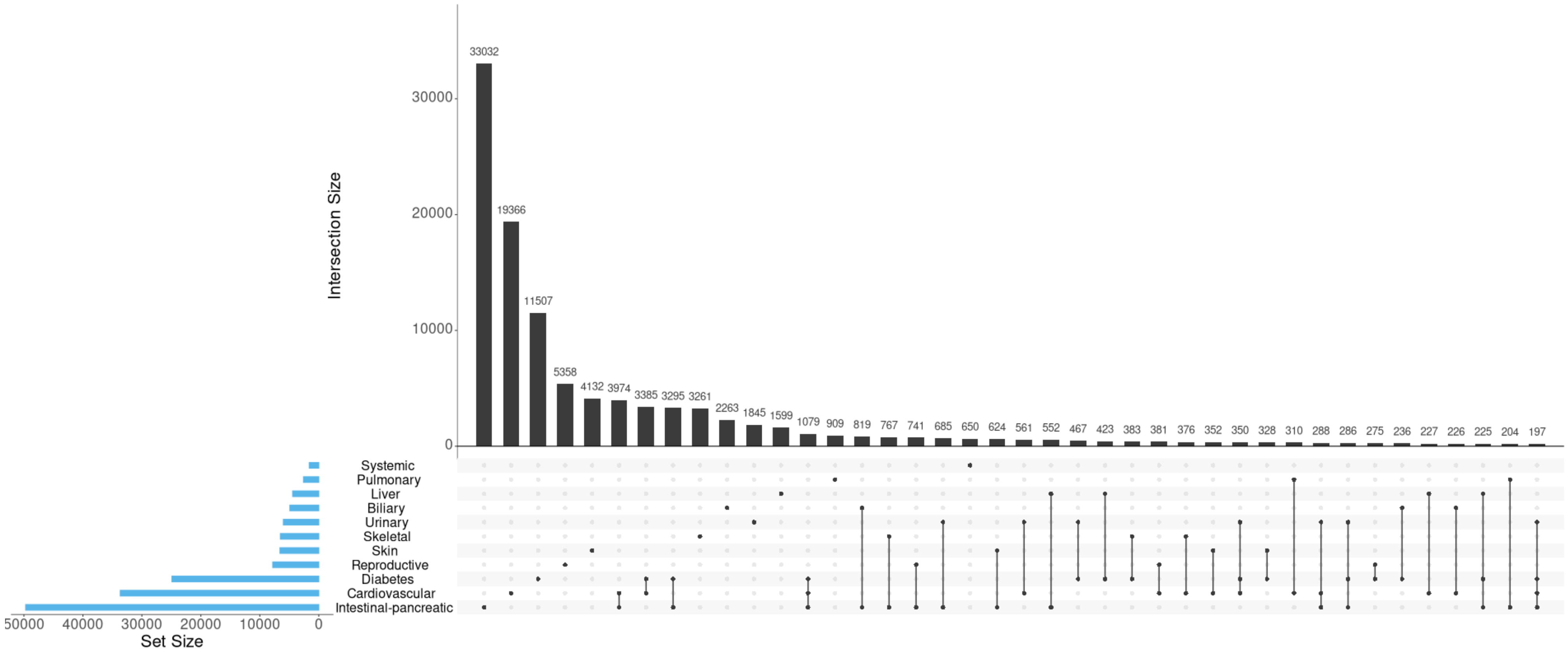

**Supplementary Figure 18: Partial genetic correlation results**

**a) Biliary**

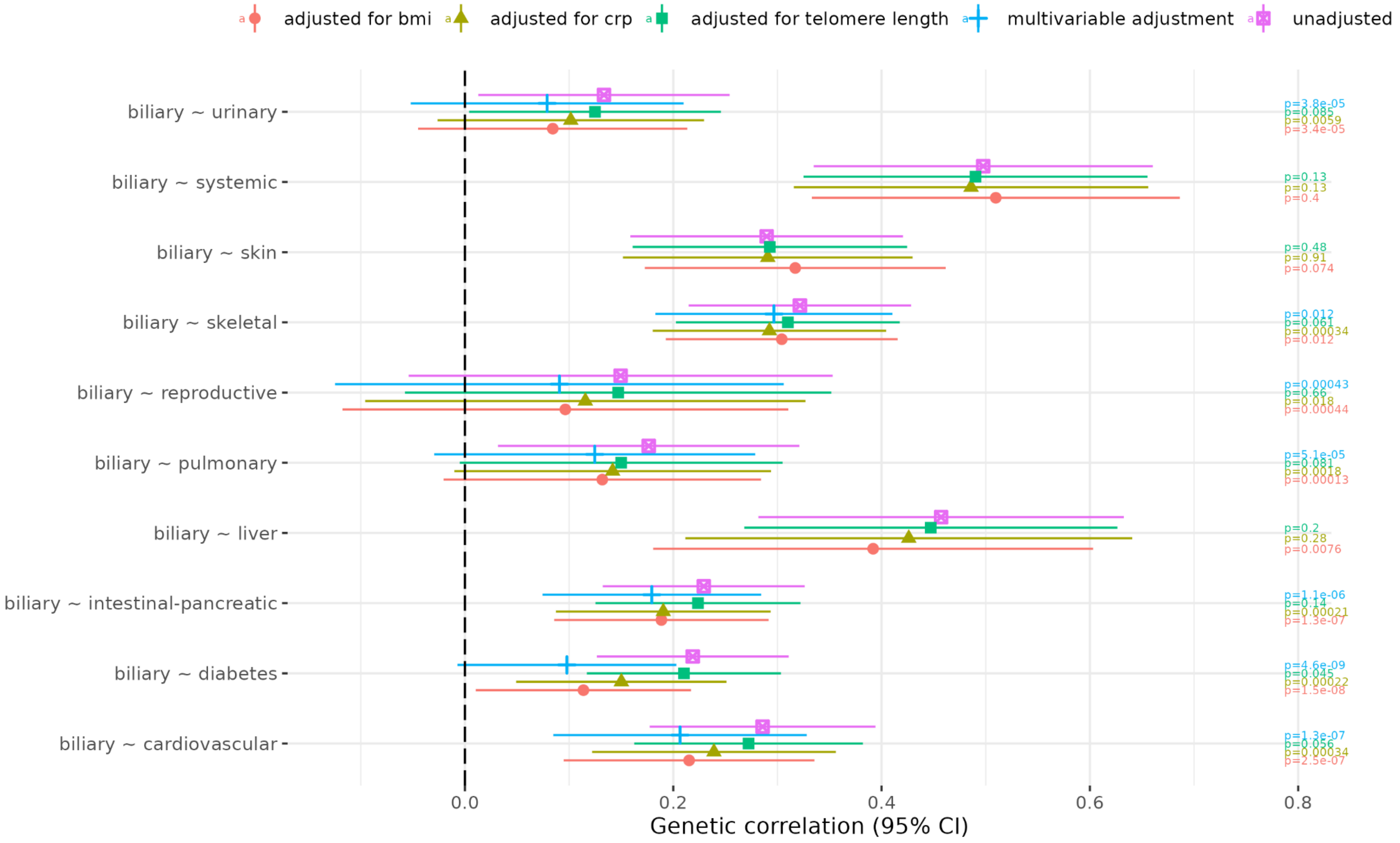

**b) Cardiovascular**

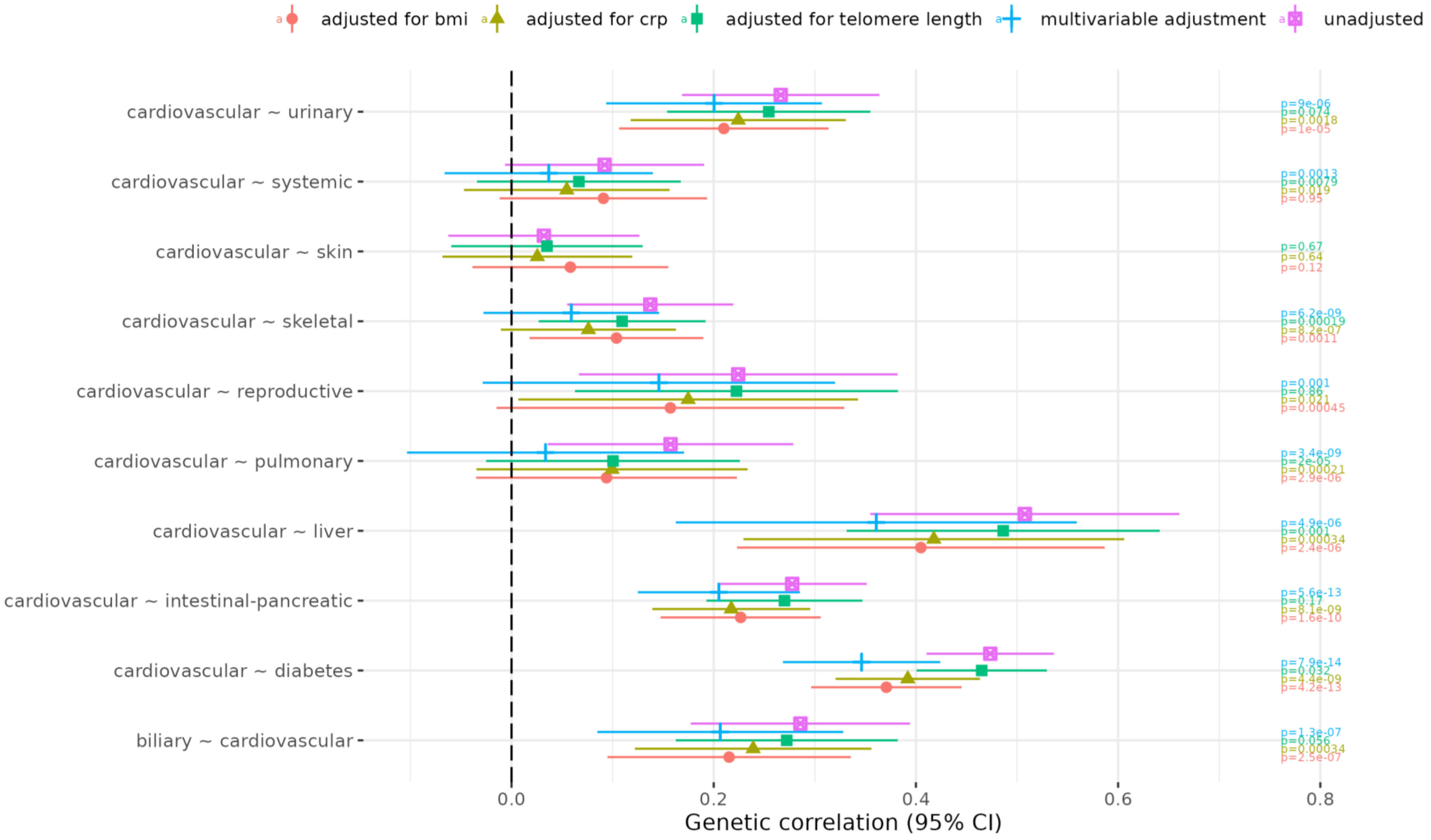

**c) Diabetes**

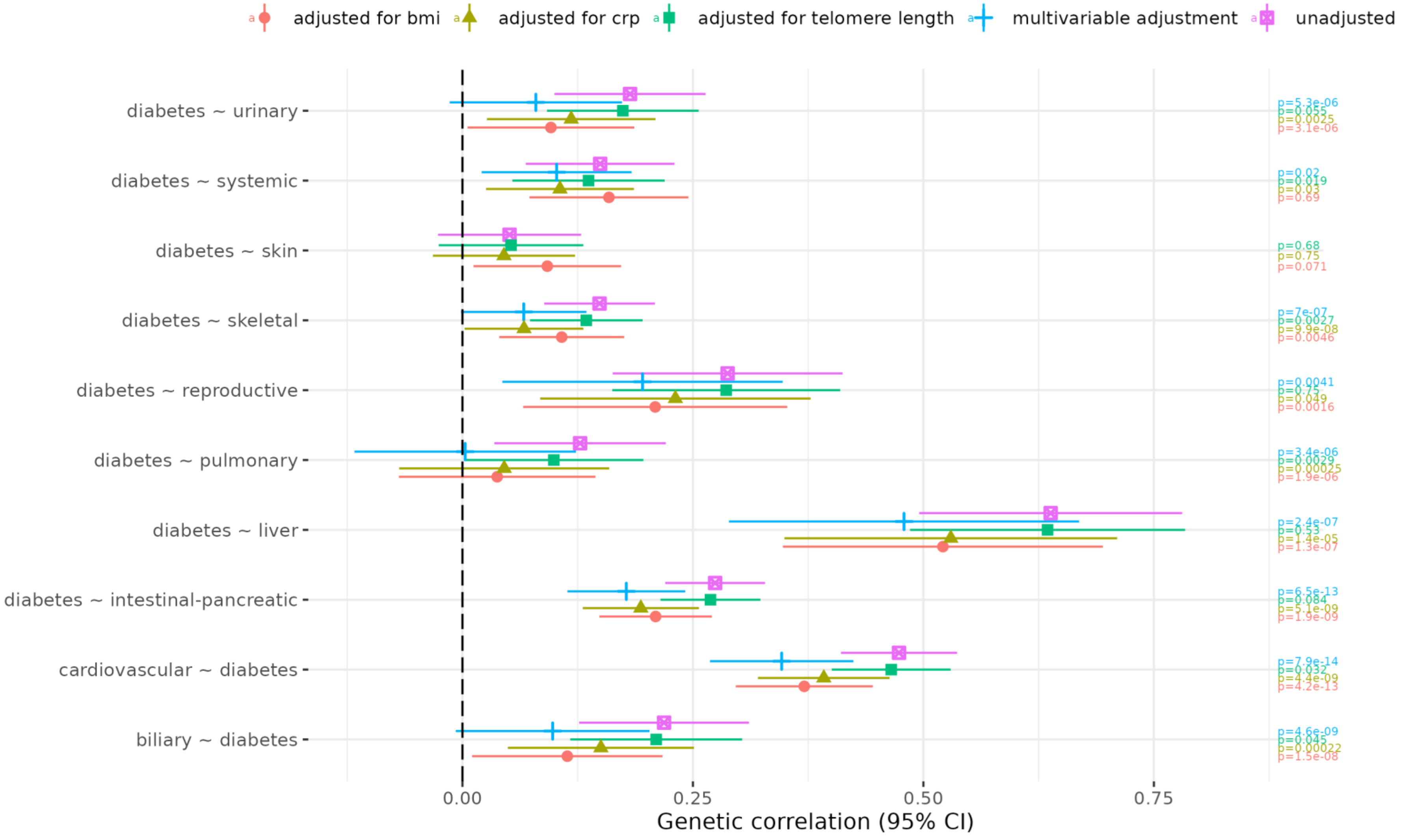

adjusted for bmi
adjusted for crp
adjusted for telomere length
multivariable adjustment
unadjusted
diabetes ~ urinary
diabetes ~ systemic
diabetes ~ skin
diabetes ~ skeletal
diabetes ~ reproductive
diabetes ~ pulmonary
diabetes ~ liver
diabetes ~ intestinal-pancreatic
cardiovascular ~ diabetes
biliary ~ diabetes
0.00
0.25
0.50
0.75
Genetic correlation (95% CI)
p=5.3e-06
p=0.055
p=0.0025
p=3.1e-06
p=0.02
p=0.019
p=0.03
p=0.69
p=0.68
p=0.75
p=0.071
p=7e-07
p=0.0027
p=9.9e-08
p=0.0046
p=0.0041
p=0.75
p=0.049
p=0.0016
p=3.4e-06
p=0.0029
p=0.00025
p=1.9e-06
p=2.4e-07
p=0.53
p=1.4e-05
p=1.3e-07
p=6.5e-13
p=0.084
p=5.1e-09
p=1.9e-09
p=7.9e-14
p=0.032
p=4.4e-09
p=4.2e-13
p=4.6e-09
p=0.045
p=0.00022
p=1.5e-08

**d) Intestinal-pancreas**

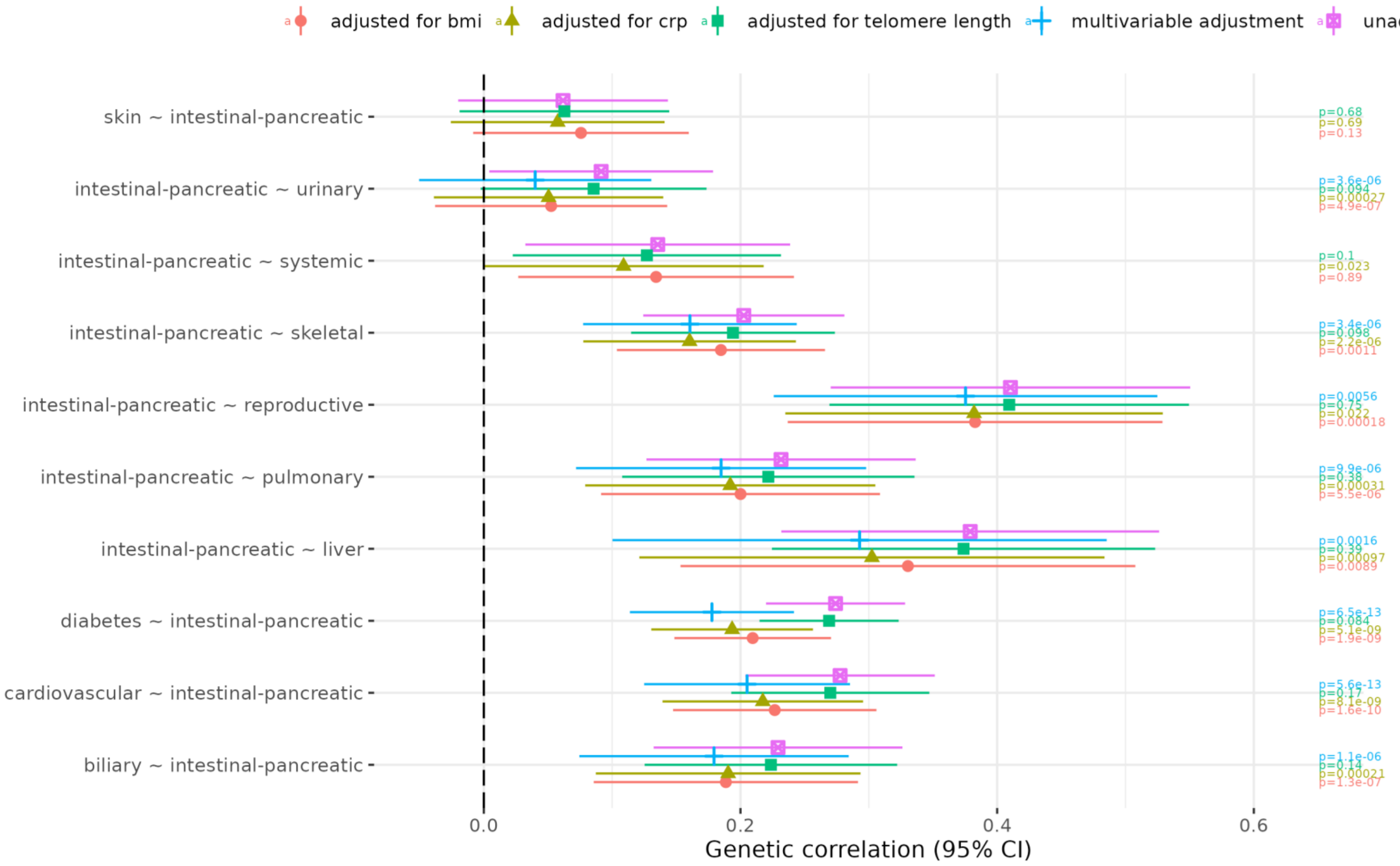

**e) Liver**

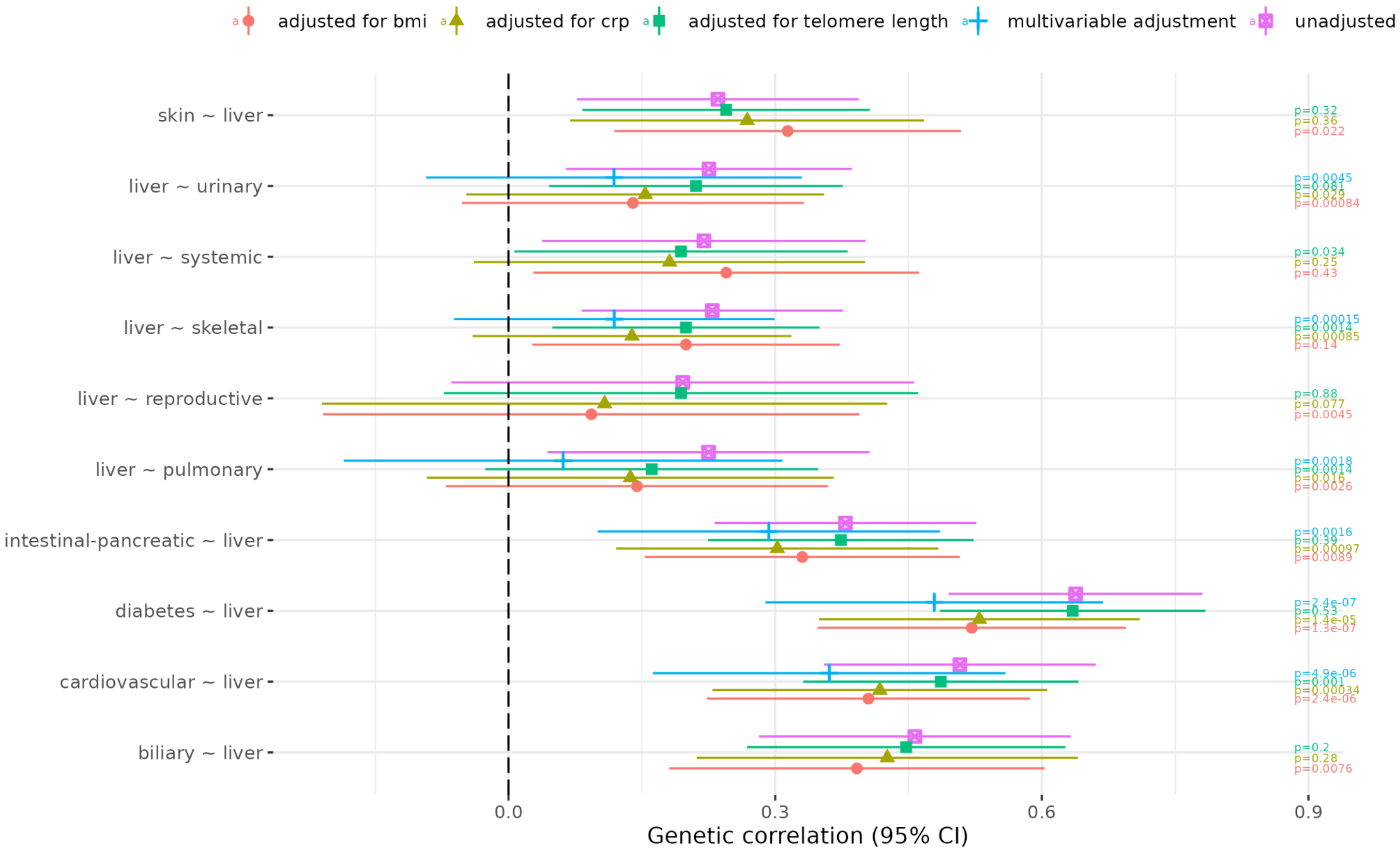
adjusted for bmi
adjusted for crp
adjusted for telomere length
multivariable adjustment
unadjusted
skin ~ liver
liver ~ urinary
liver ~ systemic
liver ~ skeletal
liver ~ reproductive
liver ~ pulmonary
intestinal-pancreatic ~ liver
diabetes ~ liver
cardiovascular ~ liver
biliary ~ liver
p=0.32
p=0.36
p=0.022
p=0.0045
p=0.081
p=0.029
p=0.00084
p=0.034
p=0.25
p=0.43
p=0.00015
p=0.0014
p=0.00085
p=0.14
p=0.88
p=0.077
p=0.0045
p=0.0018
p=0.0014
p=0.016
p=0.0026
p=0.0016
p=0.39
p=0.00097
p=0.0089
p=2.4e-07
p=0.53
p=1.4e-05
p=1.3e-07
p=4.9e-06
p=0.001
p=0.00034
p=2.4e-06
p=0.2
p=0.28
p=0.0076
0.0
0.3
0.6
0.9
Genetic correlation (95% CI)

**f) Pulmonary**

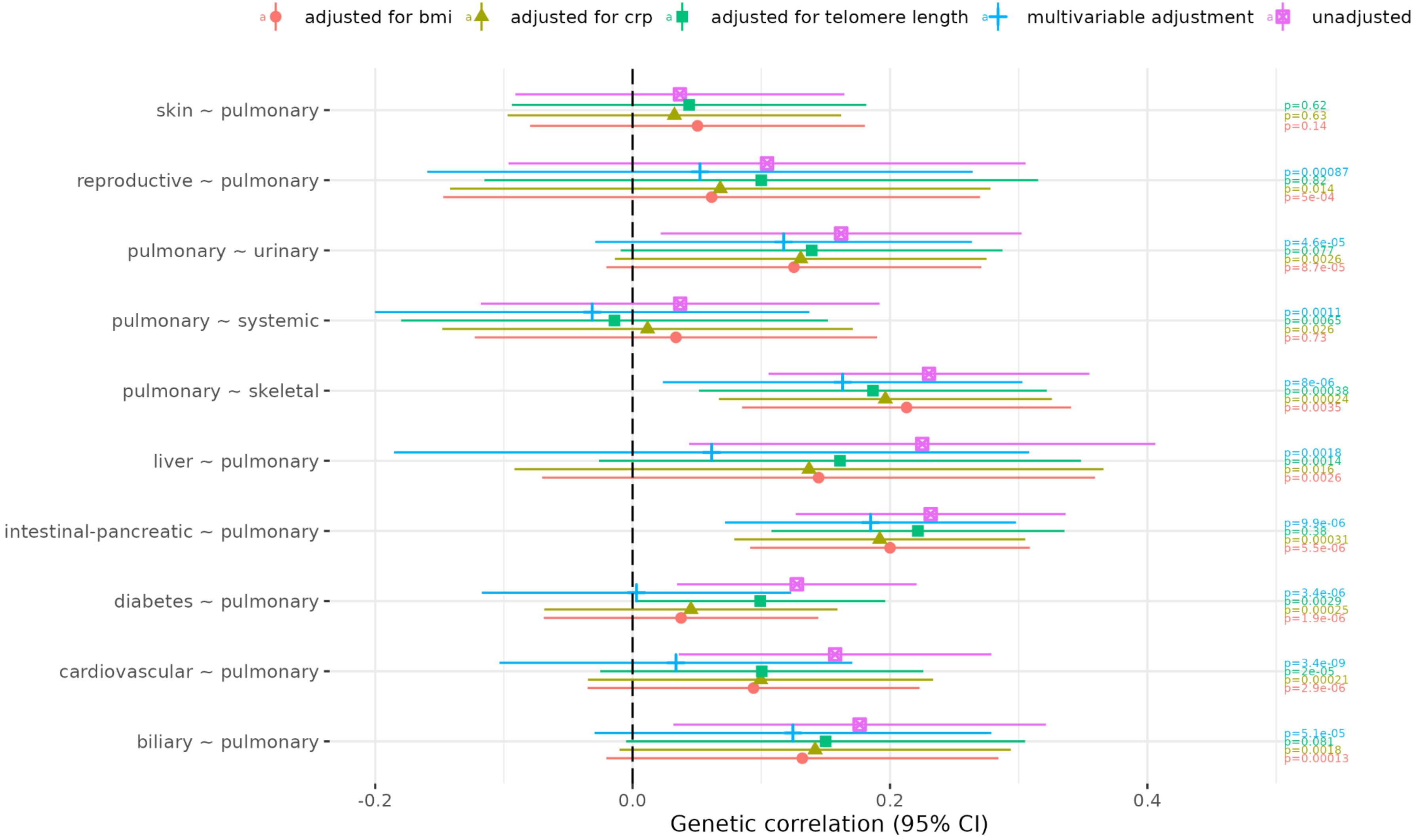

adjusted for bmi
adjusted for crp
adjusted for telomere length
multivariable adjustment
unadjusted
skin ~ pulmonary
reproductive ~ pulmonary
pulmonary ~ urinary
pulmonary ~ systemic
pulmonary ~ skeletal
liver ~ pulmonary
intestinal-pancreatic ~ pulmonary
diabetes ~ pulmonary
cardiovascular ~ pulmonary
biliary ~ pulmonary
p=0.62
p=0.63
p=0.14
p=0.00087
p=0.82
p=0.014
p=5e-04
p=4.6e-05
p=0.077
p=0.0026
p=8.7e-05
p=0.0011
p=0.0065
p=0.026
p=0.73
p=8e-06
p=0.00038
p=0.00024
p=0.0035
p=0.0018
p=0.0014
p=0.016
p=0.0026
p=9.9e-06
p=0.38
p=0.00031
p=5.5e-06
p=3.4e-06
p=0.0029
p=0.00025
p=1.9e-06
p=3.4e-09
p=2e-05
p=0.00021
p=2.9e-06
p=5.1e-05
p=0.081
p=0.0018
p=0.00013
-0.2
0.0
0.2
0.4
Genetic correlation (95% CI)

**g) Reproductive**

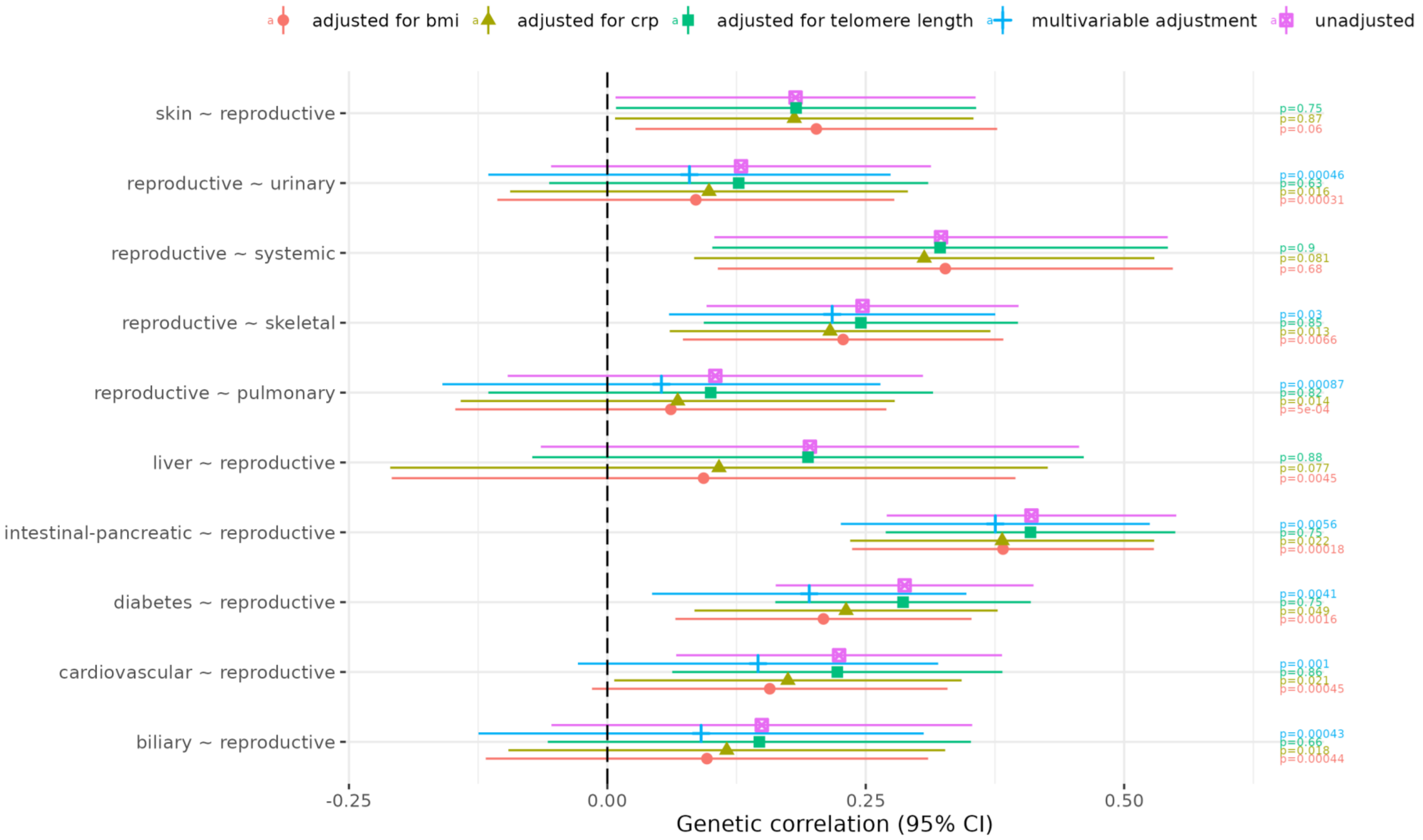

**h) Skeletal**

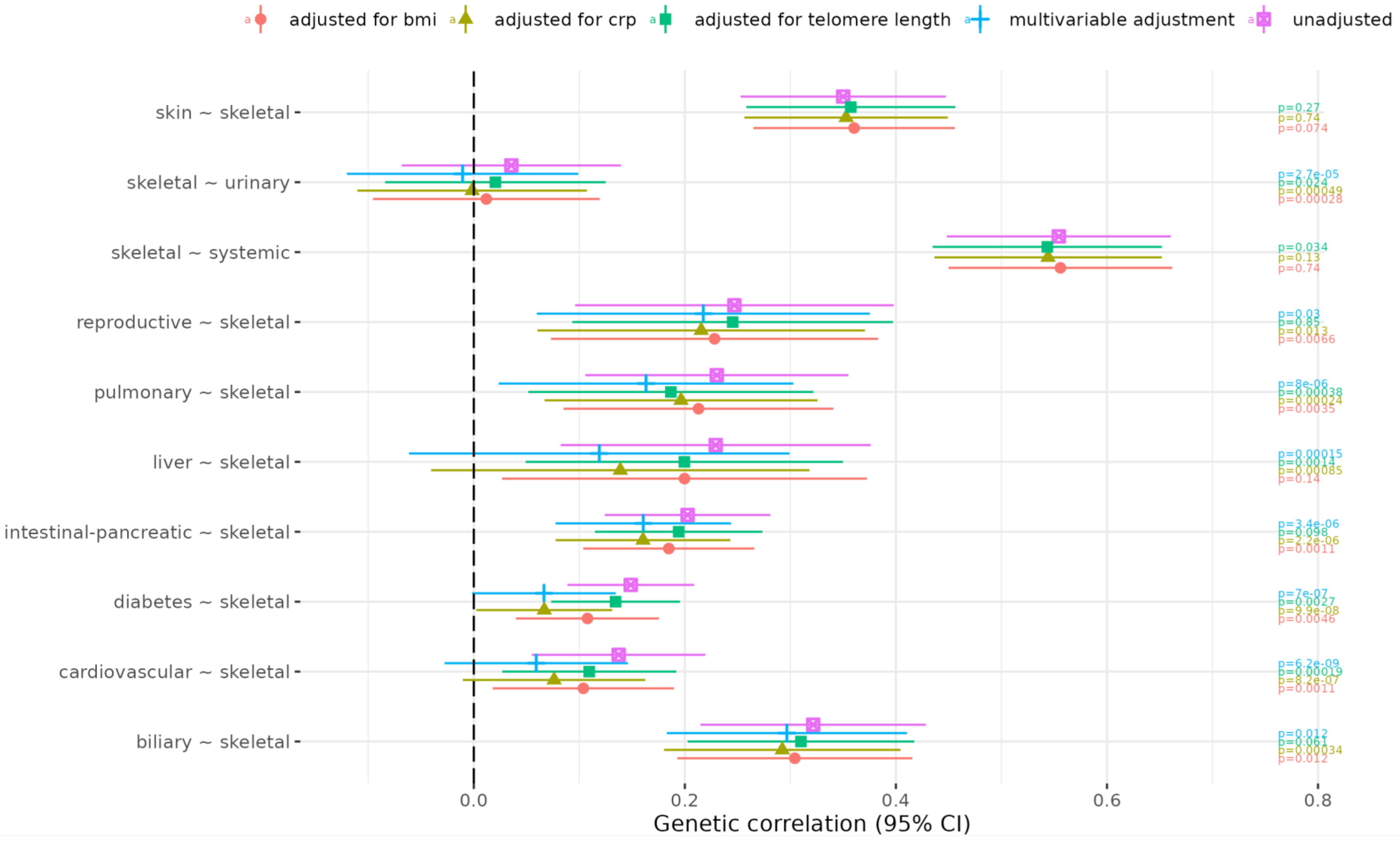

adjusted for bmi
adjusted for crp
adjusted for telomere length
multivariable adjustment
unadjusted
skin ~ skeletal
skeletal ~ urinary
skeletal ~ systemic
reproductive ~ skeletal
pulmonary ~ skeletal
liver ~ skeletal
intestinal-pancreatic ~ skeletal
diabetes ~ skeletal
cardiovascular ~ skeletal
biliary ~ skeletal
p=0.27
p=0.74
p=0.074
p=2.7e-05
p=0.024
p=0.00049
p=0.00028
p=0.034
p=0.13
p=0.74
p=0.03
p=0.85
p=0.013
p=0.0066
p=8e-06
p=0.00038
p=0.00024
p=0.0035
p=0.00015
p=0.0014
p=0.00085
p=0.14
p=3.4e-06
p=0.098
p=2.2e-06
p=0.0011
p=7e-07
p=0.0027
p=9.9e-08
p=0.0046
p=6.2e-09
p=0.00019
p=8.2e-07
p=0.0011
p=0.012
p=0.061
p=0.00034
p=0.012
0.0
0.2
0.4
0.6
0.8
Genetic correlation (95% CI)

**i) Skin**

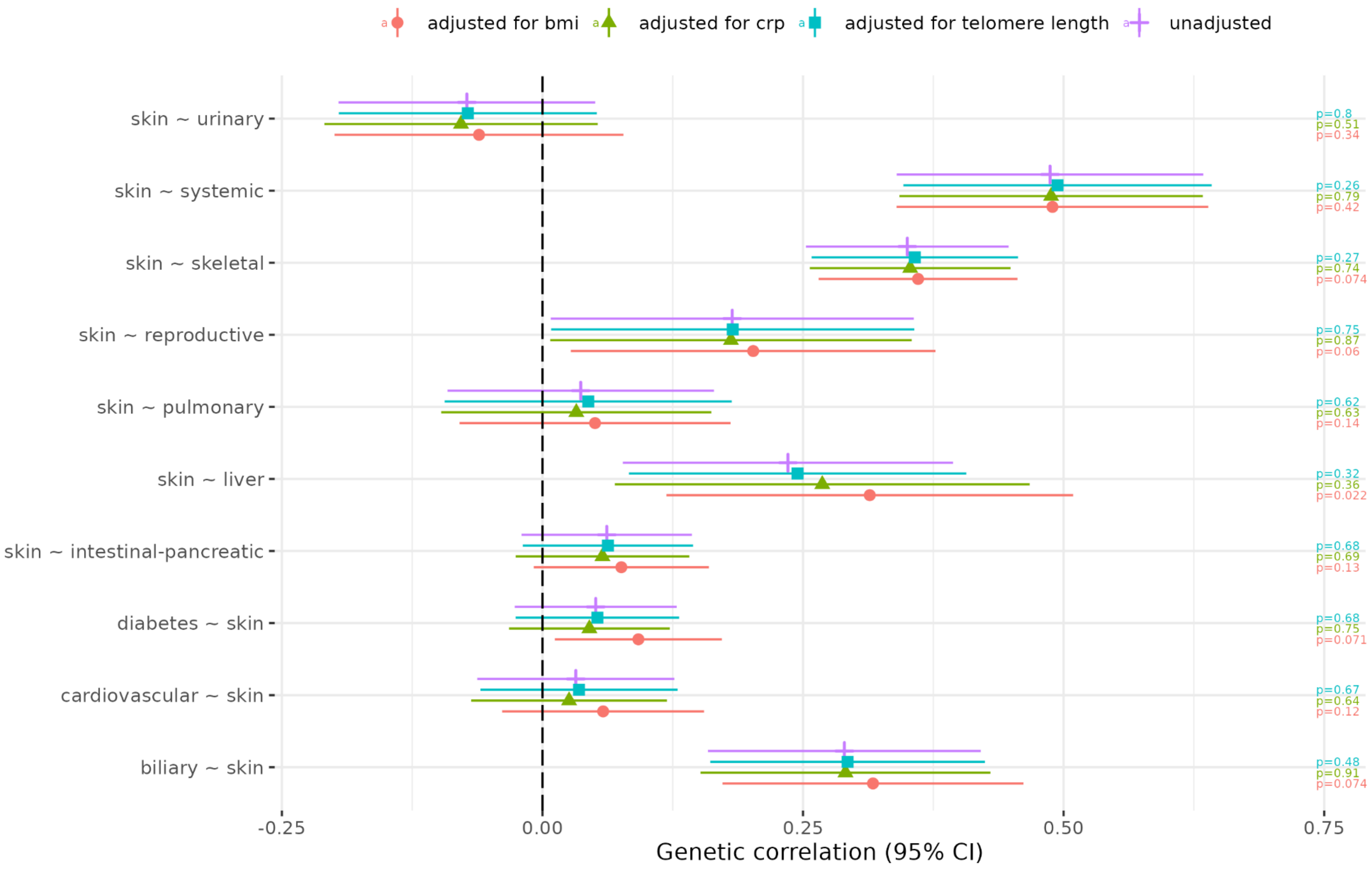

adjusted for bmi
adjusted for crp
adjusted for telomere length
unadjusted
skin ~ urinary
skin ~ systemic
skin ~ skeletal
skin ~ reproductive
skin ~ pulmonary
skin ~ liver
skin ~ intestinal-pancreatic
diabetes ~ skin
cardiovascular ~ skin
biliary ~ skin
p=0.8
p=0.51
p=0.34
p=0.26
p=0.79
p=0.42
p=0.27
p=0.74
p=0.074
p=0.75
p=0.87
p=0.06
p=0.62
p=0.63
p=0.14
p=0.32
p=0.36
p=0.022
p=0.68
p=0.69
p=0.13
p=0.68
p=0.75
p=0.071
p=0.67
p=0.64
p=0.12
p=0.48
p=0.91
p=0.074
-0.25
0.00
0.25
0.50
0.75
Genetic correlation (95% CI)

**j) Systemic**

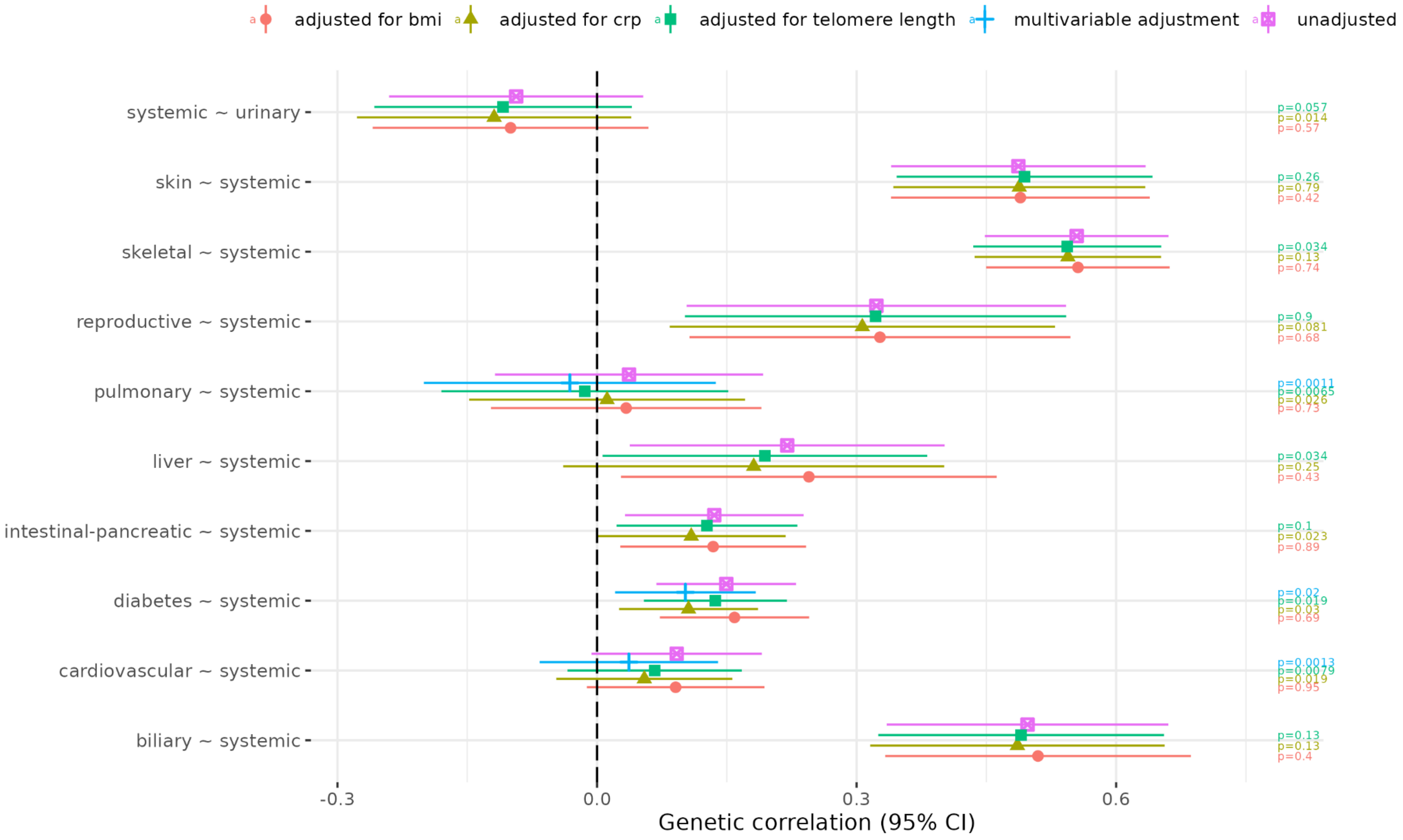

adjusted for bmi
adjusted for crp
adjusted for telomere length
multivariable adjustment
unadjusted
systemic ~ urinary
skin ~ systemic
skeletal ~ systemic
reproductive ~ systemic
pulmonary ~ systemic
liver ~ systemic
intestinal-pancreatic ~ systemic
diabetes ~ systemic
cardiovascular ~ systemic
biliary ~ systemic
p=0.057
p=0.014
p=0.57
p=0.26
p=0.79
p=0.42
p=0.034
p=0.13
p=0.74
p=0.9
p=0.081
p=0.68
p=0.0011
p=0.0065
p=0.026
p=0.73
p=0.034
p=0.25
p=0.43
p=0.1
p=0.023
p=0.89
p=0.02
p=0.019
p=0.03
p=0.69
p=0.0013
p=0.0079
p=0.019
p=0.95
p=0.13
p=0.13
p=0.4
-0.3
0.0
0.3
0.6
Genetic correlation (95% CI)

**k) Urinary**

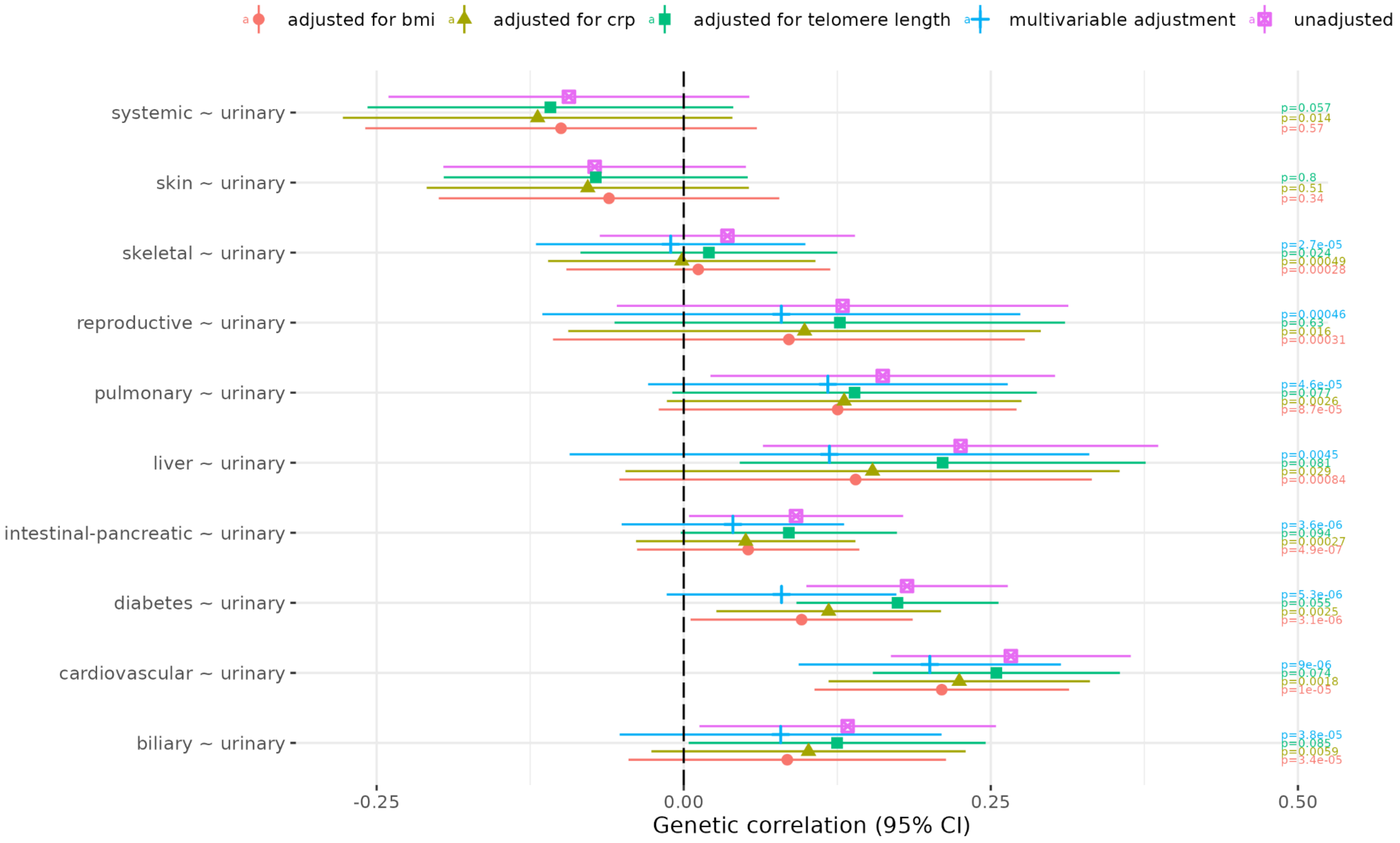
adjusted for bmi
adjusted for crp
adjusted for telomere length
multivariable adjustment
unadjusted
systemic ~ urinary
skin ~ urinary
skeletal ~ urinary
reproductive ~ urinary
pulmonary ~ urinary
liver ~ urinary
intestinal-pancreatic ~ urinary
diabetes ~ urinary
cardiovascular ~ urinary
biliary ~ urinary
p=0.057
p=0.014
p=0.57
p=0.8
p=0.51
p=0.34
p=2.7e-05
p=0.024
p=0.00049
p=0.00028
p=0.00046
p=0.63
p=0.016
p=0.00031
p=4.6e-05
p=0.077
p=0.0026
p=8.7e-05
p=0.0045
p=0.081
p=0.029
p=0.00084
p=3.6e-06
p=0.094
p=0.00027
p=4.9e-07
p=5.3e-06
p=0.055
p=0.0025
p=3.1e-06
p=9e-06
p=0.074
p=0.0018
p=1e-05
p=3.8e-05
p=0.085
p=0.0059
p=3.4e-05
-0.25
0.00
0.25
0.50
Genetic correlation (95% CI)

#### Supplementary Figure 19: Individual variant results in non-European ancestries

### a) rs6679356

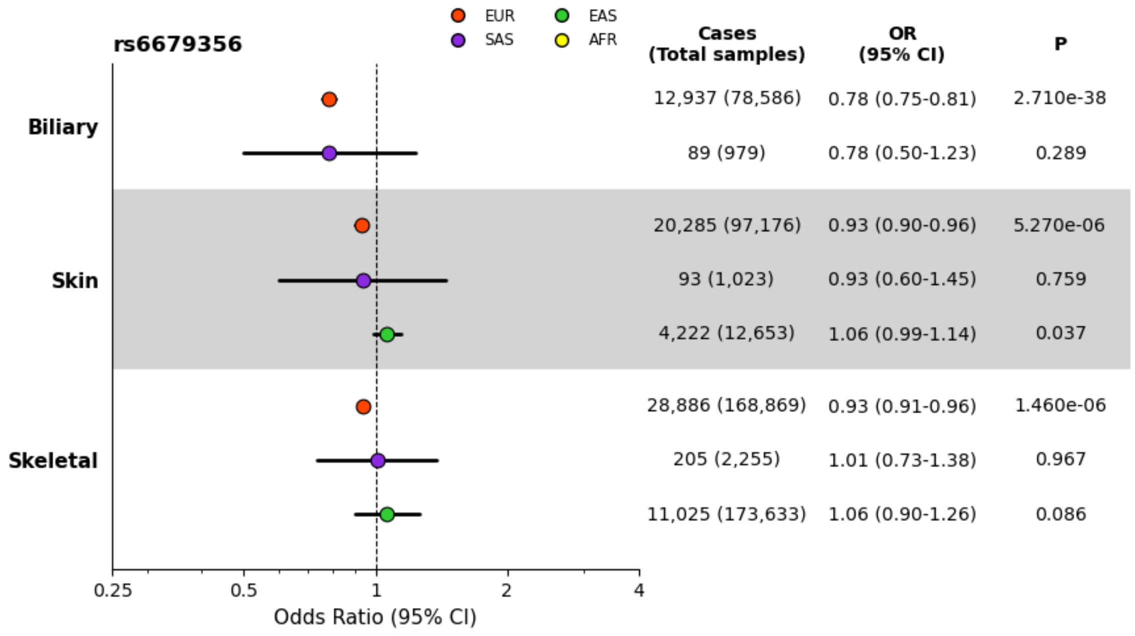

### b) rs2476601

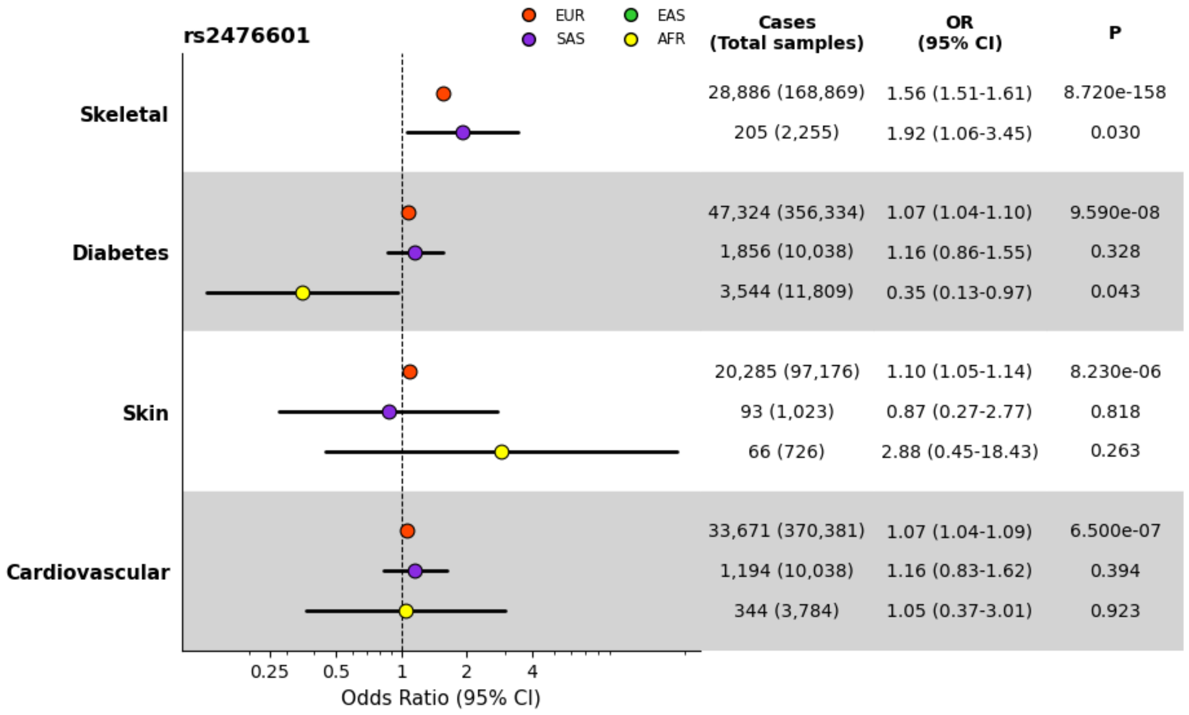

c) rs780093

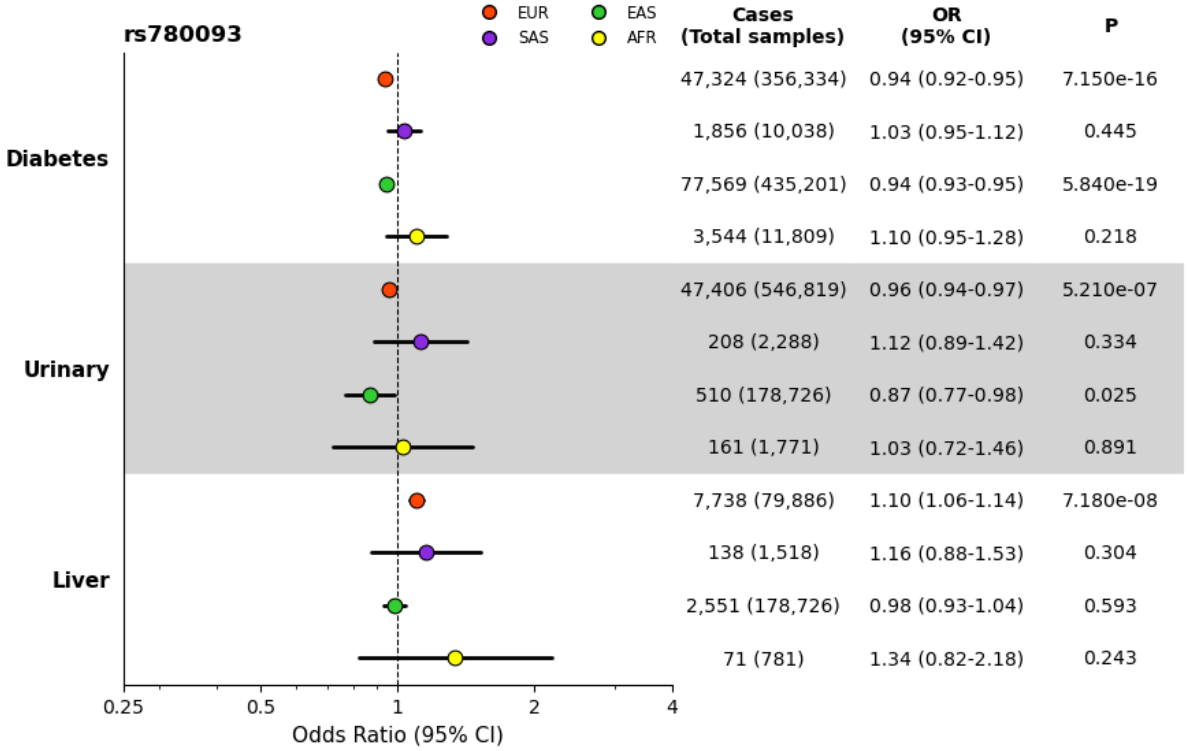

d) rs772106113

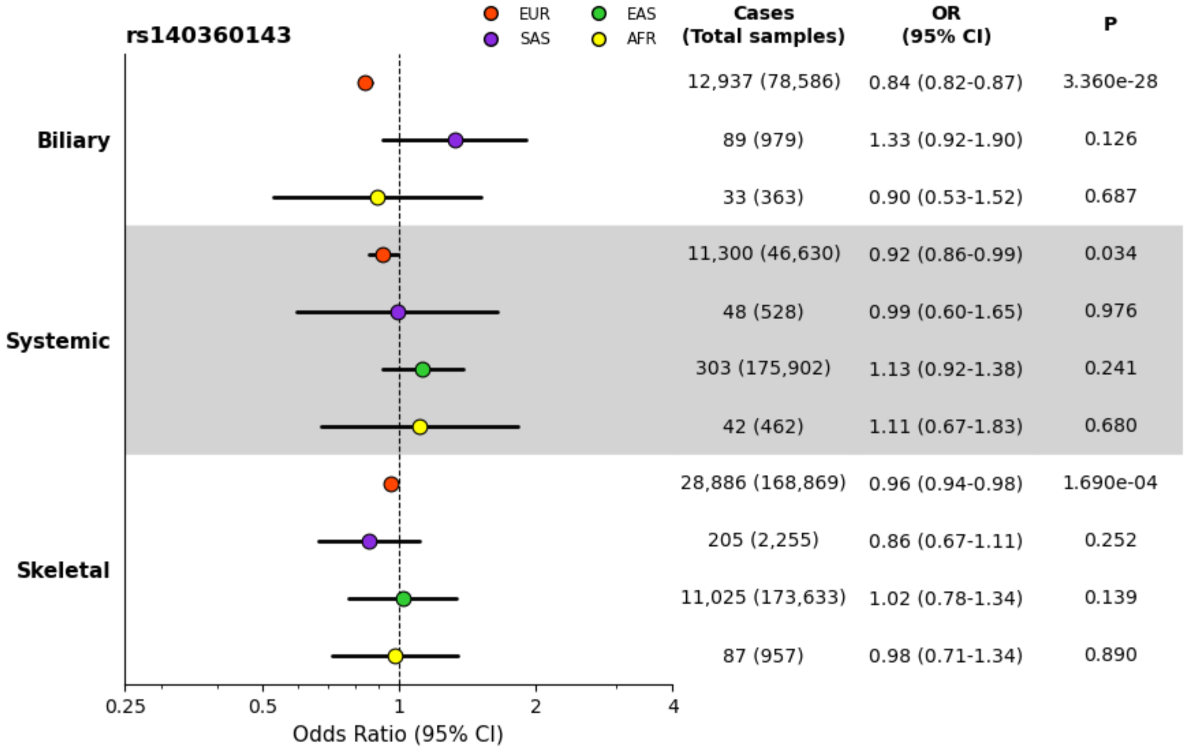

**e) rs13246321**

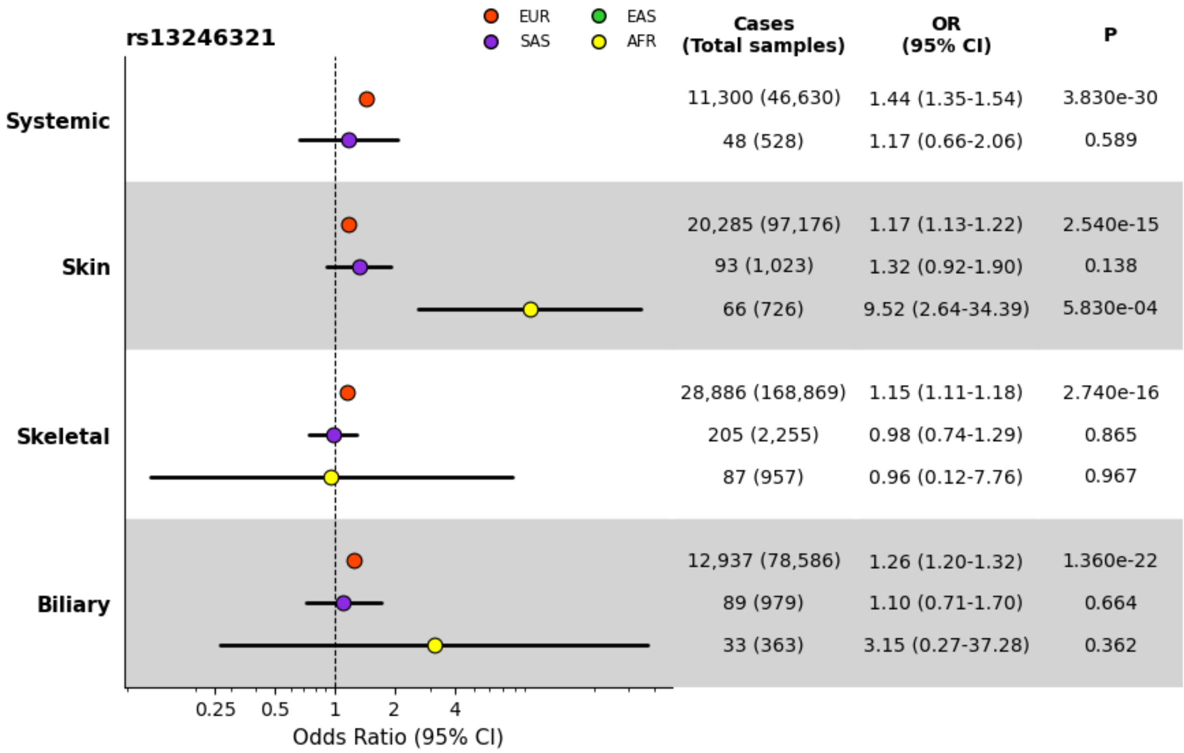

**f) rs2736340**

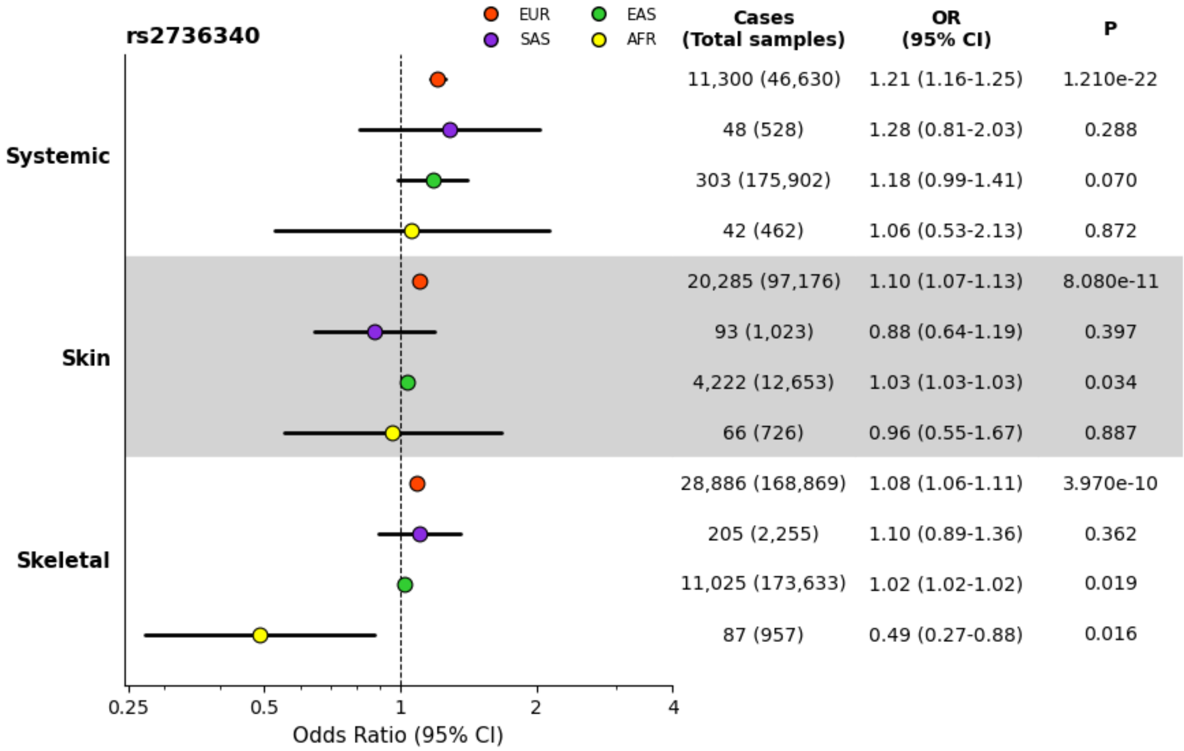

## g) rs7310615

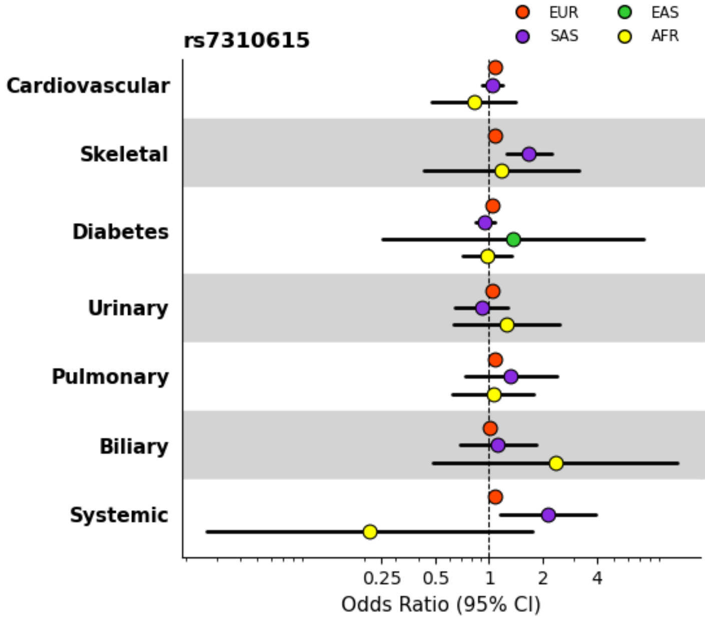

| | Cases (Total samples) | OR (95% CI) | P |
|---|---|---|---|
| Cardiovascular | 33,671 (370,381) | 1.07 (1.06-1.09) | 9.200e-18 |
| | 1,194 (10,038) | 1.04 (0.91-1.19) | 0.579 |
| | 344 (3,784) | 0.82 (0.48-1.40) | 0.466 |
| Skeletal | 28,886 (168,869) | 1.08 (1.06-1.11) | 4.190e-12 |
| | 205 (2,255) | 1.67 (1.24-2.24) | 6.221e-04 |
| | 87 (957) | 1.17 (0.43-3.17) | 0.755 |
| Diabetes | 47,324 (356,334) | 1.04 (1.02-1.05) | 8.540e-06 |
| | 1,856 (10,038) | 0.95 (0.84-1.07) | 0.397 |
| | 151 (1,661) | 1.36 (0.25-7.35) | 0.721 |
| | 3,544 (11,809) | 0.97 (0.71-1.33) | 0.859 |
| Urinary | 47,406 (546,819) | 1.04 (1.02-1.06) | 1.380e-05 |
| | 208 (2,288) | 0.91 (0.64-1.28) | 0.575 |
| | 161 (1,771) | 1.25 (0.63-2.47) | 0.526 |
| Pulmonary | 6,723 (53,167) | 1.07 (1.03-1.12) | 3.807e-04 |
| | 57 (627) | 1.32 (0.73-2.40) | 0.359 |
| | 169 (8,537) | 1.05 (0.62-1.78) | 0.844 |
| Biliary | 12,937 (78,586) | 1.00 (0.96-1.04) | 0.921 |
| | 89 (979) | 1.12 (0.69-1.83) | 0.644 |
| | 33 (363) | 2.35 (0.48-11.36) | 0.290 |
| Systemic | 11,300 (46,630) | 1.08 (1.02-1.14) | 0.006 |
| | 48 (528) | 2.15 (1.16-3.99) | 0.015 |
| | 42 (462) | 0.21 (0.03-1.75) | 0.150 |

## h) rs8032939

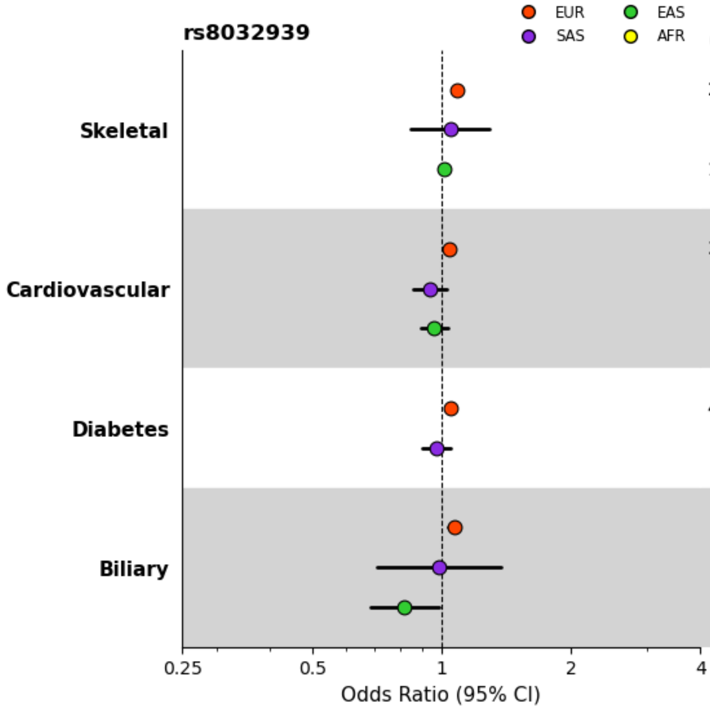

| | Cases (Total samples) | OR (95% CI) | P |
|---|---|---|---|
| Skeletal | 28,886 (168,869) | 1.09 (1.06-1.12) | 3.420e-12 |
| | 205 (2,255) | 1.05 (0.85-1.29) | 0.662 |
| | 11,025 (173,633) | 1.02 (1.02-1.02) | 0.017 |
| Cardiovascular | 33,671 (370,381) | 1.04 (1.02-1.06) | 1.140e-05 |
| | 1,194 (10,038) | 0.94 (0.86-1.03) | 0.186 |
| | 1,673 (157,213) | 0.96 (0.90-1.03) | 0.296 |
| Diabetes | 47,324 (356,334) | 1.05 (1.03-1.07) | 1.840e-08 |
| | 1,856 (10,038) | 0.98 (0.90-1.05) | 0.529 |
| Biliary | 12,937 (78,586) | 1.07 (1.03-1.11) | 9.300e-05 |
| | 89 (979) | 0.99 (0.71-1.38) | 0.936 |
| | 487 (963) | 0.82 (0.68-0.99) | 0.034 |

**i) rs11089637**

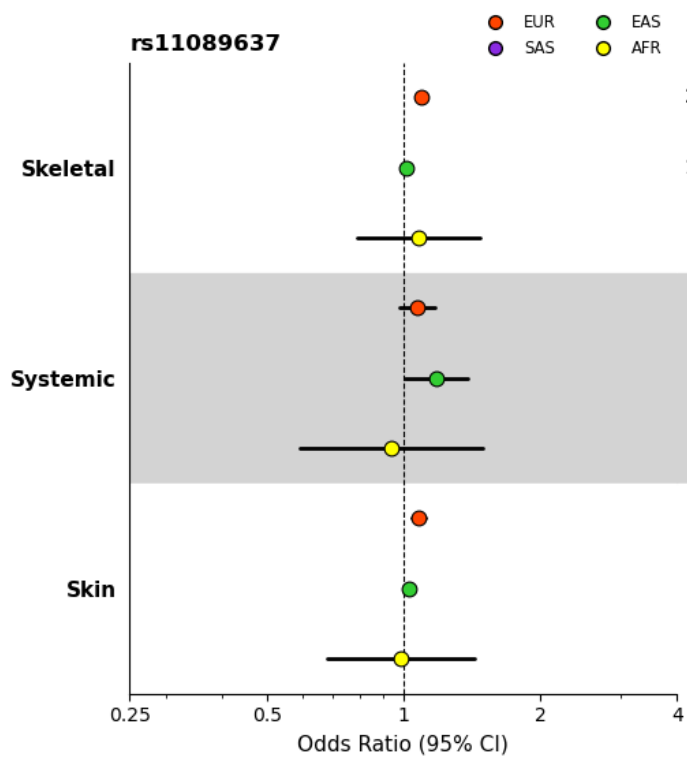

| | Cases (Total samples) | OR (95% CI) | P |
|---|---|---|---|
| Skeletal | 28,886 (168,869) | 1.09 (1.06-1.13) | 2.220e-09 |
| | 11,025 (173,633) | 1.02 (1.01-1.02) | 0.010 |
| | 87 (957) | 1.08 (0.79-1.48) | 0.630 |
| Systemic | 11,300 (46,630) | 1.07 (0.98-1.17) | 0.121 |
| | 303 (175,902) | 1.18 (1.01-1.39) | 0.040 |
| | 42 (462) | 0.94 (0.59-1.49) | 0.799 |
| Skin | 20,285 (97,176) | 1.08 (1.05-1.12) | 7.200e-06 |
| | 4,222 (12,653) | 1.03 (1.03-1.03) | 0.029 |
| | 66 (726) | 0.99 (0.68-1.43) | 0.942 |

**j) rs113477191**

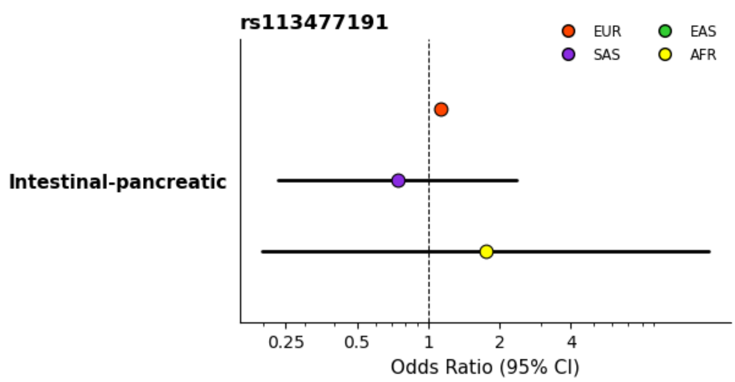

| | Cases (Total samples) | OR (95% CI) | P |
|---|---|---|---|
| Intestinal-pancreatic | 49,710 (418,002) | 1.13 (1.08-1.19) | 3.670e-08 |
| | 806 (8,866) | 0.74 (0.23-2.36) | 0.614 |
| | 480 (5,280) | 1.74 (0.20-15.24) | 0.615 |

**k) rs2580350**

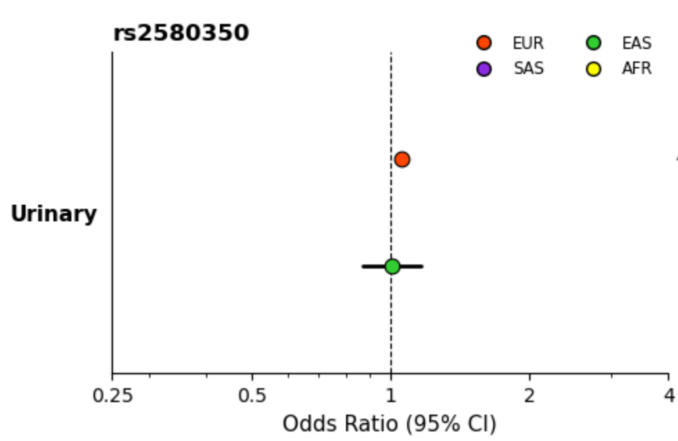

| | Cases (Total samples) | OR (95% CI) | P |
|---|---|---|---|
| Urinary | 47,406 (546,819) | 1.06 (1.04-1.07) | 1.450e-09 |
| | 510 (178,726) | 1.01 (0.87-1.16) | 0.946 |
